# Proteomic markers of colorectal cancer risk: results from the European Prospective Investigation into Cancer and Nutrition

**DOI:** 10.64898/2026.10.05.26364761

**Authors:** Matthew A Lee, Vivian Viallon, David C Muller, Ziqiao Wang, Neil Murphy, James Yarmolinsky, Amanda J. Cross, Pietro Ferrari, Paula Jakszyn, Mazda Jenab, P. Martijn Kolijn, Giovanna Masala, Domenico Palli, Salvatore Panico, Ulrike Peters, Carlotta Sacerdote, Karl Smith-Byrne, Ruth C Travis, Konstantinos K Tsilidis, Rosario Tumino, Bethany Van Guelpen, Roel Vermeulen, Monique Verschuren, Nicholas Wareham, Nilanjan Chatterjee, Elizabeth A Platz, Elio Riboli, Marc J. Gunter

## Abstract

1.

**Background:** Circulating proteins are promising biomarkers of colorectal cancer risk. Most studies combine colon and rectal cancer despite their biological differences.

**Objective:** To identify proteins associated with colorectal, colon, and rectal cancer risk and triangulate findings using cross-cohort confirmation and Mendelian randomisation (MR).

**Design:** We profiled 6,402 proteins, using the SomaScan 7K assay in a case-cohort study nested within the European Prospective Investigation into Cancer and Nutrition (EPIC; 658 colon and 319 rectal cancers). Cox models identified proteins associated with cancer risk at a false discovery rate <5%. Findings were confirmed in the Atherosclerosis Risk in Communities study (ARIC; SomaScan; 235 colon, 36 rectal) and UK Biobank (Olink Explore; 321 colon, 164 rectal). MR and genetic colocalisation evaluated potential causality.

**Results:** We identified 36 proteins associated with colorectal cancer, 50 with colon cancer, and 2 with rectal cancer; 20 of the 36 were also associated with colon cancer and none with rectal cancer, and no protein was associated with both sites. Seven of the 40 colon cancer proteins available in ARIC were confirmed, including COMP, MIC-1/GDF15, TFF3, and THIK. Of these, MIC-1/GDF15 and TFF3 were also confirmed in UK Biobank. COMP was inversely associated with colon cancer risk observationally and in MR analyses.

**Conclusion:** Colon and rectal cancers have different pre-diagnostic proteomic profiles, underlining the need for site-specific analyses. COMP, MIC-1/GDF15, and TFF3 were the most robustly supported biomarkers of colon cancer risk. Future work should prioritise experimental validation of these associations and use of repeated measures to define timing.

**Key messages:** What is already known on this topic: Circulating proteins have been linked to colorectal cancer risk, but few have been validated prospectively and most studies have combined colon and rectal cancer despite their recognised biological differences.

What this study adds: Colon and rectal cancers had distinct pre-diagnostic proteomic profiles with no overlapping associations, and the colorectal cancer signal was driven almost entirely by colon cancer. COMP, MIC-1/GDF15, and TFF3 were associated with colon cancer risk in EPIC and confirmed in independent cohorts, with genetic support for COMP.

How this study might affect research, practice or policy: Proteomic studies of colorectal cancer should be analysed by anatomical site, and COMP, MIC-1/GDF15, and TFF3 warrant experimental validation as markers of colon cancer risk.

## 3. Introduction

Proteins play fundamental roles in human physiology and have been linked to numerous diseases^1^. High-throughput proteomics is emerging as an important tool for identifying proteins for disease prediction^2^, the identification of therapeutic targets^3^, and in improving our understanding of disease development, including cancer^3–6^.

Sex, lifestyle, and genetic factors are linked to colorectal cancer development and progression^7–9^, and a number of circulating proteins associated with risk have been identified using different study designs^5,10–15^. These studies have highlighted the utility of proteins in distinguishing colorectal cancer tumour subtypes^12^, identifying pre-diagnostic biomarkers14, and potential targets of intervention^3,10,16^, yet few protein biomarkers have been validated and the number of prospective studies is limited. Colon and rectal cancers are managed as distinct diseases, with different surgical approaches, neoadjuvant chemoradiotherapy for rectal disease, and different detection by sigmoidoscopy and colonoscopy, and they differ in their somatic mutational landscape and immune behaviour^17–19^. Combining them as a single entity in proteomic analyses risks obscuring site-specific signals.

Here, we performed high-throughput proteomic profiling using the SomaScan 7K Assay in a case-cohort study nested within the European Prospective Investigation into Cancer and Nutrition (EPIC) cohort which included 658 incident colon and 319 incident rectal cancer cases. We then performed confirmation analyses in the Atherosclerosis Risk in Communities (ARIC) study (SomaScan) and UK Biobank (Olink) and compared results with those from genetic analyses.

## 4. Methods

### 4.1. Overview

We estimated the association between pre-diagnostic circulating proteins and colorectal, colon, and rectal cancer risk in EPIC. For proteins associated with cancer risk at a false discovery rate (FDR) p-value threshold in EPIC, we performed confirmatory analyses in ARIC and UK Biobank and assessed potential causal relevance through Mendelian randomisation (MR) analyses. Full details of the methods are available in Supplementary Material. Patients and the public were not involved in the design, conduct, reporting, or dissemination plans of this research.

### 4.2. European Prospective Investigation into Cancer and Nutrition

EPIC is a prospective cohort of >520,000 individuals from 10 European countries, recruited between 1992 and 2000. Participants were aged 35-70 years at recruitment and approximately 75% provided blood samples^20,21^. Incident cancers (coded using ICD-O-3; colon: C180-189 [proximal: C180-185; distal: C186-187]; rectal: C199 and C209) were identified using cancer registries, insurance records, active follow-up, and next of kin. In this analysis, we used a case-cohort study design nested within EPIC which included 5,057 individuals from 14 centres in 4 countries (Italy, Spain, the Netherlands, and the United Kingdom), including 658 and 319 incident cases of colon and rectal cancer, respectively, and 4,080 random subcohort members. Of the 658 colon cancers, 303 were proximal and 304 distal; the remaining 51 could not be assigned to a sub-site (colon not otherwise specified, n= 42; overlapping lesion, n = 9). Colon cancers included 3 appendiceal tumours (C181), which were classified as proximal. Tumour morphology is summarised in the Supplementary Methods.

#### 4.2.1. Proteomic profiling and pre-processing

Blood samples, collected at recruitment (EPIC) were stored in liquid nitrogen and underwent proteomic analysis by Somalogic using the SomaScan 7k Assay according to the manufacturers protocol. Briefly, the SomaScan platform uses modified nucleotides (Slow Off-rate Modified Aptamers) which make direct contact with proteins, enabling detection of proteins or protein complexes and quantifies them in relative fluorescence units (RFUs)^22^. RFUs were normalised by the manufacturer through the following steps (in order): hybridisation normalisation, intraplate median normalisation, plate scaling and calibration, and adaptive normalisation to a population reference.

We (in order): (1) log-transformed the normalised RFUs to reduce skewness; (2) excluded samples where the normalisation scale factor was outside 0.4–2.5; (3) excluded outlier samples using principal component analyses (PCA) and a local outlier factor^23^ using a Tukey rule^24^ modified to account for skewness and multiple testing^25^; (4) applied plate correction using a residual approach whereby, for each aptamer, its measurements were corrected for plate effect estimated in linear mixed effect models adjusted for centre, age, sex, body mass index (BMI), smoking status, and incidence of cancer, cardiovascular disease, type-2 diabetes, neurodegenerative disease, and death, to preserve possible biological variation due to these factors^26^; (5) centred and scaled measurements so that each aptamer had a mean of 0 and standard deviation of 1.

### 4.3. Statistical analysis

Associations of proteins with cancer risk were assessed through Cox proportional hazard models with age as the underlying time scale and Prentice weights and robust variance to account for the case-cohort design. Age at event was age at diagnosis or end of follow-up. We considered a “minimally adjusted” (model 1) and “adjusted” (model 2) model. Models were stratified on sex (for combined analysis only), centre, and age at recruitment (5-year categories), and adjusted for date of blood draw and fasting status. Model 2 additionally included BMI, alcohol, smoking, physical activity, and education. In additional analyses we excluded the first 2 and 5 years of follow-up to investigate potential reverse causation, and included sex and tumour site as interaction terms to investigate heterogeneity by sex and by site. We identified prospectively associated proteins as those for which the FDR adjusted p-value (Benjamini-Hochberg) was <5% and which had consistent directions of effect and overlapping confidence intervals after excluding the first 2-and 5-years of follow-up. All subsequent analyses focussed on these proteins.

#### Cross-cohort confirmation and genetic analyses

ARIC is a prospective community-based cohort of 15,792 individuals from 4 communities (Washington County, Maryland; suburbs of Minneapolis, Minnesota; Forsyth County, North Carolina; Jackson, Mississippi) across the United States of America, recruited between 1987– 1989^27,28^. Participants were aged 45 to 64 years at recruitment. Plasma protein measurements were available for 11,798 participants from plasma collected in Visit 2 in 1990-1992. Incident cancers (coded using ICD-O-3; colon: C180-189; rectal: C199 and C209) were ascertained using registries and active follow-up^29^. In the present work, we used data for 9,495 participants who were at risk for an incident cancer at Visit 2 and who provided appropriate consent. Among them, 235 incident colon and 36 incident rectal cancer cases were identified through 2015. Proteomic profiling (SomaScan 5K) was performed according to the manufacturers protocol; normalisation and adjustment for non-genetic variance are described in the Supplementary method^30,31^. We performed Cox proportional hazard models with time (year) since baseline (Visit 2) as the timescale adjusting for age at baseline, sex, BMI, waist-hip ratio, smoking, pack years, alcohol intake, diabetes, physical activity, a combined variable of race-study centre, top 10 proteomic PEER factors, and the first 10 genetic principal components. Nominal p-values were used in this confirmation analysis.

UK Biobank is a prospective cohort of >500,000 adults from the United Kingdom, recruited between 2006 and 2010^32^. In the present analysis, we used data from the randomly selected individuals included in the UK Biobank Pharma Proteomics Project^33^, which used the Olink proximity extension assay. We excluded individuals with prevalent cancers and processed proteomics data as described for EPIC, except for batch correction which was not deemed necessary. Incident cancers were identified from cancer registry data coded using ICD-10, with ICD-9 complements for the colorectal sites (colon: C180-189 or ICD-9 153 [proximal: C180-185; distal: C186-187]; rectal: C199, C209, or ICD-9 1540 and 1541). Where participants had more than 1 registered cancer, we used the first primary cancer. Data were determined via record linkage to national registries (NHS Digital for England and Wales using participants’ NHS numbers, and NHS Central Register for Scotland using the Community Health Index). Data were available until the censoring date (December 31, 2020, in England and Wales and November 30, 2021, in Scotland) or until participants died, withdrew consent for future linkage or were reported to have left the United Kingdom. A total of 42,271 individuals and 321 incident colon and 164 incident rectal cancers were available for analysis. We performed Cox proportional hazard models to estimate cancer risk using age as the underlying time variable and stratified by sex, centre, and 5-year age group and adjusted for BMI, alcohol, smoking, physical activity, education, and Townsend deprivation index.

We performed two-sample summary-level MR^34,35^ using genome-wide summary-level data for the association between genetic variants and Somalogic derived proteomic data from Ferkingstad et al., (2021)^36^ and for colorectal cancer data from Huyghe et al., (2019)8. We additionally performed genetic colocalisation^37^ to assess the validity of MR assumptions. Instruments were cis variants selected from the protein-coding gene region and a 2Mb window^38^; selection thresholds and the colocalisation assumptions are given in the Supplementary Methods. We used an inverse variance weighted (IVW) multiplicative random effects (IVW-MRE) model; odds ratios (ORs) are interpreted as the change in outcome per normalised SD unit change in the protein. To assess the validity of causal evidence we considered evidence of genetic colocalisation for a shared causal variant (posterior probability ≥ 0.8) and additionally obtained genome-wide summary level-data for Somalogic derived proteins from the smaller Pietzner et al., (2021)^6^, and Zhang et al., (2022)^39^ studies to further compare results in different populations.

## 5. Results

### 5.1. Association between circulating proteins and colorectal cancer risk

In EPIC, after data pre-processing, a total of 5,057 individuals with measurements of 7,289 aptamers, targeting 6,402 unique proteins, were included for analysis. This included 658 colon (distal: 304; proximal: 303) and 319 rectal cancer cases with a median follow-up time of 10 years (SD = 4 years) – baseline characteristics are presented inSupplementary Table 1.

In our main analysis of colorectal cancer, we identified 36 proteins associated with risk (FDR < 0.05; Figure 1 and Supplementary Table 2). Analysing colon and rectal cancer separately identified more associations than the combined analysis, and the colorectal cancer signal was dominated by colon cancer: 20 of the 36 colorectal cancer associated proteins were also associated with colon cancer, none were associated with rectal cancer, and 16 were identified in the colorectal cancer analysis alone. At a nominal threshold (p < 0.05), 28 of the 36 were associated with colon cancer and 14 with rectal cancer. Three of the 36 were also associated with distal colon cancer (CA125, LAMA4, TCAL8) and 1 with proximal colon cancer (EGLN2). Evidence of heterogeneity by sex and by sub-site among the colorectal cancer associated proteins is presented in Supplementary Figure 6 and Supplementary Table 2. Because the colorectal cancer associations were largely attributable to colon cancer, the remainder of the results focus on the site-specific analyses.

**Figure 1.**
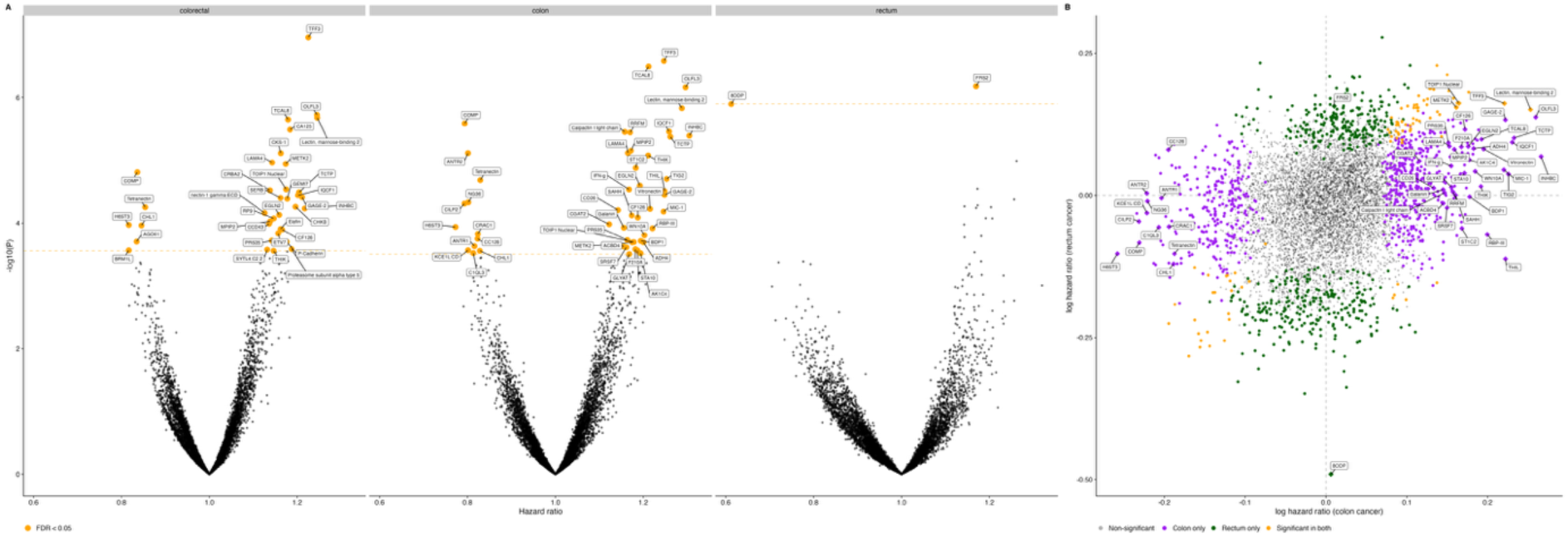
Circulating protein associations with incident colorectal, colon, and rectal cancer risk in EPIC. Associations were estimated using fully-adjusted (model 2) Cox proportional hazards regression with combined sexes and no follow-up exclusion (colorectal: up to 5,057 participants and 977 incident cases; colon: up to 4,742 participants and 658 incident cases; rectum: up to 4,430 participants and 319 incident cases). (A) Volcano plots showing the hazard ratio and −log10 p-value for each protein in the colorectal, colon, and rectal cancer analyses. Highlighted (orange, labelled) points reached the Benjamini-Hochberg false discovery rate threshold (5%), as indicated by the horizontal line; 36, 50, and 2 proteins reached this threshold for colorectal, colon, and rectal cancer, respectively. (B) Comparison of the colon and rectal cancer analyses, with each point giving a protein’s association with incident colon cancer (x-axis) and incident rectal cancer (y-axis), expressed as the log hazard ratio. Points are coloured by nominal significance (p < 0.05): grey = not associated with either cancer; purple = associated with colon cancer only; green = associated with rectal cancer only; orange = associated with both. Proteins passing the false discovery rate threshold (FDR-adjusted p < 0.05) in either analysis are shown as diamonds and labelled; no protein reached FDR significance for both.

### 5.2. Association between circulating proteins and colon and rectal cancer risk

We identified 50 proteins associated with incident colon cancer (FDR < 0.05; Figure 1 and Supplementary Table 2). These associations were consistent across minimally-(model 1) and fully-adjusted (model 2) models and analyses excluding the first 2-and 5-years of follow-up (Supplementary Figure 1 and Supplementary Table 4). We observed a cluster of proteins which were positively correlated with one another (Pearson r > 0.4) in the sub-cohort (Supplementary Figure 2, Supplementary Figure 3, and Supplementary Table 3). Of the 50, 42 were also nominally associated (p < 0.05) with colorectal cancer but only 2 (METK2 and TFF3) with rectal cancer.

We identified 2 proteins associated with incident rectal cancer (FRS2 and 8ODP; FDR < 0.05; Figure 1 and Supplementary Table 2). Although neither was associated with colon cancer at a nominal threshold (p < 0.05) both were nominally associated with colorectal cancer; we did not observe evidence of heterogeneity by sex for either protein. In sex-specific analyses we identified 6 proteins associated with rectal cancer in men and none in women (Supplementary Table 2). Given the smaller number of incident rectal cancer cases (n = 319), these analyses were less well powered than those for colon cancer for the same sized association.

For the 50 proteins associated with incident colon cancer we observed evidence of heterogeneity (p-value < 0.05) by sex and sub-site for 8 (COMP, EGLN2, LAMA4, RBP-III, TCAL8, THIL, TIG2, Vitronectin) and 2 (KCE1L:CD, SAHH) proteins, respectively (Figure 2). In analyses of colon cancer subtypes, we identified 16 proteins associated with incident proximal colon cancer and 7 proteins associated with incident distal colon cancer (Supplementary Table 2, Supplementary Figure 4, and Supplementary Figure 5). Of the 16 proteins associated with proximal colon cancer, 8 (CD26, Calpactin I light chain, EGLN2, IFN-g, KCE1L:CD, RRFM, SAHH, SRSF7) were also associated with colon cancer overall, 2 of which (KCE1L:CD and SAHH) showed evidence of heterogeneity by sub-site in the colon cancer analysis. Of the 7 proteins associated with distal colon cancer, 4 (LAMA4, ST1C2, TCAL8, TIG2) were also associated with colon cancer overall, none of which showed evidence of heterogeneity in the colon cancer analysis. The separation between proximal and distal colon cancer was less complete than that between colon and rectal cancer: at a nominal threshold (p < 0.05), 4 of the 16 proximal colon cancer associated proteins (Calpactin I light chain, CD26, EGLN2, RRFM) were also associated with distal colon cancer, and 3 of the 7 distal colon cancer associated proteins (LAMA4, ST1C2, TCAL8) were also associated with proximal colon cancer, in all cases in the same direction.

**Figure 2.**
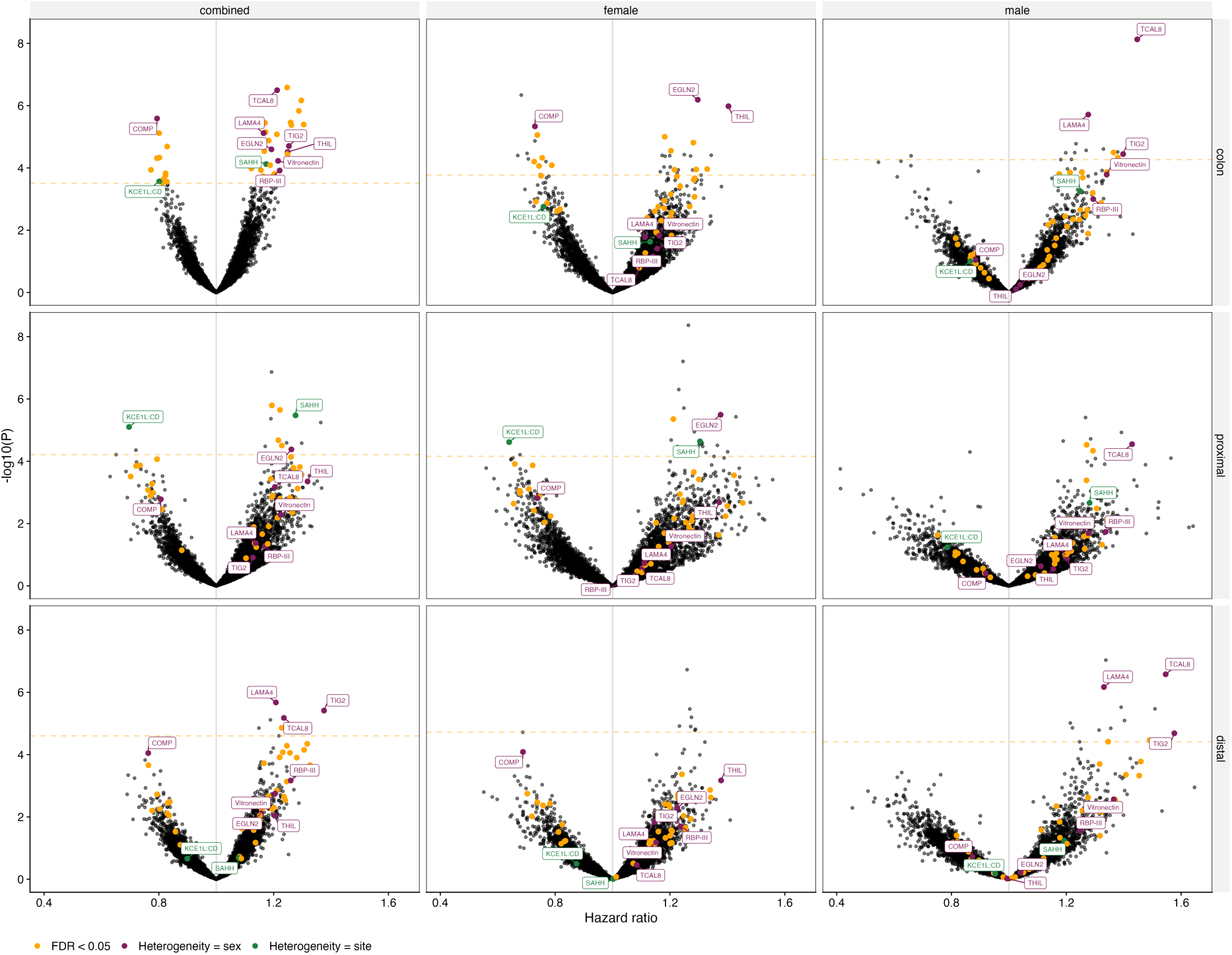
Evidence of heterogeneity by sex and/or by site in Cox proportional hazard models of colon cancer. All panels show the results from the respective sex-cancer analysis and orange points in all panels highlight the 50 proteins associated in the sex-combined colon cancer analysis. Purple points highlight proteins where there was evidence of heterogeneity by sex; green points highlight proteins where there was evidence of heterogeneity by site; blue points highlight proteins where there was evidence of heterogeneity by sex and site; the horizontal line (orange) indicates the false discovery rate (FDR) p-value threshold for the respective analysis.

In sex-specific analyses, we identified 35 and 14 proteins associated with colon cancer in women and men, respectively (Supplementary Table 2, Supplementary Figure 4, and Supplementary Figure 5). Of the 8 proteins with evidence of heterogeneity by sex in the sex-combined analysis, 3 proteins (COMP, EGLN2, and THIL) were associated with colon cancer in women and 3 proteins (LAMA4, TCAL8, and TIG2) were associated with colon cancer in men. All of sex-and site-specific associations were consistent after excluding the first 2-and 5-years of follow-up (Supplementary Figure 4).

### 5.3. Cross-cohort comparisons and genetic analysis

#### 5.3.1. Atherosclerosis Risk in Communities study

Of the 50 colon cancer associated proteins, 40 had been measured using the same aptamers in the SomaScan 5k in ARIC, as had both of the rectal cancer associated proteins and 19 of the 36 colorectal cancer associated proteins (Supplementary Table 5 and Figure 3). For colon cancer, 7 proteins reached the nominal p-value threshold. Four proteins (IQCF1, MIC-1/GDF15, TFF3, THIK) were consistently positively associated with risk of colon cancer in both EPIC and ARIC, 2 (COMP and Tetranectin/CLEC3B) were inversely associated with risk, and 1 protein (MPIP2) showed discordant associations (higher risk in EPIC and a lower risk in ARIC). Neither of the rectal cancer associated proteins reached the nominal p-value threshold and only FRS2 had a consistent direction of association with the EPIC analysis. For colorectal cancer, 3 proteins reached the nominal p-value threshold: TFF3 and THIK were positively associated with risk and Tetranectin/CLEC3B was inversely associated, all consistent in direction with EPIC.

**Figure 3.**
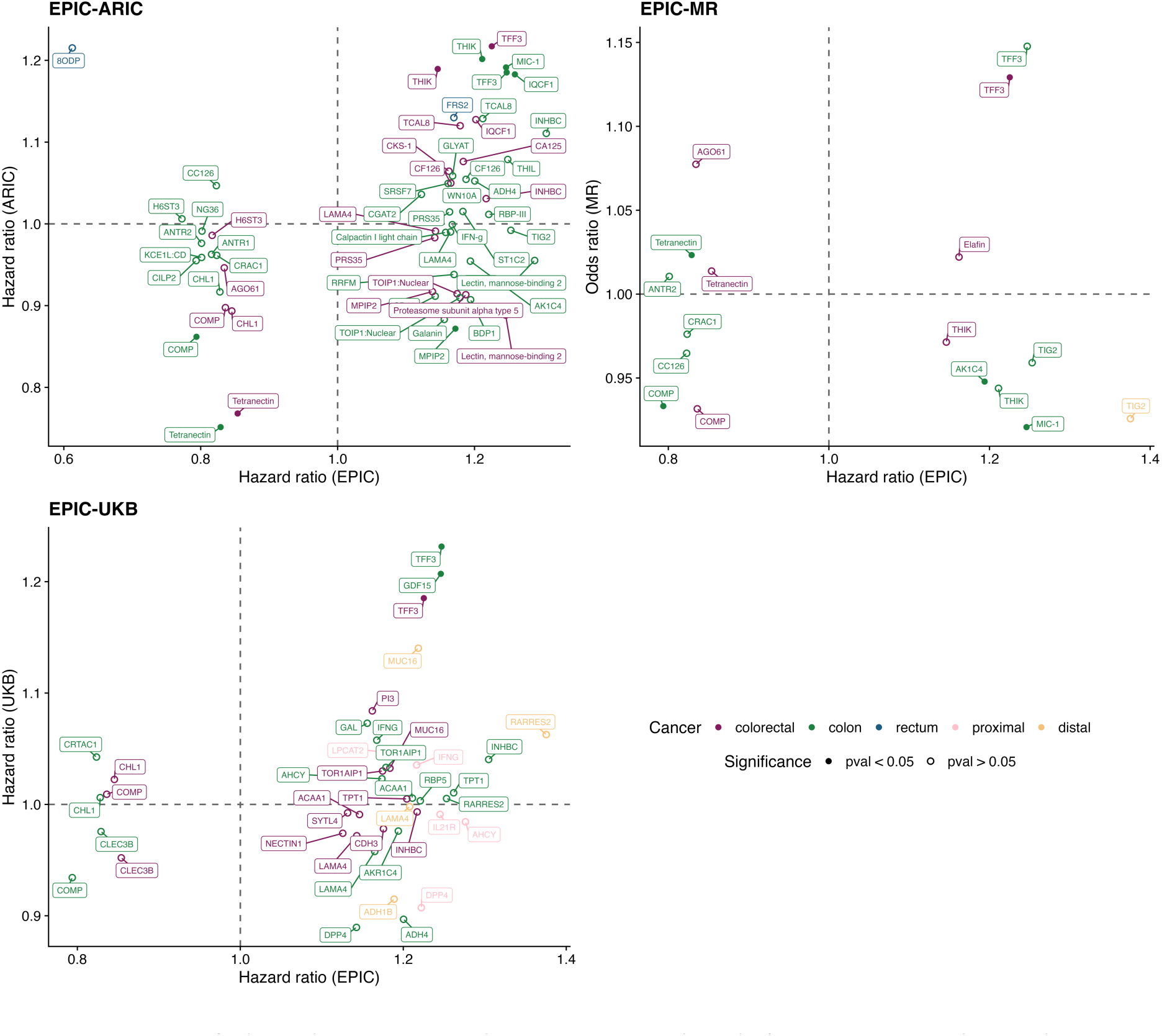
Comparison of colorectal cancer associated proteins in EPIC with results from ARIC, UKB, and MR analyses. Scatter plots show the hazard ratios (EPIC, ARIC, and UKB) and odds ratios (MR) for proteins associated with colorectal, colon, rectal, and distal colon cancer in the main analysis performed in the European Prospective Investigation into Cancer and Nutrition (EPIC) that were tested in the Atherosclerosis Risk in Communities (ARIC) study, the UK Biobank (UKB), and in Mendelian randomisation (MR) analyses using genome-wide summary-level data. Solid points indicate a nominal p-value threshold (0.05) was reached in the non-EPIC analysis. In the EPIC-ARIC and EPIC-UKB panels only proteins with consistent directions of effect in the 2 studies are labelled; in the EPIC-MR panel all points are labelled. NB: Tetranectin (EPIC, ARIC, and MR) and CLEC3B (UKB) are the same protein.

#### 5.3.2. UK Biobank

Of the 50 colon cancer associated proteins, 19 were available in the Olink Explore panel used in UK Biobank, as were 14 of the 36 colorectal cancer associated proteins; none of the rectal cancer associated proteins were available (Supplementary Table 5 and Figure 3). For colon cancer, 2 proteins (MIC-1/GDF15, TFF3) reached the nominal p-value threshold (0.05), both associated with a higher risk in EPIC and UK Biobank. IQCF1, which was positively associated with colon cancer risk in ARIC, was not measured in UK Biobank. THIK, COMP, and Tetranectin/CLEC3B were measured and showed the same direction of association as in EPIC and ARIC, but none reached the nominal p-value threshold. We were able to test 5 of the 16 proximal colon cancer associated proteins and 4 of the 7 distal colon cancer associated proteins in UK Biobank, but there was little consistency in directions of association and none reached the nominal p-value threshold. For colorectal cancer, only TFF3 reached the nominal p-value threshold, and was associated with a higher risk in both EPIC and UK Biobank.

#### 5.3.3. Genetic analyses

We were able to test 10 of the 50 colon cancer associated proteins and 6 of the 36 colorectal cancer associated proteins in MR and genetic colocalisation analyses (Figure 3, Supplementary Table 6, Supplementary Table 7, and Supplementary Table 8). For colon cancer, 4 proteins reached the nominal p-value threshold (0.05): COMP was associated with a lower risk, consistent with the observational analyses; Tetranectin/CLEC3B was associated with a higher risk in the MR analysis but a lower risk in EPIC, ARIC, and UK Biobank; AK1C4 was associated with a lower risk in the MR and ARIC analyses but a higher risk in EPIC; and MIC-1/GDF15 was associated with a lower risk in the MR analysis but a higher risk in EPIC, ARIC, and UK Biobank. For colorectal cancer, TFF3 reached the nominal p-value threshold and was associated with a higher risk, consistent with the EPIC, ARIC, and UK Biobank analyses. In additional analyses using genome-wide summary-level data from 2 independent proteomic genome-wide association studies (Pietzner et al. and Zhang et al.) we found similar evidence (p-value < 0.05) for a lower risk of colon cancer for Tetranectin/CLEC3B and a higher risk of colon cancer for (Supplementary Figure 7 and Supplementary Table 8). We did not observe evidence of genetic colocalisation for any protein (posterior probability for a shared causal variant < 0.8).

## 6. Discussion

Using the SomaScan 7K assay in a large prospective case-cohort study nested within EPIC, we identified 36 proteins associated with incident colorectal cancer. Analysing the anatomical sites separately identified 50 proteins associated with incident colon cancer and 2 associated with incident rectal cancer, with no overlap between sites, showing that the colorectal cancer signal was driven almost entirely by colon cancer. The distinct proteomic profiles of colon and rectal cancer, together with evidence of sex-and sub-site-specific associations, underscore the biological heterogeneity of colorectal cancer and the importance of site-specific analyses. Cross-cohort confirmation in ARIC and UK Biobank provided the strongest evidence for MIC-1/GDF15 and TFF3 while genetic analyses provided additional support for COMP as prospective biomarkers of colon cancer risk.

The absence of overlap between proteins associated with colon and rectal cancer is consistent with the recognised molecular, pathological, and epidemiological distinctions between these cancers^17–19^, yet they are frequently combined in epidemiological studies. The limited confirmation of the 2 rectal cancer-associated proteins in ARIC – where the number of incident rectal cancer cases was small (n=36) – means that statistical power was a likely constraint and these findings should be interpreted cautiously. The inability to test these proteins in UK Biobank (due to platform differences) further limits conclusions about rectal cancer-specific protein biomarkers, and larger prospective studies with sufficient rectal cancer cases are needed. Proximal and distal colon cancers showed only partially overlapping proteomic associations. The absence of overlap at the false discovery rate threshold does not imply complete independence: at a nominal threshold (p < 0.05), CA125 was associated with both distal colon and rectal cancer, and PCAT2 with both proximal colon and rectal cancer, in each case in the same direction. This partial overlap is consistent with the shared embryonic origin, luminal exposures, and modifiable risk factors of these cancers, and suggests the site-specific profiles differ in degree rather than being wholly distinct. These findings suggest site-specific analyses should be the focus of future work.

Across all sex-and site-specific analyses we identified 241 associations involving 139 unique proteins. Using curated UniProt annotations (Supplementary Table 9), we organised the 50 proteins associated with colon cancer risk into broad functional categories (Supplementary Table 10). The associated proteins spanned several recurring biological themes, including mucosal epithelial and digestive function, extracellular matrix and tissue remodelling, immune, inflammatory and cancer-antigen signalling, and metabolic enzymes involved in lipid and small-molecule handling. Representative examples illustrate these themes: TFF3, a mediator of gastrointestinal epithelial maintenance and mucosal repair; MIC-1/GDF15, a stress-responsive cytokine implicated in immune and inflammatory signalling; and COMP, an extracellular matrix protein best known for maintaining the structural integrity of cartilage.

Trefoil factor 3 (TFF3; UniProt: Q07654) is primarily expressed in goblet cells and interacts with mucin glycoproteins in the maintenance and repair of intestinal mucosa^40^. TFF3 levels differ in colorectal cancer cases and non-cases^41^ and recent work has highlighted this protein as a predictive biomarker^42^. Our analysis in EPIC supports TFF3 as a potential marker of risk as we saw consistent evidence of an increased risk of colon and colorectal cancer after excluding the first 2-and 5-years of follow-up. These results were supported by evidence from ARIC, UK Biobank, and MR analyses. In EPIC we also measured MUCL1 (UniProt: Q96DR8), a mucin like glycoprotein, and observed nominal evidence (p-value < 0.05) of an increased risk of colon and colorectal cancer.

Previous work has shown evidence that stromal extracellular matrix dysregulation may contribute to colorectal cancer development^43^. Higher levels of COMP (UniProt: P49747) and Tetranectin/CLEC3B (UniProt: P05452) were associated with a lower risk of colon cancer in both EPIC and ARIC, and Tetranectin/CLEC3B was also associated with a lower risk of colorectal cancer in both studies; our MR analysis supported the association between COMP and colon cancer risk. In UK Biobank both proteins were associated with a lower risk of colon cancer but neither reached the nominal p-value threshold. A related protein, CILP2, was associated with a lower risk of colon cancer in EPIC and had a consistent direction of association in ARIC, although it did not reach the nominal p-value threshold. Biologically, COMP and Tetranectin/CLEC3B are linked through their roles in extracellular matrix organisation and remodelling: COMP functions as a matricellular protein that supports collagen assembly and cell-matrix interactions^44^, while Tetranectin binds plasminogen and influences pericellular proteolysis and matrix turnover^45^. This protective association stands in contrast to the role of GREM1, a potent antagonist of Bone Morphogenetic Proteins (BMPs), which previous MR analyses has linked with increased risk of colon and colorectal cancer^16^. Since BMP signalling is required to maintain epithelial differentiation and suppress tumorigenesis, the elevated levels of GREM1 associated with increased cancer risk may represent a loss of the signalling environment that normally facilitates the healthy matrix organisation provided by COMP and Tetranectin/CLEC3B.

THIK (UniProt: P09110), also known as ACAA1 (3-ketoacyl-CoA thiolase, peroxisomal), was associated with a higher risk of colon and colorectal cancer in EPIC and was confirmed in ARIC. It is a cleavage enzyme involved in fatty acid beta-oxidation in peroxisomes^46^. This process yields acetyl-CoA, which is subsequently used for energy production and various biosynthetic processes; normal functioning of this pathway is essential for maintaining overall lipid turnover and systemic lipid homeostasis. Previous work has shown that docosahexaenoic acid (DHA) – a very-long-chain polyunsaturated fatty acid – is associated with an increased risk of colorectal cancer and that this increased risk may be observed during early adulthood^47^. Additional research has identified a causal relationship between a number of fatty acids and colorectal cancer risk^48^, however, the directionality and causality of these associations are uncertain and warrant mechanistic investigation, particularly to distinguish whether THIK levels are a driver of tumorigenesis or a compensatory response to altered fatty acid composition.

Blood was collected a median of 10 years before diagnosis, so these proteins are candidate markers of risk rather than of early detection, and would be relevant to deciding who is screened and how often rather than to diagnosis. Our confirmation analysis in ARIC and comparison with UK Biobank were limited by available case numbers and by the proteins measured on each platform, and not all proteins identified in EPIC could be tested by MR. We did not have information on stage at diagnosis; many SomaScan analytes are produced or cleared by the liver, so advanced disease at blood draw could in principle distort levels, although the consistency of estimates after excluding 2-and 5-years of follow-up argues against this. Although MR analyses were not supported by genetic colocalisation, the broad consistency across studies suggests a common factor may underlie a number of associations. We present all sex-and site-specific analyses for completeness, but caution over-interpretation given the limited case numbers for some.

## Conclusion

Colon and rectal cancers had distinct pre-diagnostic proteomic profiles, and the colorectal cancer signal was driven almost entirely by colon cancer. COMP, MIC-1/GDF15, and TFF3 are the most robustly supported candidate markers of colon cancer risk. Establishing whether they are clinically useful will require testing whether they add to existing risk models, in whom, and over what interval, alongside experimental validation and replication in cohorts with broader protein coverage and repeated measures.

## 7. Legends

*Supplementary Figure 1 Comparison of models and follow-up exclusions for proteins associated with colorectal, colon, and rectal cancer risk in the main analysis. Forest plots show the hazard ratio and 95% confidence interval for each protein reaching the false discovery rate threshold (FDR-adjusted p < 0.05) in the sex-combined analysis of colorectal (36 proteins), colon (50 proteins), or rectal (2 proteins) cancer. Proteins are grouped by cancer and labelled as cancer: protein. (Left) Comparison of the minimally-adjusted (model 1) and fully-adjusted (model 2) models, with no follow-up exclusion. (Right) Comparison of the fully-adjusted model (model 2) with no exclusion and after excluding the first 2-and 5-years of follow-up. Filled points indicate FDR-adjusted p < 0.05 in the analysis shown*.

*Supplementary Figure 2 Correlation heatmaps for proteins associated with cancer risk within each cancer-sex analysis. Supplementary Figure 2 is a multi-page PDF with one page for each cancer (colorectal, colon, rectum, proximal colon, distal colon) and sex (combined, female, male) combination. Each page shows 3 heatmaps of the pairwise Pearson correlations between the proteins reaching the false discovery rate threshold (FDR-adjusted p < 0.05) in that analysis, calculated among all participants, among cases only, and among non-cases only, using pairwise complete observations. Correlations are reported in Supplementary Table 3*.

*Supplementary Figure 3 Correlation networks for proteins associated with cancer risk within each cancer-sex analysis. Supplementary Figure 3 is a multi-page PDF with one page for each cancer (colorectal, colon, rectum, proximal colon, distal colon) and sex (combined, female, male) combination. Each page shows 3 network maps, calculated among all participants, among cases only, and among non-cases only, in which proteins are connected where the pairwise Pearson correlation exceeds 0.4; edge width and shading are scaled to the strength of the correlation*.

*Supplementary Figure 4 Comparison of models and follow-up exclusions for proteins associated with cancer risk within each cancer-sex analysis. Supplementary Figure 4 is a multi-page PDF with one page for each cancer (colorectal, colon, rectum, proximal colon, distal colon) and sex (combined, female, male) combination. Each page shows 2 forest plots for the proteins reaching the false discovery rate threshold (FDR-adjusted p < 0.05) in that analysis: (left) a comparison of the fully-adjusted model (model 2) with no exclusion and after excluding the first 2-and 5-years of follow-up, and (right) a comparison of the minimally-adjusted (model 1) and fully-adjusted (model 2) models with no follow-up exclusion. Filled points indicate FDR-adjusted p < 0.05. Combinations with no proteins reaching the threshold are shown as an empty page*.

*Supplementary Figure 5 Cox proportional hazard models of the association between 7,289 aptamers, targeting 6,402 unique proteins, and all cancer-sex combinations. The volcano plot shows the hazard ratio and -log10 p-value for each aptamer. Highlighted (orange) points reached the Benjamini-Hochberg false discovery rate p-value threshold (5%) as indicated by the horizontal line. Labelled points are proteins associated (FDR) in ≥ 3 cancer-sex analyses*.

*Supplementary Figure 6 Evidence of heterogeneity by sex and/or by site in Cox proportional hazard models of colorectal cancer. All panels show the results from the respective sex-cancer analysis and orange points in all panels highlight the 36 proteins associated in the sex-combined colorectal cancer analysis. Purple points highlight proteins where there was evidence of heterogeneity by sex; green points highlight proteins where there was evidence of heterogeneity by site; blue points highlight proteins where there was evidence of heterogeneity by sex and site; the horizontal line (orange) indicates the false discovery rate (FDR) p-value threshold for the respective analysis*.

*Supplementary Figure 7 Comparison of colorectal cancer associated proteins in EPIC with results from MR analyses using different genome-wide summary-level datasets to identify protein instruments. Scatter plots show the hazard ratio (EPIC) and odds ratios (MR) for proteins associated with colorectal, colon, rectal, proximal colon, and distal colon cancer, with 1 panel per protein-QTL dataset (Ferkingstad et al., 2021; Pietzner et al., 2021; Zhang et al., 2022). Solid points indicate a nominal p-value threshold (0.05) was reached in the MR analysis*.

*Supplementary Table 1 Baseline characteristics by cancer site and sex group. Baseline characteristics are shown for each cancer site and sex-specific analysis group. Continuous variables are reported as median (SD); categorical variables are reported as n (%), among cases unless otherwise indicated*.

*Supplementary Table 2 Proteins associated with cancer site and sex groups at FDR < 0.05 in the primary EPIC analysis. Columns: protein, SomaLogic protein name; UNIPROT, UniProt accession; cancer, cancer outcome; sex, analysis sex group; HR, hazard ratio; CI_lower/CI_upper, 95% confidence interval; pval, nominal P value; pval_FDR, FDR-adjusted P value; het_sex, sex-heterogeneity P value where applicable; het_site, anatomical-site heterogeneity P value where applicable*.

*Supplementary Table 3 Pairwise Pearson correlations between proteins associated with cancer risk within each cancer-sex analysis. For each cancer and sex combination in which at least 1 protein reached the false discovery rate threshold (FDR-adjusted p < 0.05), correlations were calculated between all pairs of those proteins using pairwise complete observations, separately among all participants, among cases only, and among non-cases only. Thirteen of the 15 cancer-sex combinations are represented; proximal colon cancer in men and rectal cancer in women are absent because no protein reached the threshold in those analyses. Columns: cancer, cancer outcome; sex, analysis sex group; subset, whether the correlation was calculated among all participants, cases only, or non-cases only; n_samples, number of participants contributing to the correlation; n_proteins, number of proteins included in that analysis; protein_1/protein_2, the protein pair; correlation, Pearson correlation coefficient. Correlations are shown graphically in Supplementary Figure 2, and as networks for pairs with a correlation exceeding 0.4 in Supplementary Figure 3*.

*Supplementary Table 4 All Cox proportional hazards results in EPIC across models and follow-up exclusions. Results are reported for all 7,289 aptamers, targeting 6,402 unique proteins, in each of the 15 cancer-sex analyses (colorectal, colon, rectum, proximal colon, and distal colon, each for combined sexes, women, and men), under the minimally-adjusted (model 1) and fully-adjusted (model 2) models, and with no follow-up exclusion and after excluding the first 2-and 5-years of follow-up. Unlike Supplementary Table 2, this table is not restricted to proteins reaching the false discovery rate threshold. Columns: protein, SomaLogic protein name; UNIPROT, UniProt accession; cancer, cancer outcome; sex, analysis sex group; model, 1 for the minimally-adjusted and 2 for the fully-adjusted model; followup, number of years of follow-up excluded (0 indicates no exclusion); HR, hazard ratio; CI_lower/CI_upper, 95% confidence interval; pval, nominal P value; pval_FDR, Benjamini-Hochberg FDR-adjusted P value*.

*Supplementary Table 5 Comparison of protein-cancer associations across EPIC, ARIC, and UK Biobank for all FDR-significant proteins within EPIC. Only proteins available in at least 1 comparison cohort are listed. Where ARIC measured a protein with an additional aptamer that did not match the aptamer used in EPIC, that row is labelled protein (SeqId) to distinguish it from the matched measure*.

*Supplementary Table 6 Mendelian randomisation estimates for protein effects on cancer outcomes across external protein-QTL studies and MR methods for all FDR-significant proteins within EPIC. Columns: protein, SomaLogic protein name; study, the author, year, and PubMed ID of the protein-QTL study used to identify instruments; UNIPROT, UniProt accession; cancer, cancer outcome; sex, analysis sex group; method, MR method; OR, odds ratio; CI_lower/CI_upper, 95% confidence interval; pval, nominal P value; nsnp, number of variants in the instrument; n_exposure/n_outcome, sample sizes of the exposure and outcome genome-wide association studies*.

*Supplementary Table 7 Colocalisation posterior probabilities for protein-cancer genetic association signals across external protein-QTL studies for all FDR-significant proteins within EPIC. Posterior probabilities are reported for each hypothesis (h0 to h4), where h4 is the posterior probability of a shared causal variant, together with the prior probabilities used*.

*Supplementary Table 8 Comparison of cancer associated proteins in EPIC with results from ARIC, UKB, and MR analyses. Scatter plots show the hazard ratios (EPIC, ARIC, and UKB) and odds ratios (MR) for proteins associated with colorectal, colon, rectal, proximal colon, and distal colon cancer in the main analysis performed in the European Prospective Investigation into Cancer and Nutrition (EPIC) that were tested in the Atherosclerosis Risk in Communities (ARIC) study, the UK Biobank (UKB), and in Mendelian randomisation (MR) analyses using genome-wide summary-level data. Solid points indicate a nominal p-value threshold (0.05) was reached in the non-EPIC analysis. In the EPIC-ARIC and EPIC-UKB panels only proteins with consistent directions of effect in the 2 studies are labelled; in the EPIC-MR panel all points are labelled. NB: Tetranectin (EPIC, ARIC, and MR) and CLEC3B (UKB) are the same protein*.

*Supplementary Table 9 Protein annotation information for FDR-significant proteins. This table lists unique proteins identified in Supplementary Table 2 and provides functional annotation from UniProtKB. Cancer-sex combinations indicate all analyses in which each protein was FDR-significant; UNIPROT: UniProt accession identifier; cancer-sex: Cancer and sex analysis combinations where the protein was identified as FDR-significant; function_summary: UniProt functional description of the protein; tissue_specificity: UniProt tissue-specificity annotation, where available; disease_involvement: UniProt disease association annotation, where available; go_biological_process: Gene Ontology biological process terms; go_molecular_function: Gene Ontology molecular function terms; go_cellular_component: Gene Ontology cellular component terms*.

*Supplementary Table 10 Broad functional categories of colon cancer associated proteins. This table summarises the proteins identified in the colon cancer, sex-combined analysis by broad functional category. Categories were assigned using the functional, tissue, disease, and gene ontology annotations reported in Supplementary Table 9. Category: broad biological category assigned to the proteins; summary: brief description of the shared biological features or annotation themes supporting the category*.

## 8. Declarations

### 8.1. Ethics approval

The EPIC study was conducted in accordance with the Declaration of Helsinki. The study was approved by the local ethical committees in participating countries and the IARC ethical committee. All participants provided written informed consent for data collection and storage, as well as individual follow-up before study entry. This study is listed at clinicaltrials.gov as NCT03285230. Approval for the ARIC study was received from the Institutional Review Board at each study centre. All participants provided informed consent.

### 8.2. Author contributions

MAL: Formal analysis; Investigation; Project administration; Writing – original draft; VV: Data curation; Methodology; Project administration; Supervision; Writing – review and editing; DCM: Methodology; Project administration; Writing – review and editing; ZW: Formal analysis; Writing – review and editing; ER: Conceptualization; Funding acquisition; Resources; Writing – review and editing; MJG: Conceptualization; Funding acquisition; Resources; Supervision; Writing – review and editing; NM: Supervision; JY: Writing – review and editing; AJC: Writing – review and editing; MJ: Writing – review and editing; UP: Writing – review and editing; KKT: Writing – review and editing; PJ: Funding acquisition; Resources; Writing – review and editing; PF: Funding acquisition; Resources; Writing – review and editing GM: Funding acquisition; Resources; Writing – review and editing; DP: Funding acquisition; Resources; Writing – review and editing; SP: Funding acquisition; Resources; Writing – review and editing; CS: Funding acquisition; Resources; Writing – review and editing; KSB: Funding acquisition; Resources; Writing – review and editing; RCT: Funding acquisition; Resources; Writing – review and editing; RT: Funding acquisition; Resources; Writing – review and editing; PMK: Validation; Writing – review and editing; RV: Funding acquisition; Resources; Writing – review and editing; MV: Funding acquisition; Resources; Writing – review and editing; NW: Funding acquisition; Resources; Writing – review and editing; NC: Funding acquisition; Resources; Writing – review and editing; EAP: Funding acquisition; Resources; Writing – review and editing; BVG: Funding acquisition; Resources; Writing – review and editing

## Data Availability

All data produced in the present work are contained in the manuscript. ARIC data, including the data used in this analysis, can be accessed by the following policy: https://sites.cscc.unc.edu/aric/sites/default/files/public/listings/ARIC%20data%20sharing.pdf. ARIC data also are available via BioLINCC (controlled access database).

## 8.3. Acknowledgments

The coordination of EPIC-Europe is financially supported by International Agency for Research on Cancer (IARC) and also by the Department of Epidemiology and Biostatistics, School of Public Health, Imperial College London which has additional infrastructure support provided by the NIHR Imperial Biomedical Research Centre (BRC). The national cohorts are supported by Associazione Italiana per la Ricerca sul Cancro-AIRC-Italy, Italian Ministry of Health, Italian Ministry of University and Research (MUR), Compagnia di San Paolo (Italy); Dutch Ministry of Public Health, Welfare and Sports (VWS), the Netherlands Organisation for Health Research and Development (ZonMW), World Cancer Research Fund (WCRF), (The Netherlands); Instituto de Salud Carlos III (ISCIII), Regional Governments of Andalucía, Asturias, Basque Country, Murcia and Navarra, and the Catalan Institute of Oncology - ICO (Spain); Cancer Research UK (C864/A14136 to EPIC-Norfolk; C8221/A29017 to EPIC-Oxford), Medical Research Council (MR/N003284/1, MC-UU_12015/1 and MC_UU_00006/1 to EPIC-Norfolk; MR/Y013662/1 to EPIC-Oxford) (United Kingdom). Previous support has come from “Europe against Cancer” Programme of the European Commission (DG SANCO). The generation of the proteomic data was partly funded by the Michael J Fox Foundation (#008994 to Christina M. Lill and Elio Riboli), the Cure Alzheimer’s Fund (to Christina M. Lill and Lars Bertram), the ‘CReATe-Clinical Research in ALS and Related Disorders for Therapeutic Development’ Consortium (to Christina M. Lill and Lars Bertram), with additional grant support from the Heisenberg program of the Deutsche Forschungsgemeinschaft (DFG; LI 2654/4-1 to Christina M. Lill). SomaScan® data were generated under Master Research Agreement, 14th December 2021, between Imperial College London and SomaLogic Inc. SomaLogic were not involved in analyzing or interpreting the data; or in writing or submitting the manuscript for publication.

The authors thank the National Institute for Public Health and the Environment (RIVM), Bilthoven, the Netherlands, for their contribution and ongoing support to the EPIC Study.

The authors thank the staff and participants of the ARIC study for their important contributions. Cancer data were provided by the Maryland Cancer Registry, Center for Cancer Prevention and Control, Maryland Department of Health, with funding from the State of Maryland and the Maryland Cigarette Restitution Fund. The collection and availability of cancer registry data are also supported by the Cooperative Agreement NU58DP007114, funded by the Centers for Disease Control and Prevention. Its contents are solely the responsibility of the authors and do not necessarily represent the official views of the Centers for Disease Control and Prevention or the Department of Health and Human Services.

This research has been conducted using the UK Biobank Resource under Application Number 97846.

## 9. Code and data availability

Proteomic pre-processing was performed using R (4.1.2)^49^; local outlier factor based PCA was implemented using bigutilsr^50^ (0.3.8) and plate correction was performed as described in Viallon et al., (2021)^26^ using: pcpr2^51^ (0.0.0.1), lme4^52^ (1.1-35.5), nlme^53,54^ (3.1-166). All analyses were performed using R (4.4.1). All data produced in the present work are contained in the manuscript. ARIC data, including the data used in this analysis, can be accessed by the following policy: https://sites.cscc.unc.edu/aric/sites/default/files/public/listings/ARIC%20data%20sharing.pdf. ARIC data also are available via BioLINCC (controlled access database).

## 10. Supplementary data

Supplementary Data are available online

## 11. Funding

The coordination of EPIC-Europe is financially supported by International Agency for Research on Cancer (IARC) and also by the Department of Epidemiology and Biostatistics, School of Public Health, Imperial College London which has additional infrastructure support provided by the NIHR Imperial Biomedical Research Centre (BRC). The national cohorts are supported by: Danish Cancer Society (Denmark);Ligue Nationale Contre le Cancer, Institut Gustave Roussy, Mutuelle Générale de l’Education Nationale (MGEN), Institut National de la Santé et de la Recherche Médicale (INSERM), French National Research Agency (ANR, reference ANR-10-COHO-0006), French Ministry for Higher Education (subsidy 2102918823, 2103236497, and 2103586016) (France); German Cancer Aid, German Cancer Research Center (DKFZ), German Institute of Human Nutrition Potsdam-Rehbruecke (DIfE), Federal Ministry of Research, Technology and Space (BMFTR) (Germany); Associazione Italiana per la Ricerca sul Cancro-AIRC-Italy, Italian Ministry of Health, Italian Ministry of University and Research (MUR), Compagnia di San Paolo (Italy); Dutch Ministry of Public Health, Welfare and Sports (VWS), the Netherlands Organisation for Health Research and Development (ZonMW), World Cancer Research Fund (WCRF), (The Netherlands); UiT The Arctic University of Norway; Health Research Fund (FIS) - Instituto de Salud Carlos III (ISCIII), Regional Governments of Andalucía, Asturias, Basque Country, Murcia and Navarra, and the Catalan Institute of Oncology - ICO (Spain); Swedish Cancer Society, Swedish Research Council of Region Skåne and Region Västerbotten (Sweden); Cancer Research UK (C864/A14136 to EPIC-Norfolk; C8221/A29017 to EPIC-Oxford), Medical Research Council (MR/N003284/1, MC-UU_12015/1 and MC_UU_00006/1 to EPIC-Norfolk (DOI 10.22025/2019.10.105.00004); MR/Y013662/1 to EPIC-Oxford) (United Kingdom). Previous support has come from “Europe against Cancer” Programme of the European Commission (DGSANCO).

The Atherosclerosis Risk in Communities study has been funded in whole or in part with Federal funds from the National Heart, Lung, and Blood Institute, National Institutes of Health, Department of Health and Human Services, under Contract nos. (75N92022D00001, 75N92022D00002, 75N92022D00003, 75N92022D00004, 75N92022D00005). Studies on cancer in ARIC are also supported by the National Cancer Institute (U01 CA164975). SomaLogic Inc. conducted the SomaScan® assays in exchange for use of ARIC data. This work was also supported in part by NIH/NHLBI grant R01 HL134320. The content of this work is solely the responsibility of the authors and does not necessarily represent the official views of the National Institutes of Health.

## 12. Conflict of interest

U Peters was a consultant with AbbVie and her family was or is holding individual stocks for the following companies: Alphabet Inc CL C, Amazon Com Inc, Aptera Motors Corp, ARM Holdings PLC, BioNTech, Boeing Company, BYD Company Limited, Crowdstrike Holdings Inc, Curvac BV, Microsoft Corp, MicroStrategy Inc, NVIDIA Corp, Stellantis, Tesla Inc, Walt Disney CO

## 13. Disclaimer

Where authors are identified as personnel of the International Agency for Research on Cancer/World Health Organization, the authors alone are responsible for the views expressed in this article and they do not necessarily represent the decisions, policy, or views of the International Agency for Research on Cancer/World Health Organization. This article is the result of the scientific work of Dr Murphy while he was affiliated at the International Agency for Research on Cancer.

