## Supplementary Material for "Proteomic markers of colorectal cancer risk: results from the European Prospective Investigation into Cancer and Nutrition"

In EPIC: “sex” was self-reported; “centre” was reported as the centre where participants were recruited; “date of blood draw” was modelled using a spherical transformation with four parameters such that the "effect" at time X is expected to be similar to the effect at time Y; “fasting status” was self-reported and coded as: *no*, *in between*, *yes*, *missing;* “body mass index” was measured as weight in kg divided by height in m^2^; “alcohol consumption” was self-reported as grams per day; “smoking” was self-reported as any one of: *never*, *former*, *current*, *unknown*; “physical activity” was self-reported using the Cambridge physical activity index as any one of: *inactive*, *moderately inactive*, *moderately active*, *active*, *missing*; “education” was self-reported as any one of: *none*, *primary school completed*, *secondary school*, *technical/professional school*, *longer education (incl. University degree)*, *not-specified*. Of the 977 colorectal cancers, 951 (97.3%) were adenocarcinomas, including mucinous and signet ring cell variants; the remaining 26 were carcinomas not otherwise specified (n = 15), signet ring cell carcinoma (n = 9), undifferentiated carcinoma (n = 1), and cribriform carcinoma (n = 1). Separate aptamers can bind to isoforms of the same protein but can also bind to the same protein at different sites, which can be impacted by post-translational modifications or complexes formed with other proteins. The SomaScan 7K assay therefore measured 7,289 aptamers targeting 6,402 unique proteins.

### 2. Atherosclerosis Risk in Communities study

Relative fluorescence units were log2 transformed and, to account for non-genetic variance, the PEER method was used to estimate a set of latent covariates. Measurements were then adjusted in a linear regression model including PEER factors, study site, and the first 10 genetic principal components; residuals were inverse-rank transformed and used as the corrected protein value. In ARIC: “sex” was self-reported; “race” was self-reported; “study centre” was reported as the centre where participants were recruited; proteomic PEER factors were calculated using R package MOFA2; genetic principal components were calculated using PLINK (version 2.0) – genotyping in ARIC was performed using the Affymetrix Genome-Wide Human SNP Array 6.0, with imputation based on the TOPMed reference panel, and genetic PCs were calculated by projecting onto the PC axes derived from the 1000 Genomes + HGDP Project; “body mass index” was measured by trained staff as weight in kg divided by height in m^2^; “waist hip ratio” was derived from girth of waist in cm divided by girth of hip in cm; “diabetes status” was modelled as *diagnosed*, *undiagnosed*, *at risk*, or *normoglycemic* and defined as self-reported physician diagnosis (yes, no) or use of diabetes medication; and among participants without a diabetes diagnosis, undiagnosed diabetes (yes, no) was defined as fasting glucose ≥126 mg/dL (non-fasting ≥200 mg/dL) or HbA1c ≥6.5%; at risk for diabetes (yes, no) was defined as fasting glucose 100-125 mg/dL (non-fasting 140-199 mg/dL) or HbA1c 5.7-6.4%; and otherwise participants were considered to be normoglycemic (reference); “smoking status” was self-reported as *never (reference)*, *former*, or *current*; packyears was calculated from detailed self-reported information on years of smoking and usual number of cigarettes smoked; and “physical activity” was measured by work index, sport index and leisure time index^1^.

### 3. UK Biobank

In UK Biobank: “sex” was self-reported; “centre” was the assessment centre at recruitment; “body mass index” was measured as weight in kg divided by height in m2; “alcohol consumption” was self-reported as never, special occasion only, one to three times a month, once or twice a week, three or four times a week, or daily or almost daily; “smoking status” was self-reported as never, previous, or current; “physical activity” was self-reported using the International Physical Activity Questionnaire and categorised as low, moderate, or high; “education” was self-reported and categorised as primary school or below, secondary school, technical/professional school, or university degree or higher; and “Townsend deprivation index” was derived from the postcode of residence at recruitment and categorised into tertiles. Cox proportional hazard models were stratified by sex, assessment centre, and 5-year age group, and adjusted for age at recruitment, smoking status, alcohol consumption, education, physical activity, Townsend deprivation index, and body mass index.

### 4. Mendelian randomisation and genetic colocalisation analyses

We obtained genome-wide summary-level data for the association between genetic variants and Somalogic derived proteomic data from Ferkingstad et al., (2021)^2^, Pietzner et al., (2021)^3^, and Zhang et al., (2022)^4^, and for colorectal cancer from Huyghe et al., (2019)^5^. Briefly, data from Ferkingstad et al., included up-to 35,287 individuals from an Icelandic ancestral population; data from Pietzner et al., included up-to 10,708 individuals from a European ancestral population; data from Zhang et al., included up-to 7,213 individuals from a European American ancestral population; colorectal cancer data from Huyghe et al., of up-to 125,478 individuals of European and East Asian ancestral populations has previously been reported^6,7^ as physician diagnosed and we make the assumption from the supplementary data that cases are predominantly incident cases of unknow stage.

MR relies upon 3 core assumptions: (1) the instrument is associated with the exposure, (2) there are no confounders of the association between the instrument and the outcome, and (3) the instrument is not related to the outcome except via its effect on the exposure^8^. Assumption 1 is generally satisfied by using a genome-wide significance threshold of 5 x 10^-8^. Assumptions 2 and 3 are unverifiable but evidence for a shared causal variant identified through genetic colocalisation can strengthen evidence from MR analyses. Enumeration colocalisation systematically appraises evidence for different hypotheses in a Bayesian framework and assumes that the causal variant is included in the analysis and that the underlying populations are the same. The hypotheses are: h0, neither trait has a causal variant in the region; h1, only trait 1 has a causal variant in the region; h2, only trait 2 has a causal variant in the region; h3, both traits are associated with distinct causal variants; h4, both traits are associated with a shared causal variant.

Assuming that a protein quantitative trait loci is generally within 1Mb (mega base) of the protein-coding gene^9^ we assume there are likely multiple causal variants within a 2Mb window around the protein coding gene (i.e., 1Mb up and down from the protein coding gene start and end position) and as such use a standard genome-wide significance p-value threshold of 5x10^-8^ and use a strict linkage disequilibrium R^2^ threshold of 0.001 for a 10kb block to identify genetic variants for MR analyses. We used the 1000 Genomes (phase 3)^10^ European ancestral population to identify independent genetic variants. Where genetic variants were not available in the colorectal cancer GWAS, genetic variants with an LD R^2^ ≥ 0.8 were used instead; the reference strand for ambiguous genetic variants was inferred using minor allele frequency, so long as the minor allele frequency was not ≥0.3 in which case the genetic variant was excluded; genetic variants for the exposure and outcome were harmonised such that the exposure effect allele was on the increasing scale.

In genetic colocalisation analyses, as the number of available genetic variants differed between protein-colorectal cancer pairs, we used the following thresholds to ensure the per-hypothesis priors (h0 = ~80%, h1=h2 = ~10%, h3 = ~1%, h4 = ~1%) remained consistent across different numbers of genetic variants: up-to 2,499 genetic variants p^1^=p^2^=10^-4^ p^12^=10^-5^; 2,500–7,499 genetic variants p^1^=p^2^=20^-5^ p^12^=20^-6^; ≥7,500 genetic variants: p^1^=p^2^=10^-5^ p^12^=10^-611^; where p^1^ is the probability that each variant is the causal variant for the protein; p^2^ is the probability that each variant is the causal variant for CRC; and p^12^ is the probability that each variant is the causal variant for both traits. We considered a threshold of h4 ≥ 0.8 as evidence of a shared causal signal.

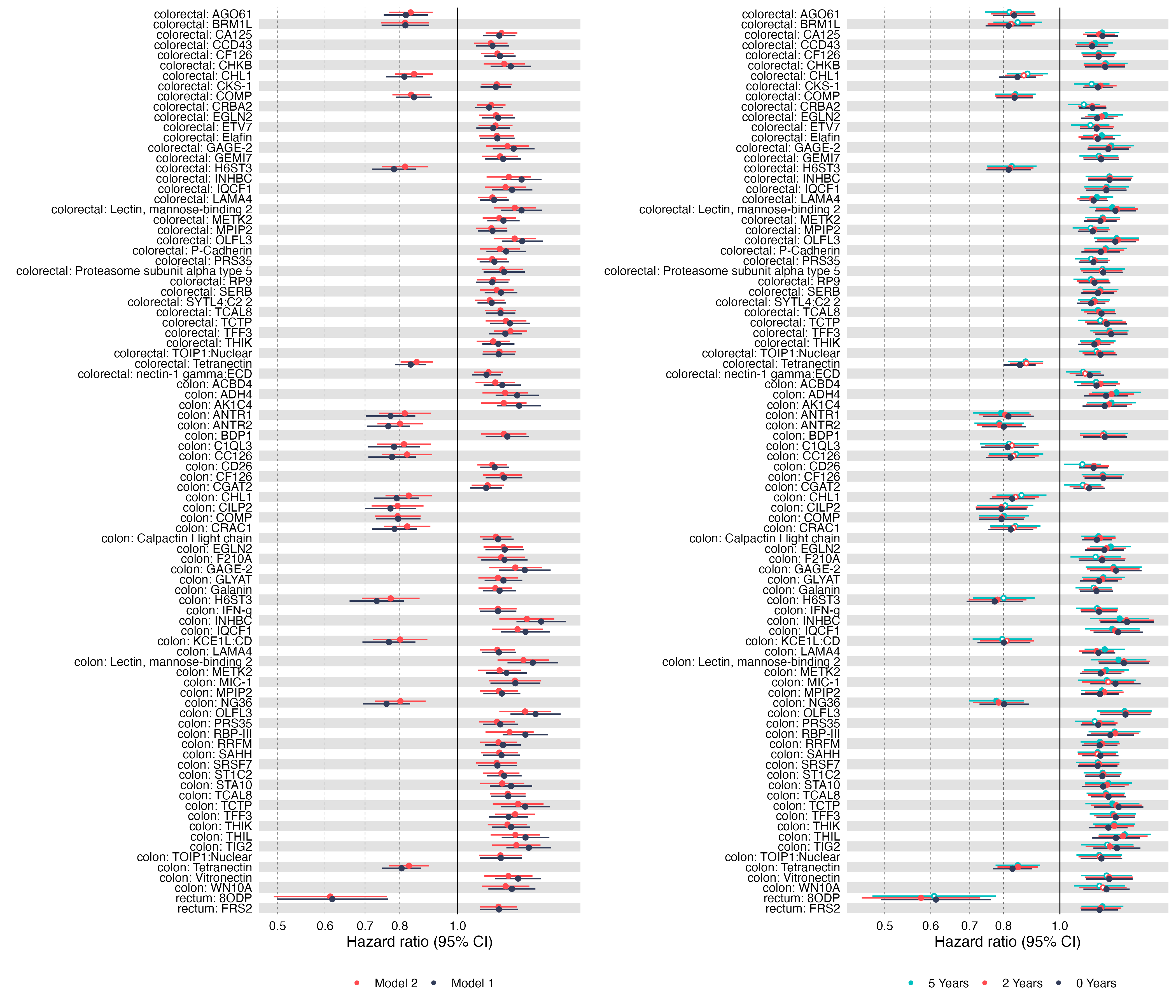

Supplementary Figure 1 Comparison of models and follow-up exclusions for proteins associated with colorectal, colon, and rectal cancer risk in the main analysis. Forest plots show the hazard ratio and 95% confidence interval for each protein reaching the false discovery rate threshold (FDR-adjusted p < 0.05) in the sex-combined analysis of colorectal (36 proteins), colon (50 proteins), or rectal (2 proteins) cancer. Proteins are grouped by cancer and labelled as cancer: protein. (Left) Comparison of the minimally-adjusted (model 1) and fully-adjusted (model 2) models, with no follow-up exclusion. (Right) Comparison of the fully-adjusted model (model 2) with no exclusion and after excluding the first 2- and 5-years of follow-up. Filled points indicate FDR-adjusted p < 0.05 in the analysis shown.

Supplementary Figure 2 Correlation heatmaps for proteins associated with cancer risk within each cancer-sex analysis. Supplementary Figure 2 is a multi-page PDF with one page for each cancer (colorectal, colon, rectum, proximal colon, distal colon) and sex (combined, female, male) combination. Each page shows 3 heatmaps of the pairwise Pearson correlations between the proteins reaching the false discovery rate threshold (FDR-adjusted p < 0.05) in that analysis, calculated among all participants, among cases only, and among non-cases only, using pairwise complete observations. Correlations are reported in Supplementary Table 3.

Supplementary Figure 3 Correlation networks for proteins associated with cancer risk within each cancer-sex analysis. Supplementary Figure 3 is a multi-page PDF with one page for each cancer (colorectal, colon, rectum, proximal colon, distal colon) and sex (combined, female, male) combination. Each page shows 3 network maps, calculated among all participants, among cases only, and among non-cases only, in which proteins are connected where the pairwise Pearson correlation exceeds 0.4; edge width and shading are scaled to the strength of the correlation.

Supplementary Figure 4 Comparison of models and follow-up exclusions for proteins associated with cancer risk within each cancer-sex analysis. Supplementary Figure 4 is a multi-page PDF with one page for each cancer (colorectal, colon, rectum, proximal colon, distal colon) and sex (combined, female, male) combination. Each page shows 2 forest plots for the proteins reaching the false discovery rate threshold (FDR-adjusted p < 0.05) in that analysis: (left) a comparison of the fully-adjusted model (model 2) with no exclusion and after excluding the first 2- and 5-years of follow-up, and (right) a comparison of the minimally-adjusted (model 1) and fully-adjusted (model 2) models with no follow-up exclusion. Filled points indicate FDR-adjusted p < 0.05. Combinations with no proteins reaching the threshold are shown as an empty page.

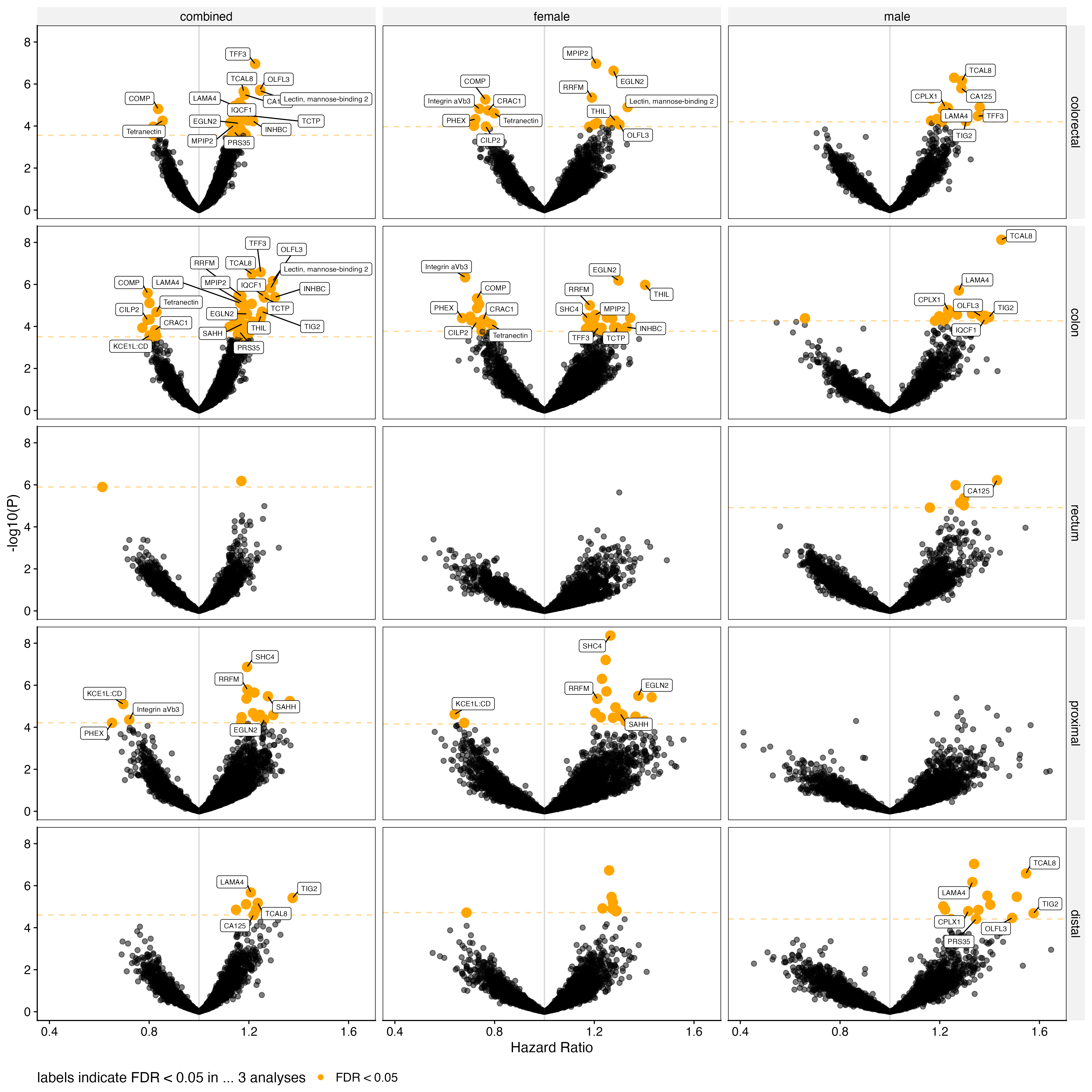

Supplementary Figure 5 Cox proportional hazard models of the association between 7,289 aptamers, targeting 6,402 unique proteins, and all cancer-sex combinations. The volcano plot shows the hazard ratio and -log10 p-value for each aptamer. Highlighted (orange) points reached the Benjamini-Hochberg false discovery rate p-value threshold (5%) as indicated by the horizontal line. Labelled points are proteins associated (FDR) in ≥ 3 cancer-sex analyses.

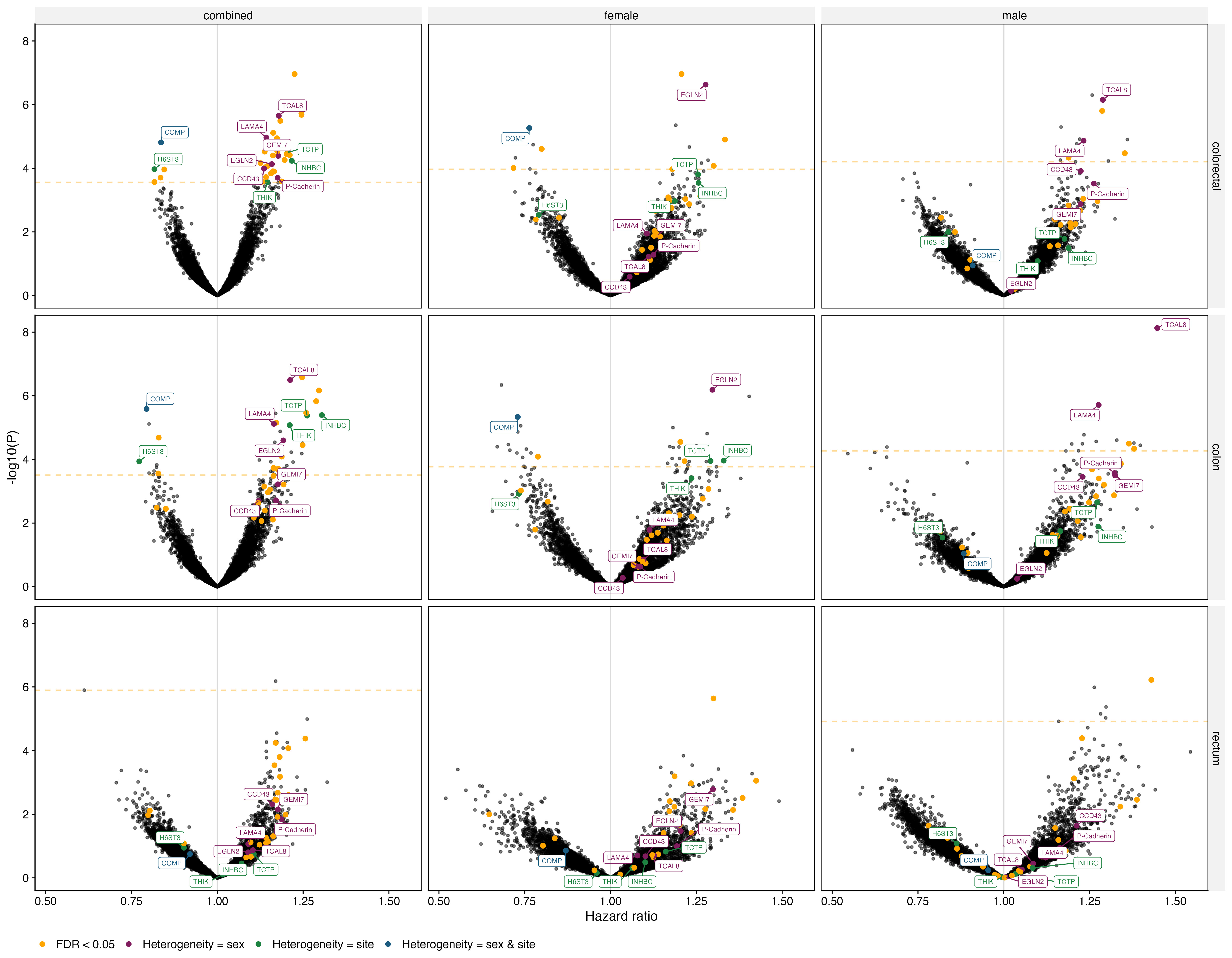

Supplementary Figure 6 Evidence of heterogeneity by sex and/or by site in Cox proportional hazard models of colorectal cancer. All panels show the results from the respective sex-cancer analysis and orange points in all panels highlight the 36 proteins associated in the sex-combined colorectal cancer analysis. Purple points highlight proteins where there was evidence of heterogeneity by sex; green points highlight proteins where there was evidence of heterogeneity by site; blue points highlight proteins where there was evidence of heterogeneity by sex and site; the horizontal line (orange) indicates the false discovery rate (FDR) p-value threshold for the respective analysis.

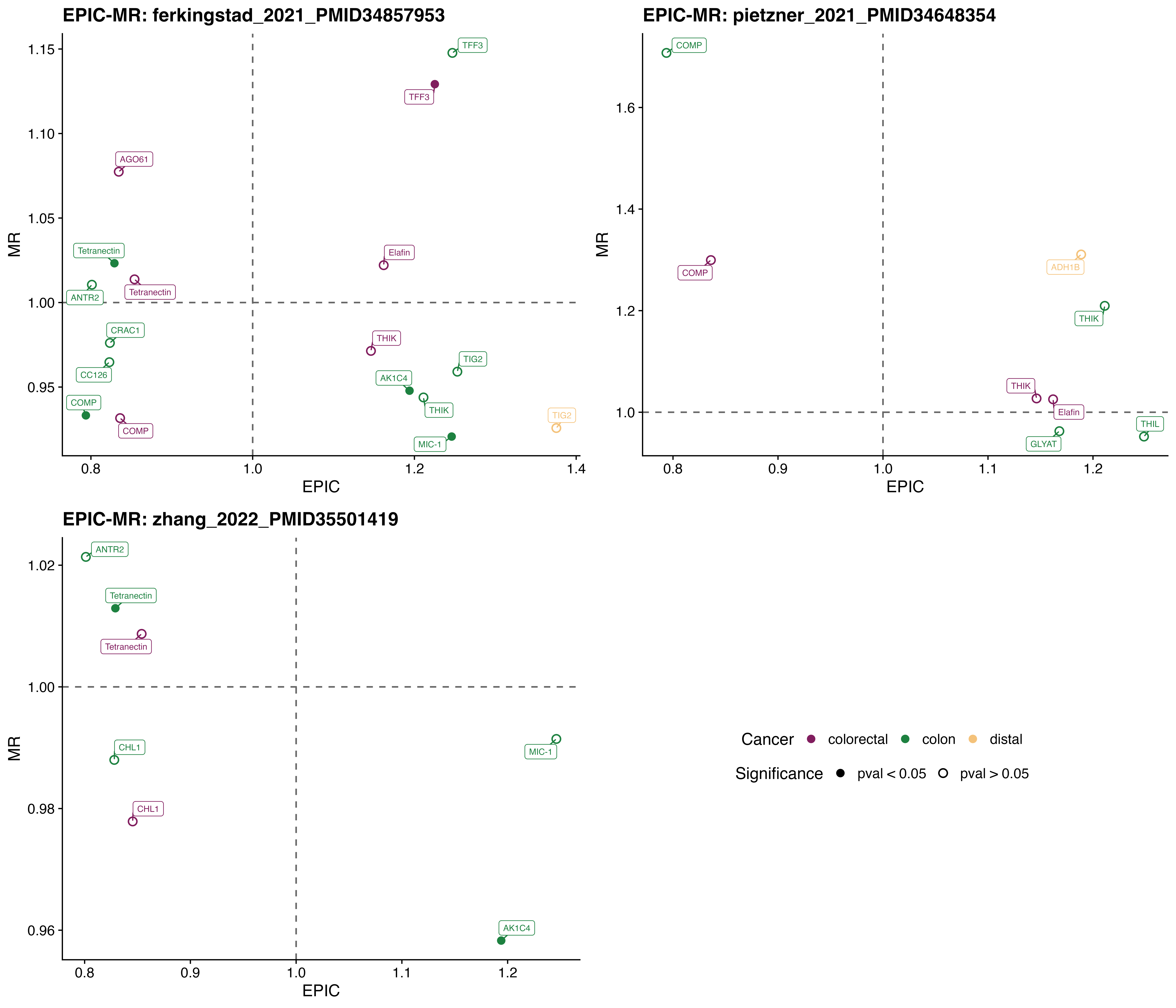

Supplementary Figure 7 Comparison of colorectal cancer associated proteins in EPIC with results from MR analyses using different genome-wide summary-level datasets to identify protein instruments. Scatter plots show the hazard ratio (EPIC) and odds ratios (MR) for proteins associated with colorectal, colon, rectal, proximal colon, and distal colon cancer, with 1 panel per protein-QTL dataset (Ferkingstad et al., 2021; Pietzner et al., 2021; Zhang et al., 2022). Solid points indicate a nominal p-value threshold (0.05) was reached in the MR analysis.
