## Supplementary Figure 2 for "Proteomic markers of colorectal cancer risk: results from the European Prospective Investigation into Cancer and Nutrition"

colorectal / combined − all (N=5057)

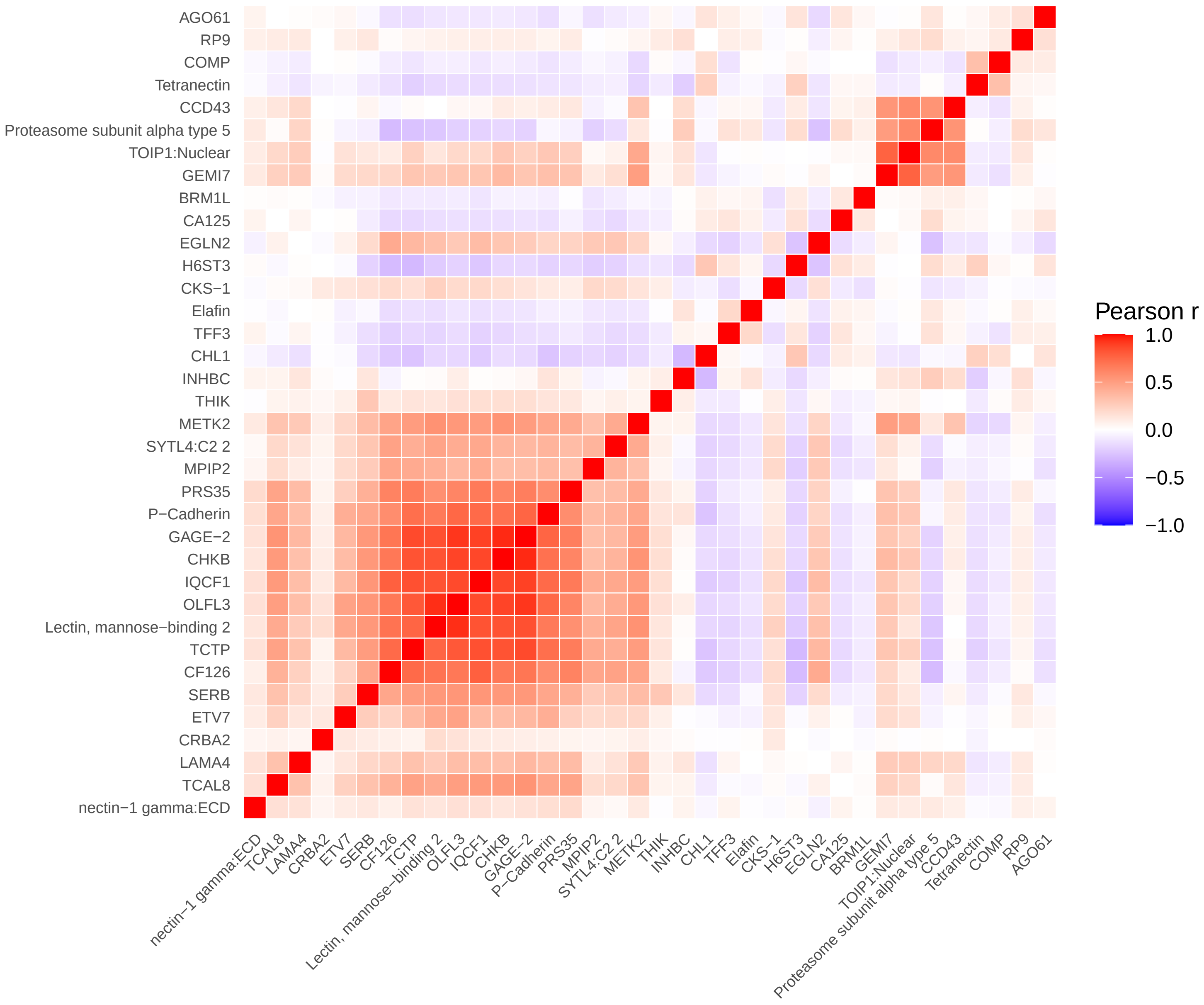


colorectal / combined − cases (N=977)

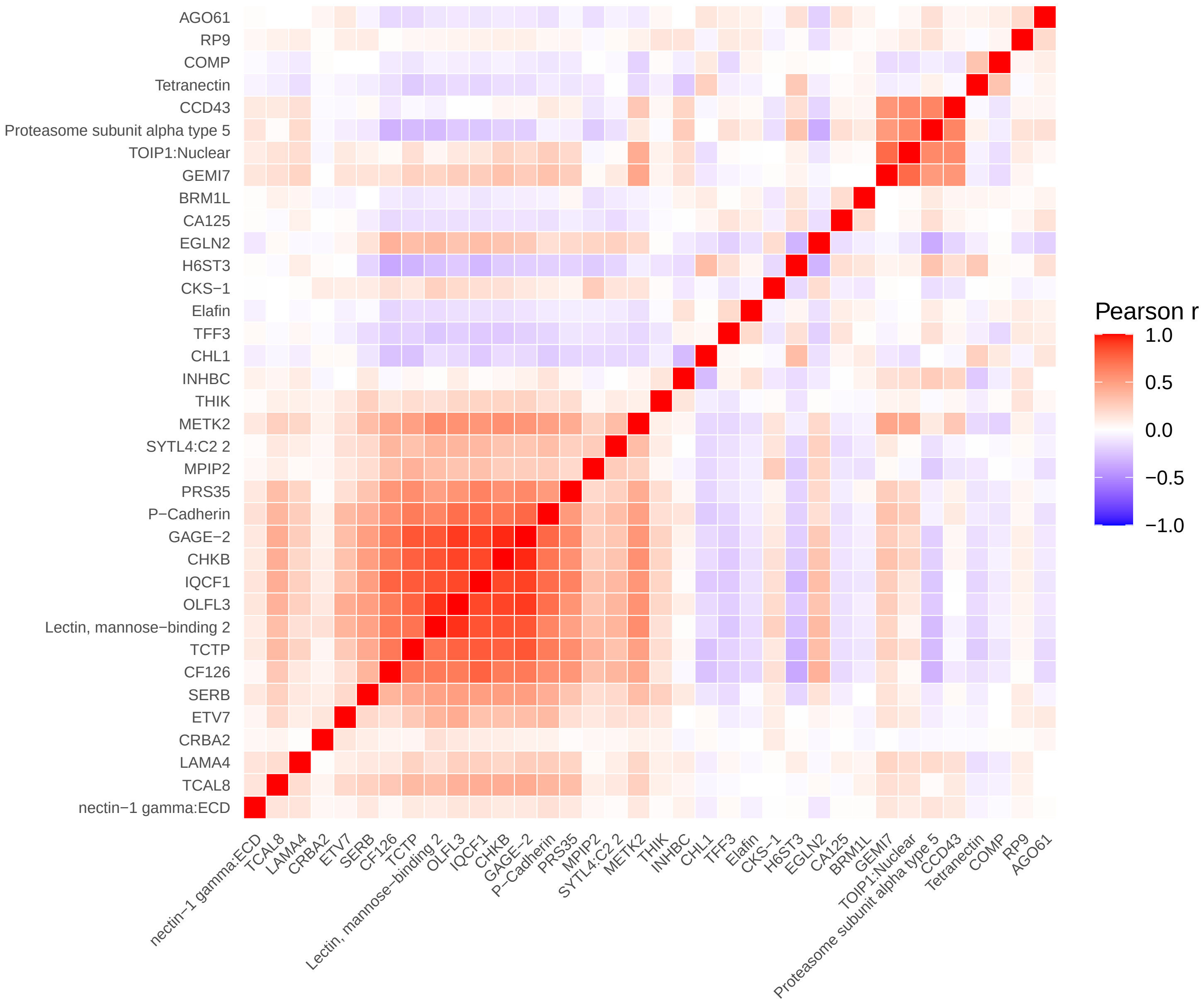


colorectal / combined − non−cases (N=4080)

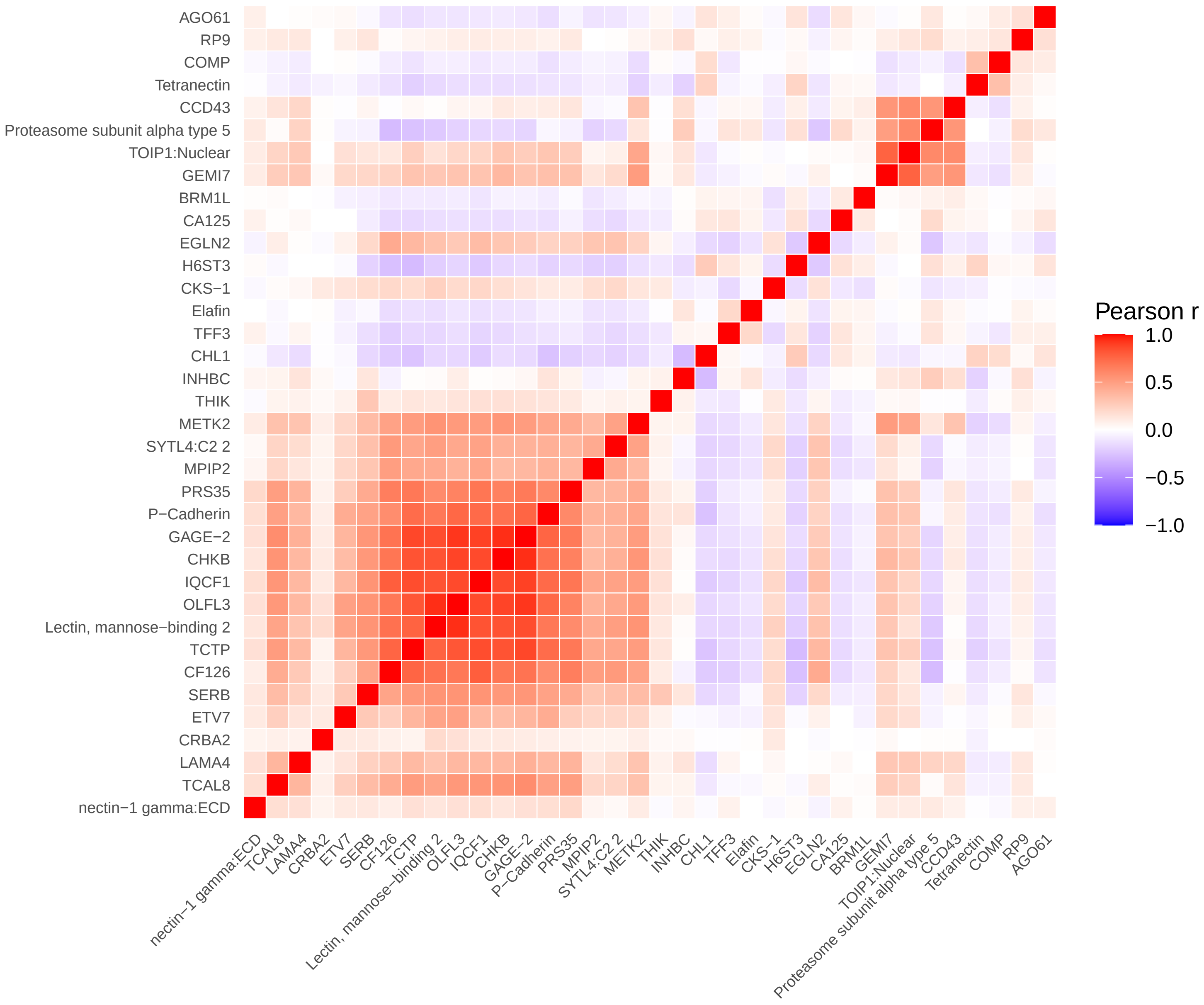

colorectal / female − all (N=3046)

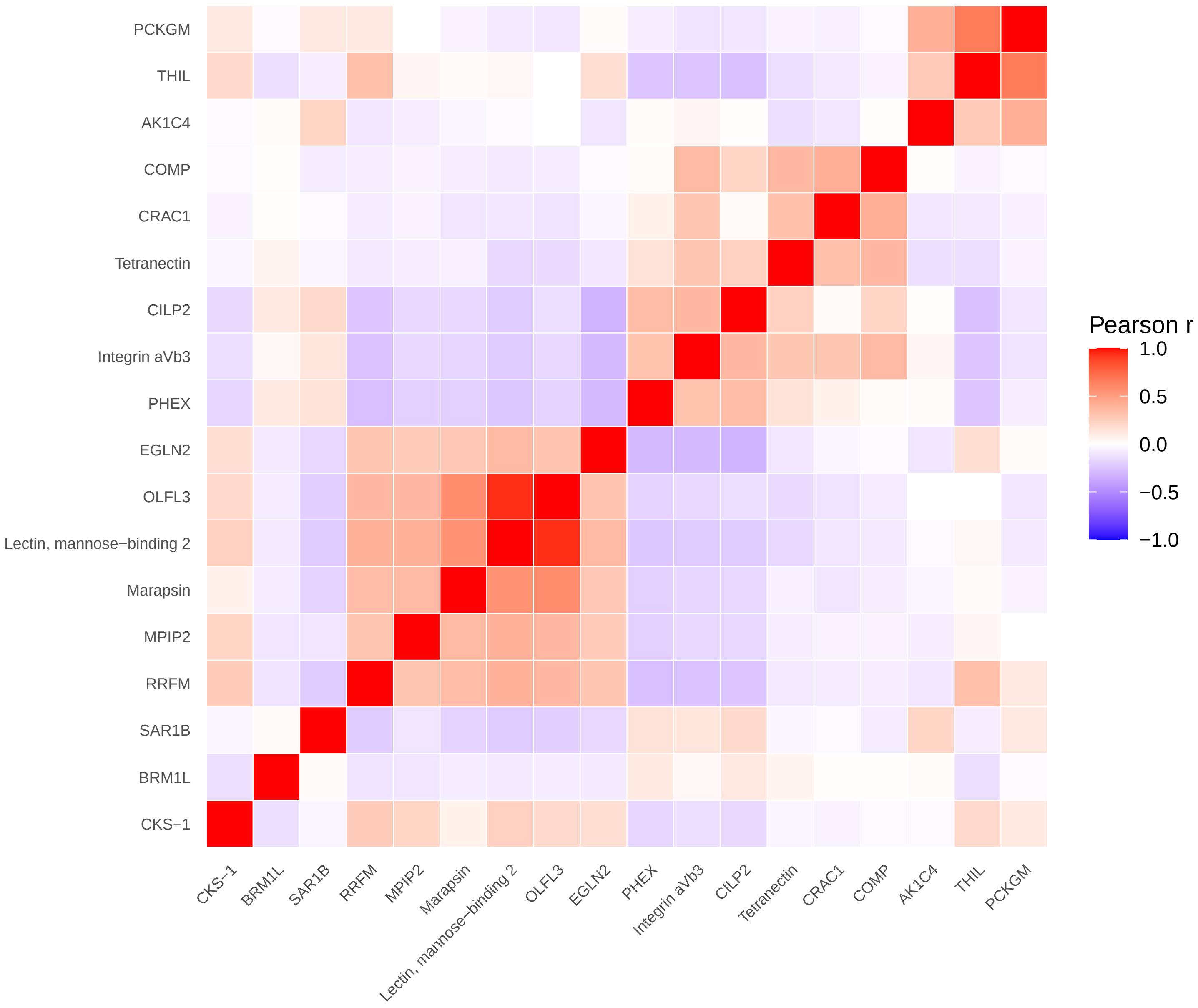


colorectal / female − cases (N=519)

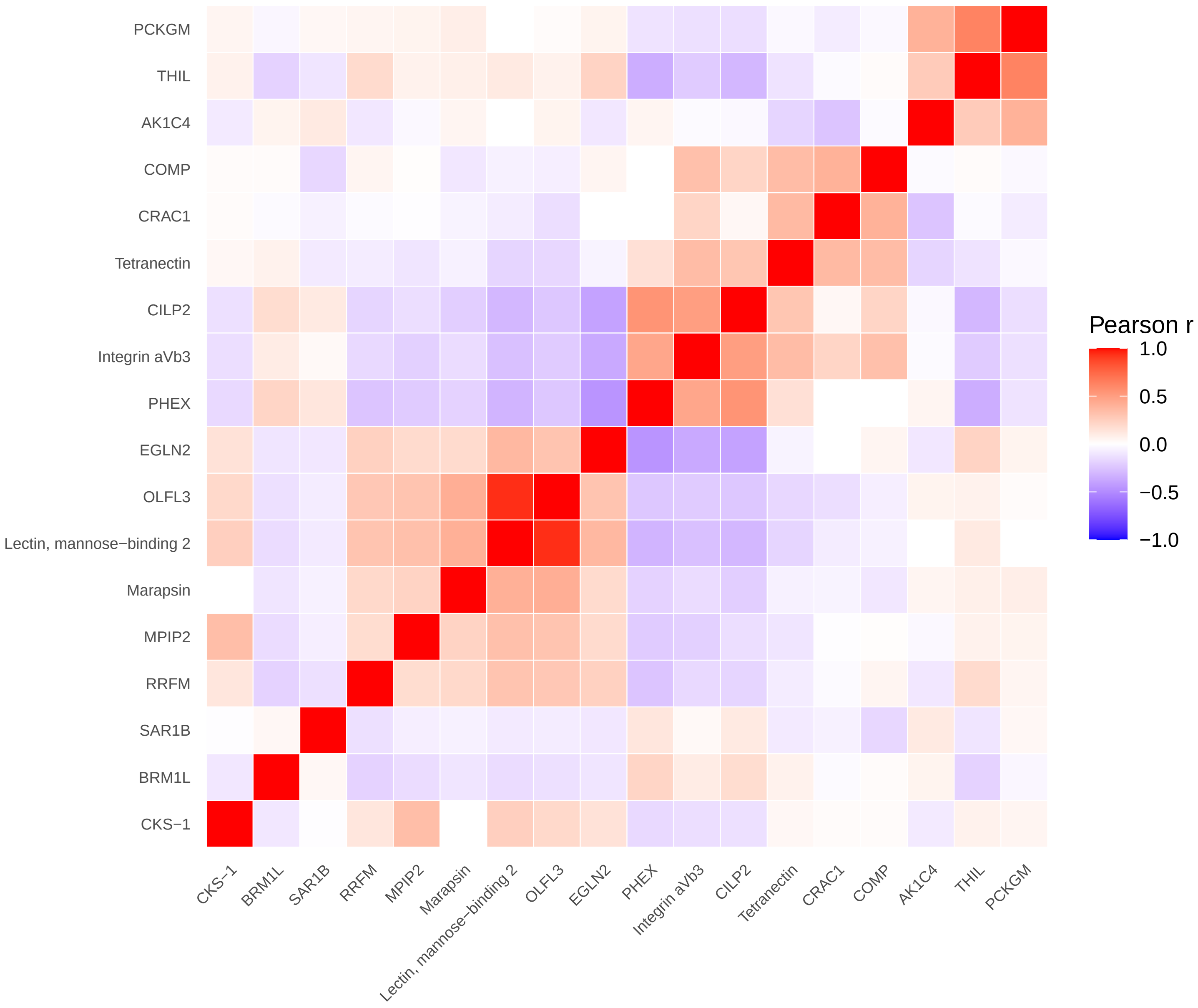


colorectal / female − non−cases (N=2527)

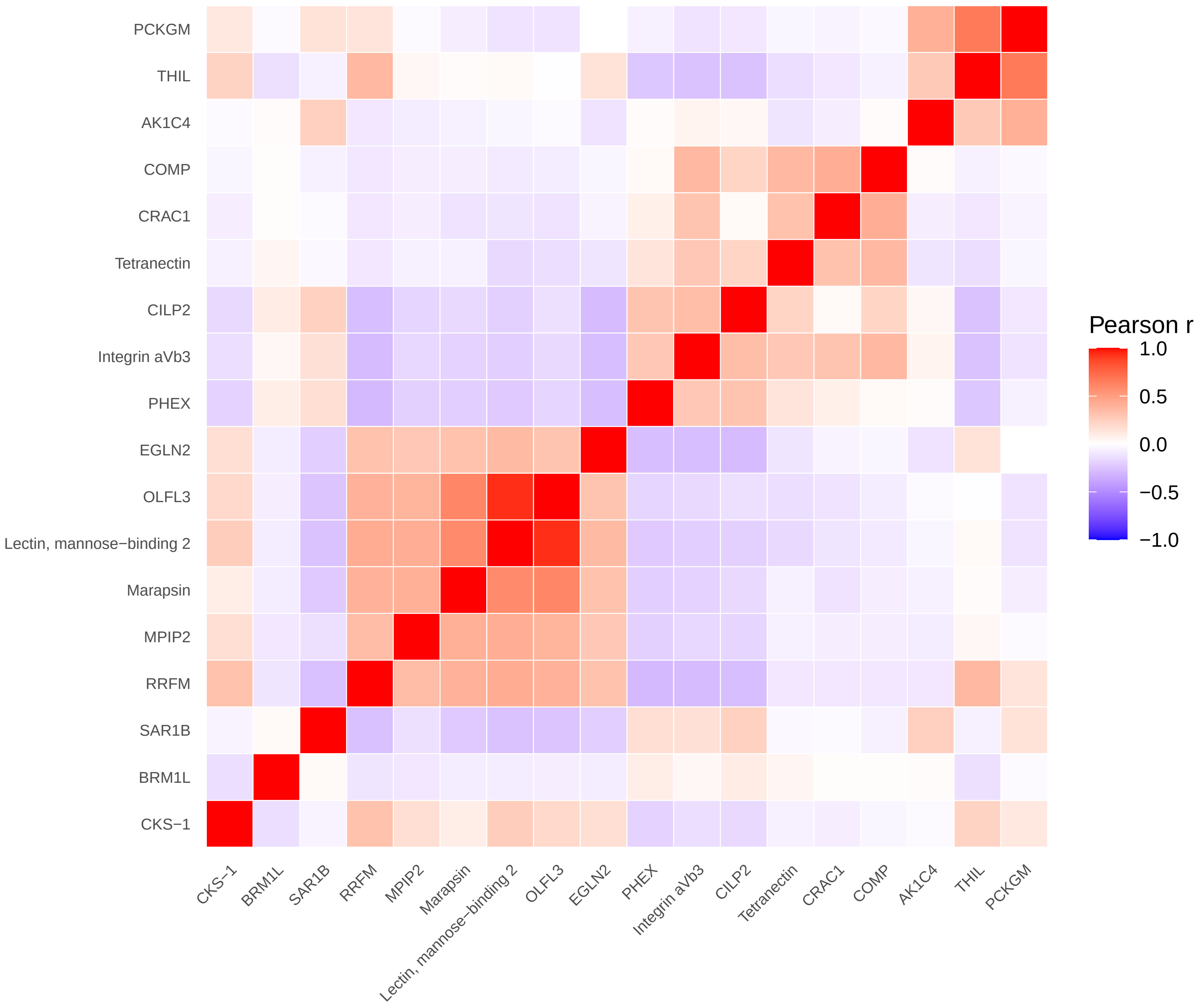

colorectal / male − all (N=2011)

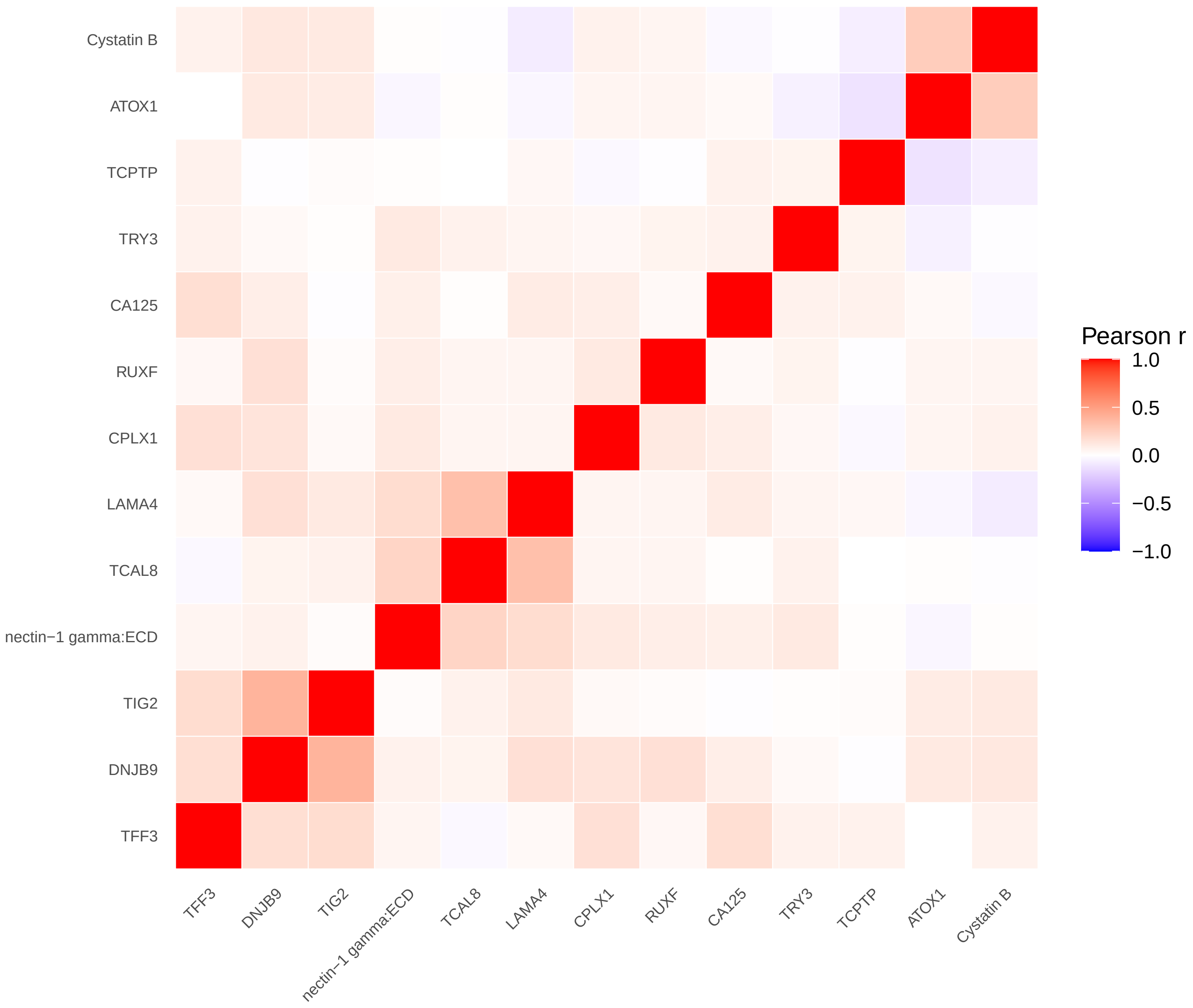


colorectal / male − cases (N=458)

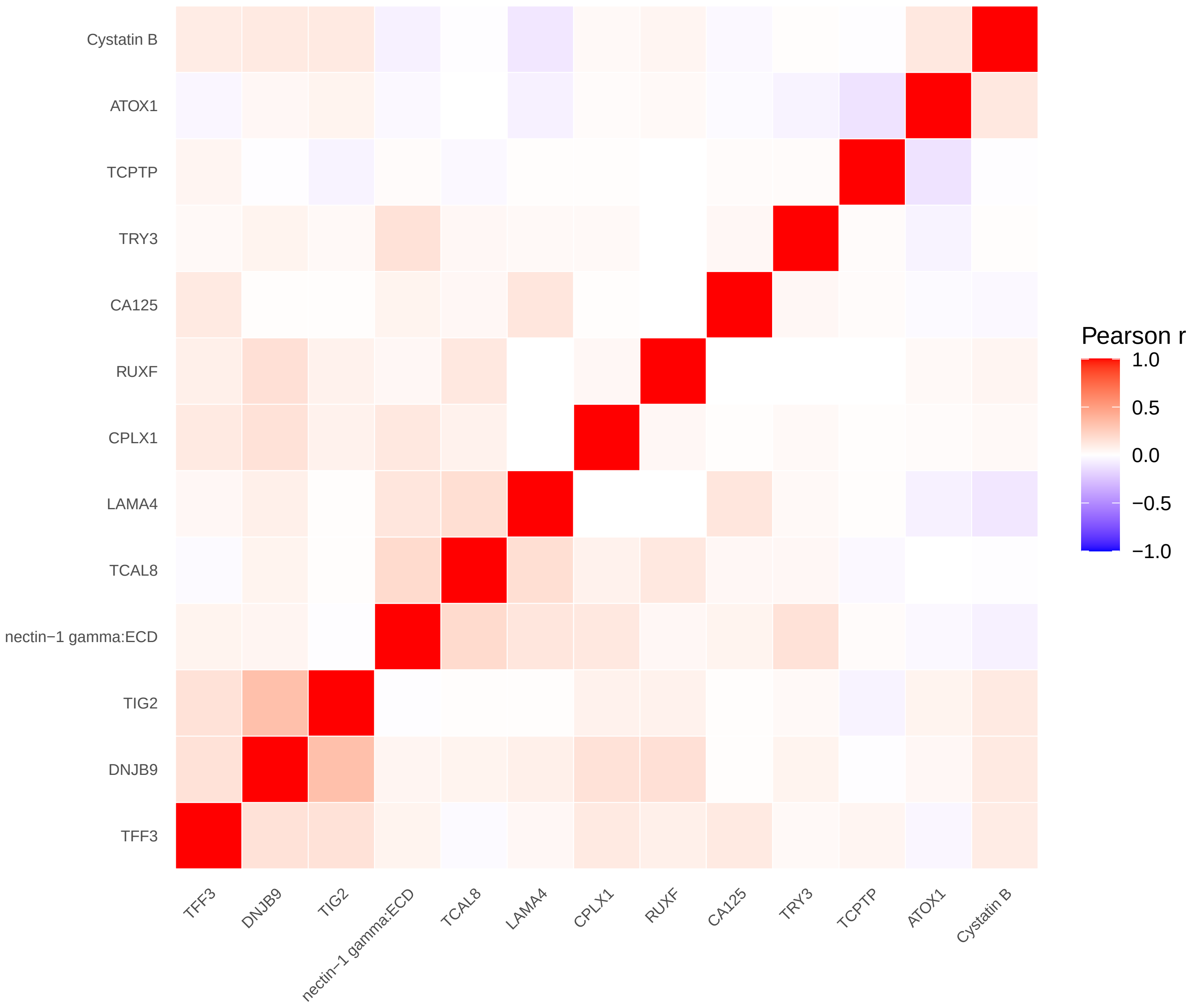


colorectal / male − non−cases (N=1553)

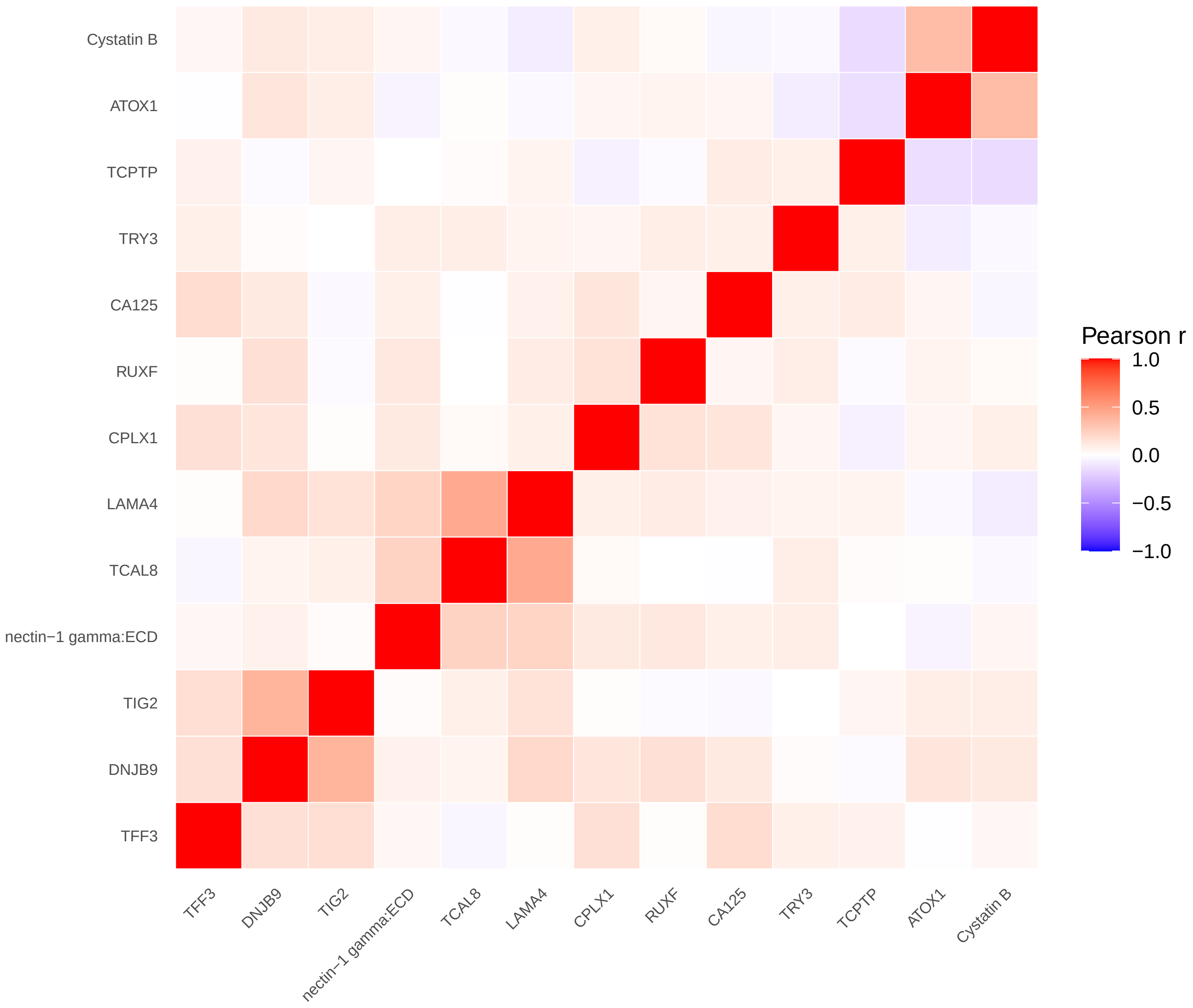

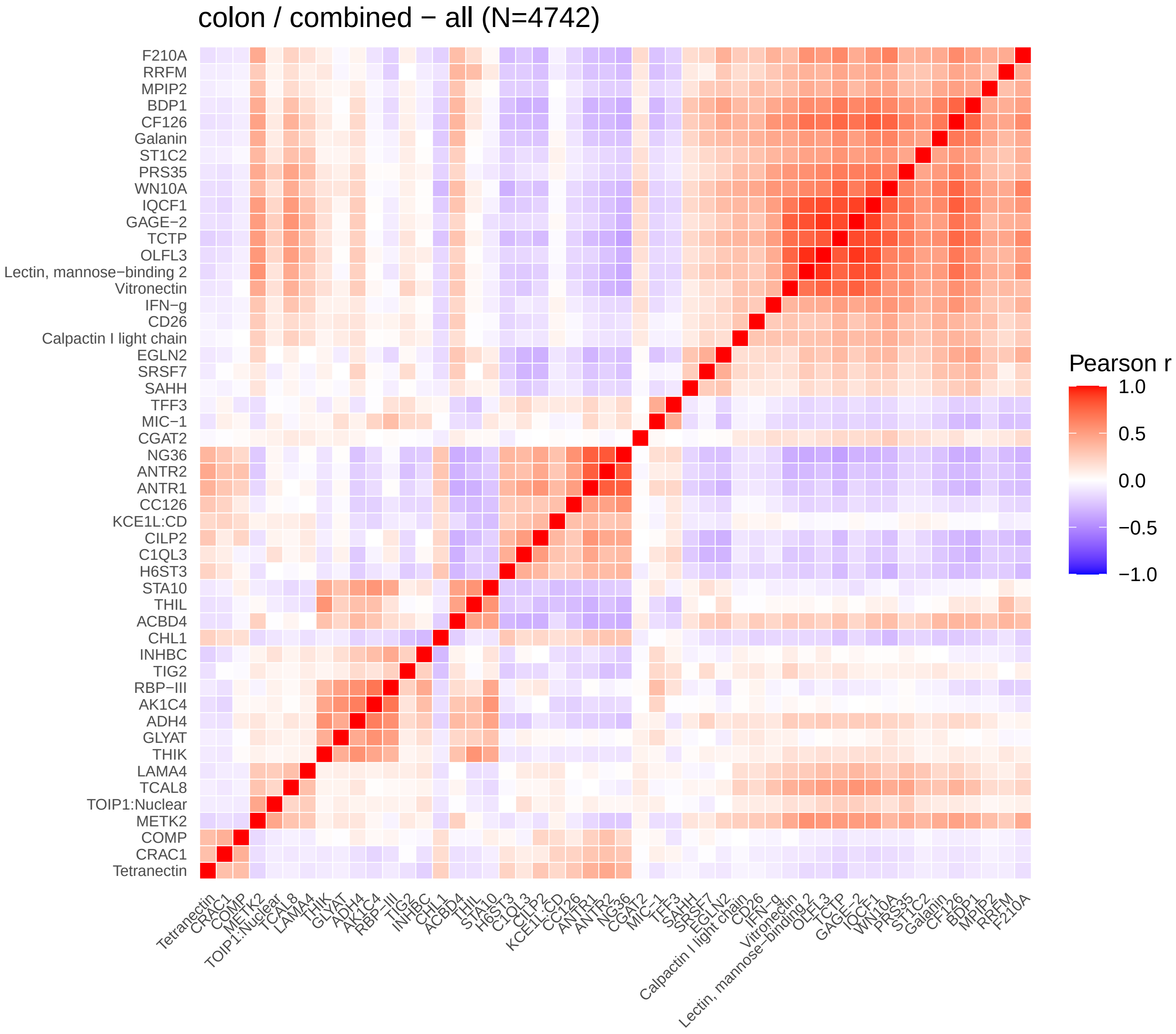
colon / combined − all (N=4742)
F210A
RRFM
MPIP2
BDP1
CF126
Galanin
ST1C2
PRS35
WN10A
IQCF1
GAGE−2
TCTP
OLFL3
Lectin, mannose−binding 2
Vitronectin
IFN−g
CD26
Calpactin I light chain
EGLN2
SRSF7
SAHH
TFF3
MIC−1
CGAT2
NG36
ANTR2
ANTR1
CC126
KCE1L:CD
CILP2
C1QL3
H6ST3
STA10
THIL
ACBD4
CHL1
INHBC
TIG2
RBP−III
AK1C4
ADH4
GLYAT
THIK
LAMA4
TCAL8
TOIP1:Nuclear
METK2
COMP
CRAC1
Tetranectin
Pearson r
1.0
0.5
0.0
−0.5
−1.0

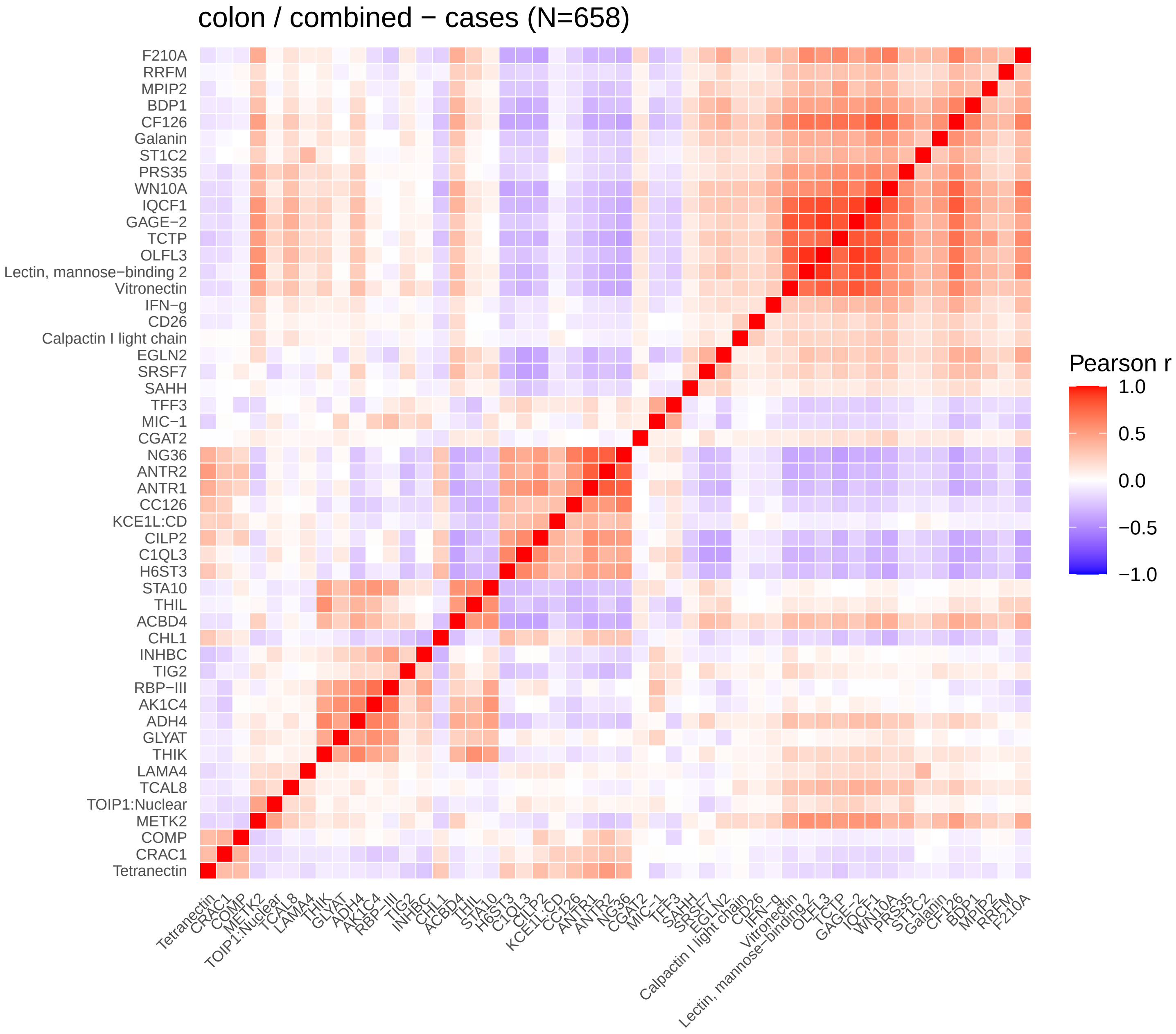
colon / combined − cases (N=658)
F210A
RRFM
MPIP2
BDP1
CF126
Galanin
ST1C2
PRS35
WN10A
IQCF1
GAGE−2
TCTP
OLFL3
Lectin, mannose−binding 2
Vitronectin
IFN−g
CD26
Calpactin I light chain
EGLN2
SRSF7
SAHH
TFF3
MIC−1
CGAT2
NG36
ANTR2
ANTR1
CC126
KCE1L:CD
CILP2
C1QL3
H6ST3
STA10
THIL
ACBD4
CHL1
INHBC
TIG2
RBP−III
AK1C4
ADH4
GLYAT
THIK
LAMA4
TCAL8
TOIP1:Nuclear
METK2
COMP
CRAC1
Tetranectin
Pearson r
1.0
0.5
0.0
−0.5
−1.0

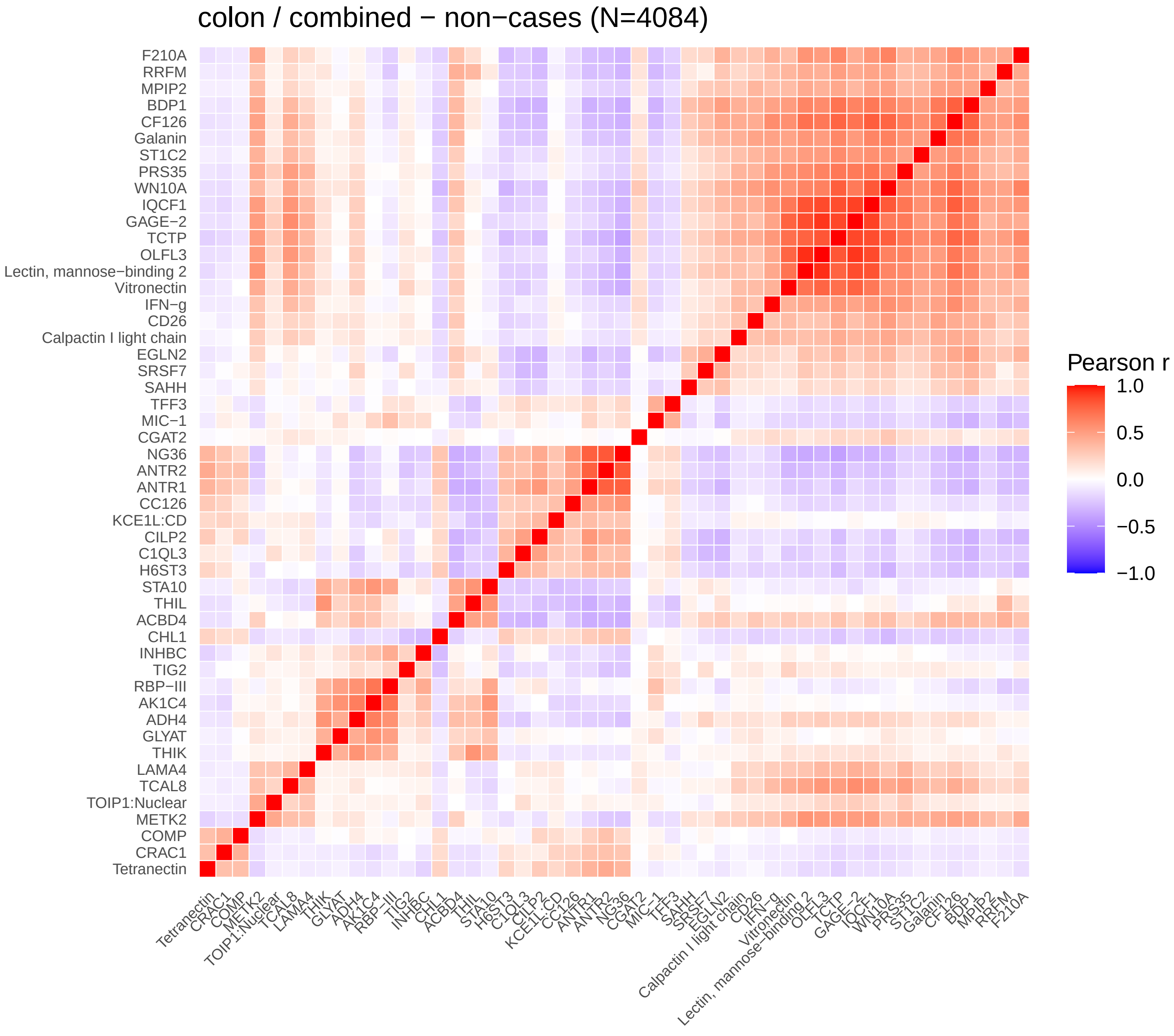
colon / combined − non−cases (N=4084)
F210A
RRFM
MPIP2
BDP1
CF126
Galanin
ST1C2
PRS35
WN10A
IQCF1
GAGE−2
TCTP
OLFL3
Lectin, mannose−binding 2
Vitronectin
IFN−g
CD26
Calpactin I light chain
EGLN2
SRSF7
SAHH
TFF3
MIC−1
CGAT2
NG36
ANTR2
ANTR1
CC126
KCE1L:CD
CILP2
C1QL3
H6ST3
STA10
THIL
ACBD4
CHL1
INHBC
TIG2
RBP−III
AK1C4
ADH4
GLYAT
THIK
LAMA4
TCAL8
TOIP1:Nuclear
METK2
COMP
CRAC1
Tetranectin
Pearson r
1.0
0.5
0.0
−0.5
−1.0

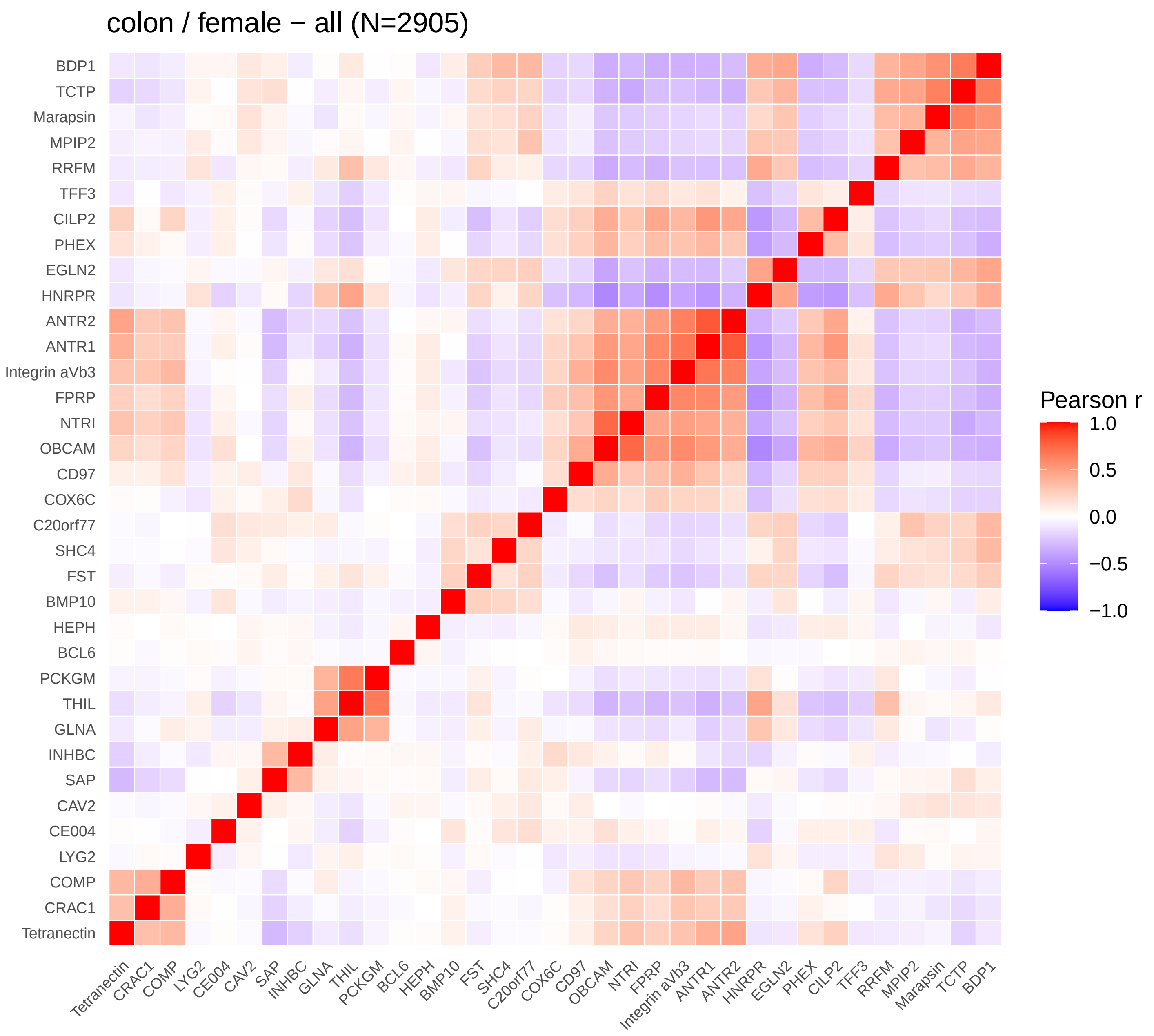
colon / female − all (N=2905)
BDP1
TCTP
Marapsin
MPIP2
RRFM
TFF3
CILP2
PHEX
EGLN2
HNRPR
ANTR2
ANTR1
Integrin aVb3
FPRP
NTRI
OBCAM
CD97
COX6C
C20orf77
SHC4
FST
BMP10
HEPH
BCL6
PCKGM
THIL
GLNA
INHBC
SAP
CAV2
CE004
LYG2
COMP
CRAC1
Tetranectin
Pearson r
1.0
0.5
0.0
−0.5
−1.0

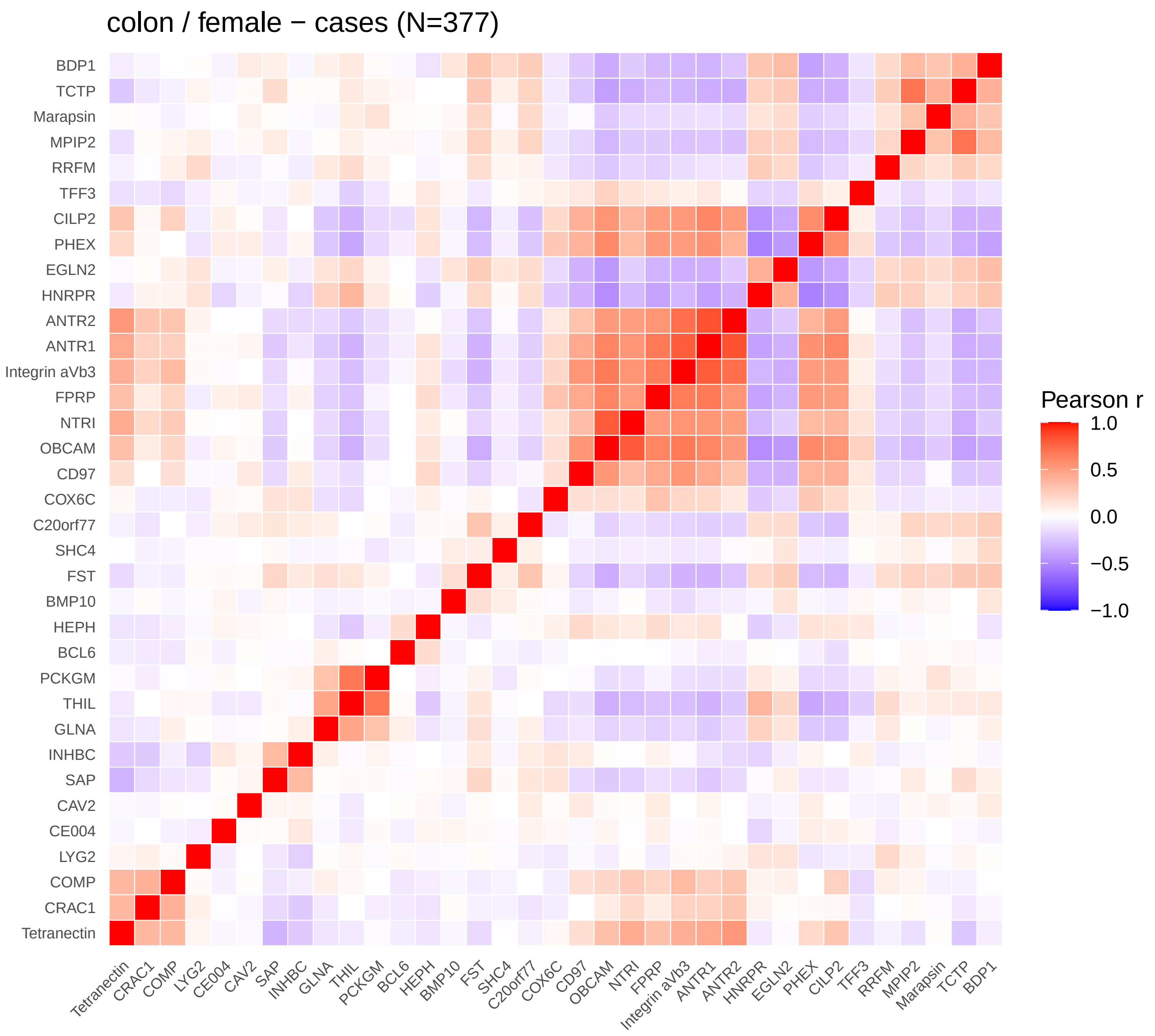
colon / female − cases (N=377)
BDP1
TCTP
Marapsin
MPIP2
RRFM
TFF3
CILP2
PHEX
EGLN2
HNRPR
ANTR2
ANTR1
Integrin aVb3
FPRP
NTRI
OBCAM
CD97
COX6C
C20orf77
SHC4
FST
BMP10
HEPH
BCL6
PCKGM
THIL
GLNA
INHBC
SAP
CAV2
CE004
LYG2
COMP
CRAC1
Tetranectin
Pearson r
1.0
0.5
0.0
−0.5
−1.0

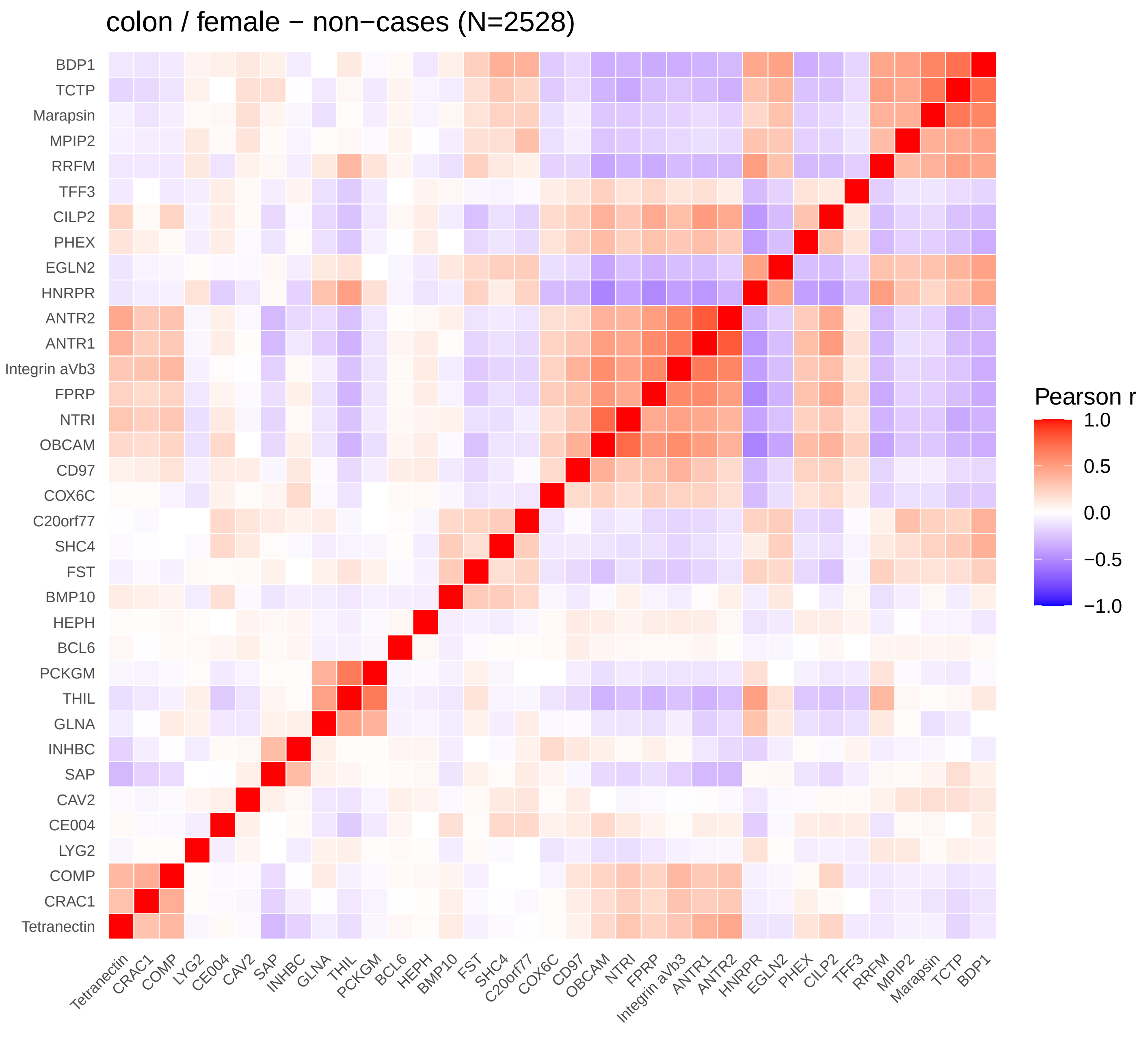
colon / female − non−cases (N=2528)
BDP1
TCTP
Marapsin
MPIP2
RRFM
TFF3
CILP2
PHEX
EGLN2
HNRPR
ANTR2
ANTR1
Integrin aVb3
FPRP
NTRI
OBCAM
CD97
COX6C
C20orf77
SHC4
FST
BMP10
HEPH
BCL6
PCKGM
THIL
GLNA
INHBC
SAP
CAV2
CE004
LYG2
COMP
CRAC1
Tetranectin
Pearson r
1.0
0.5
0.0
−0.5
−1.0

colon / male − all (N=1837)

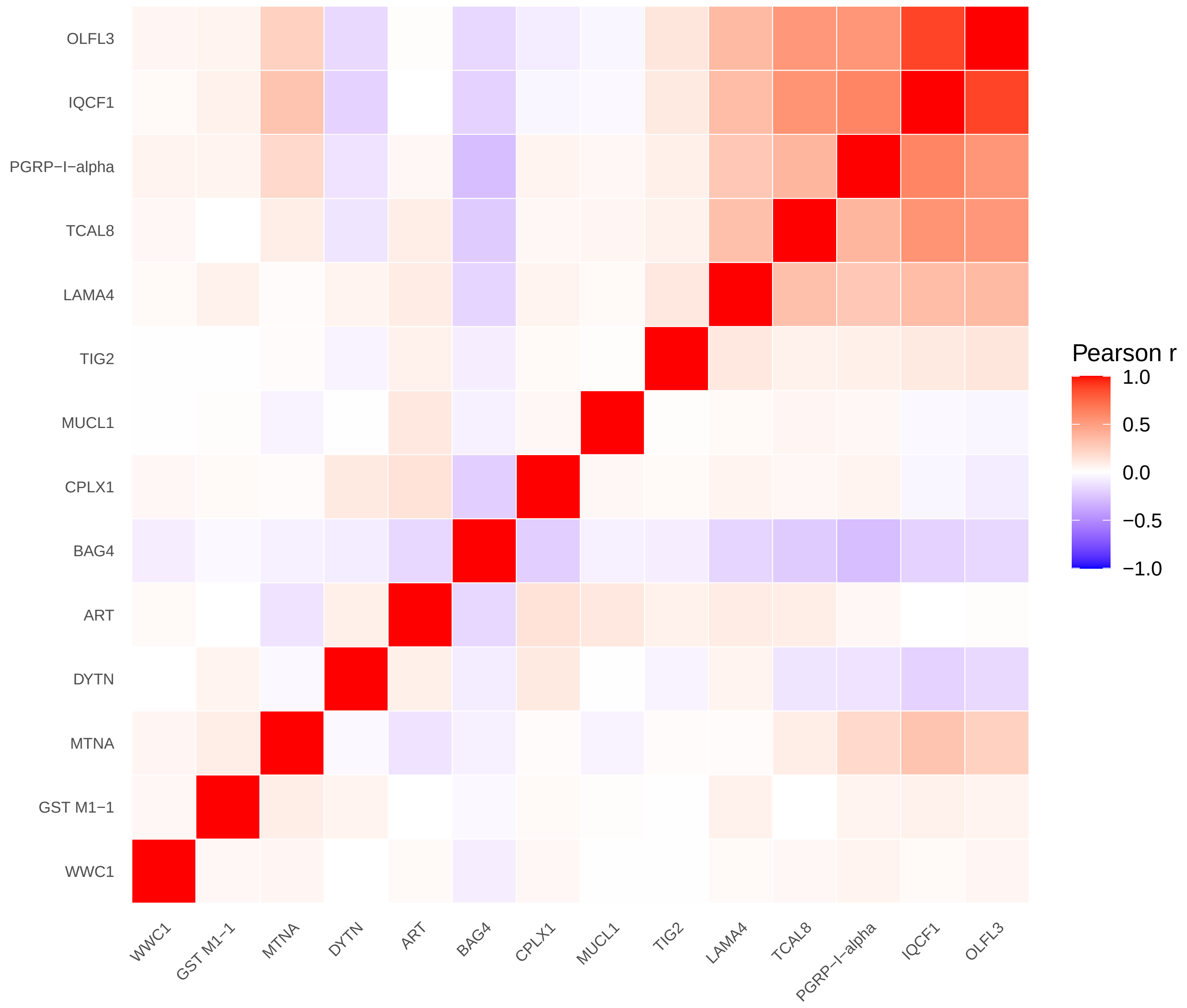


colon / male − cases (N=281)

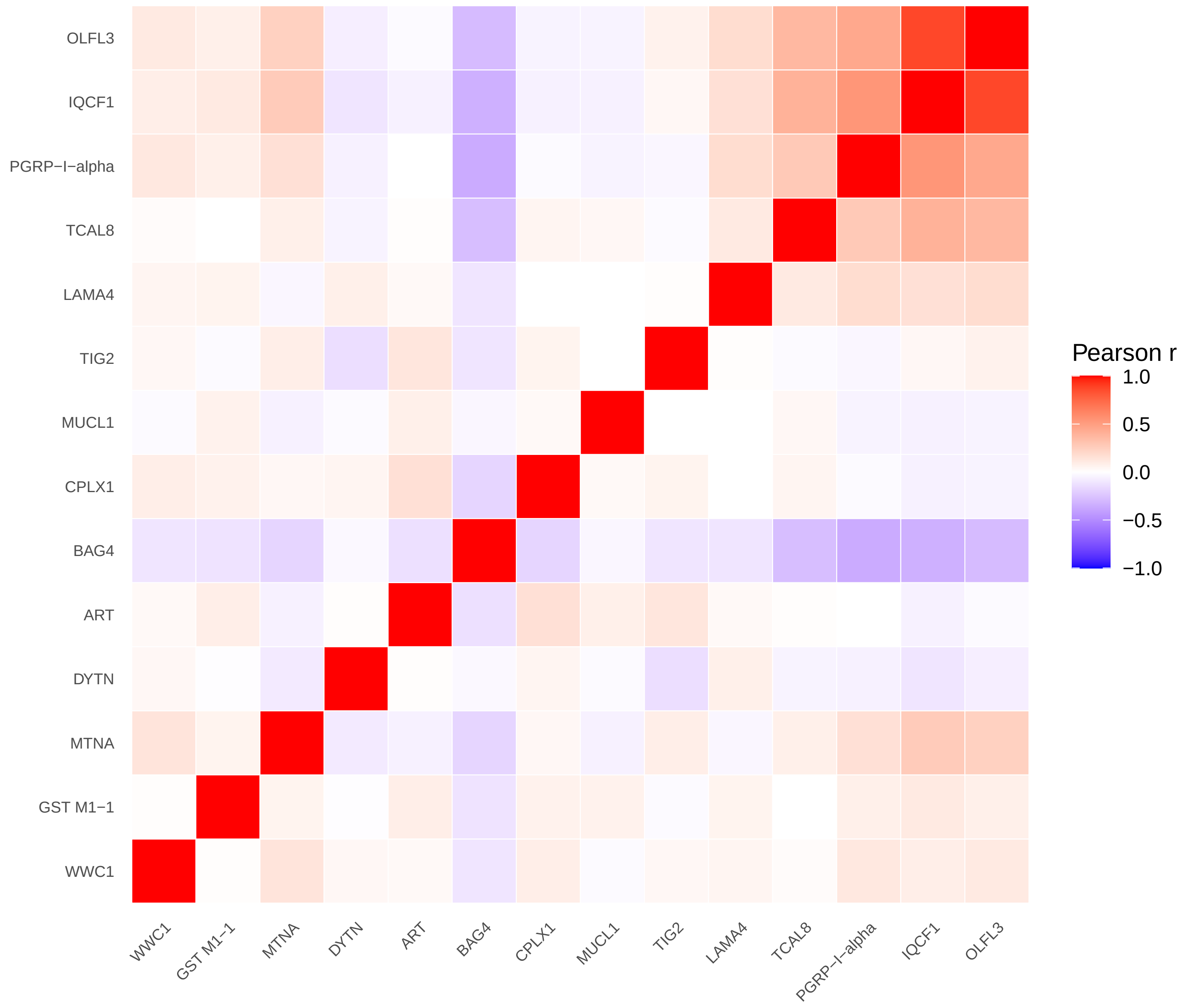


colon / male − non−cases (N=1556)

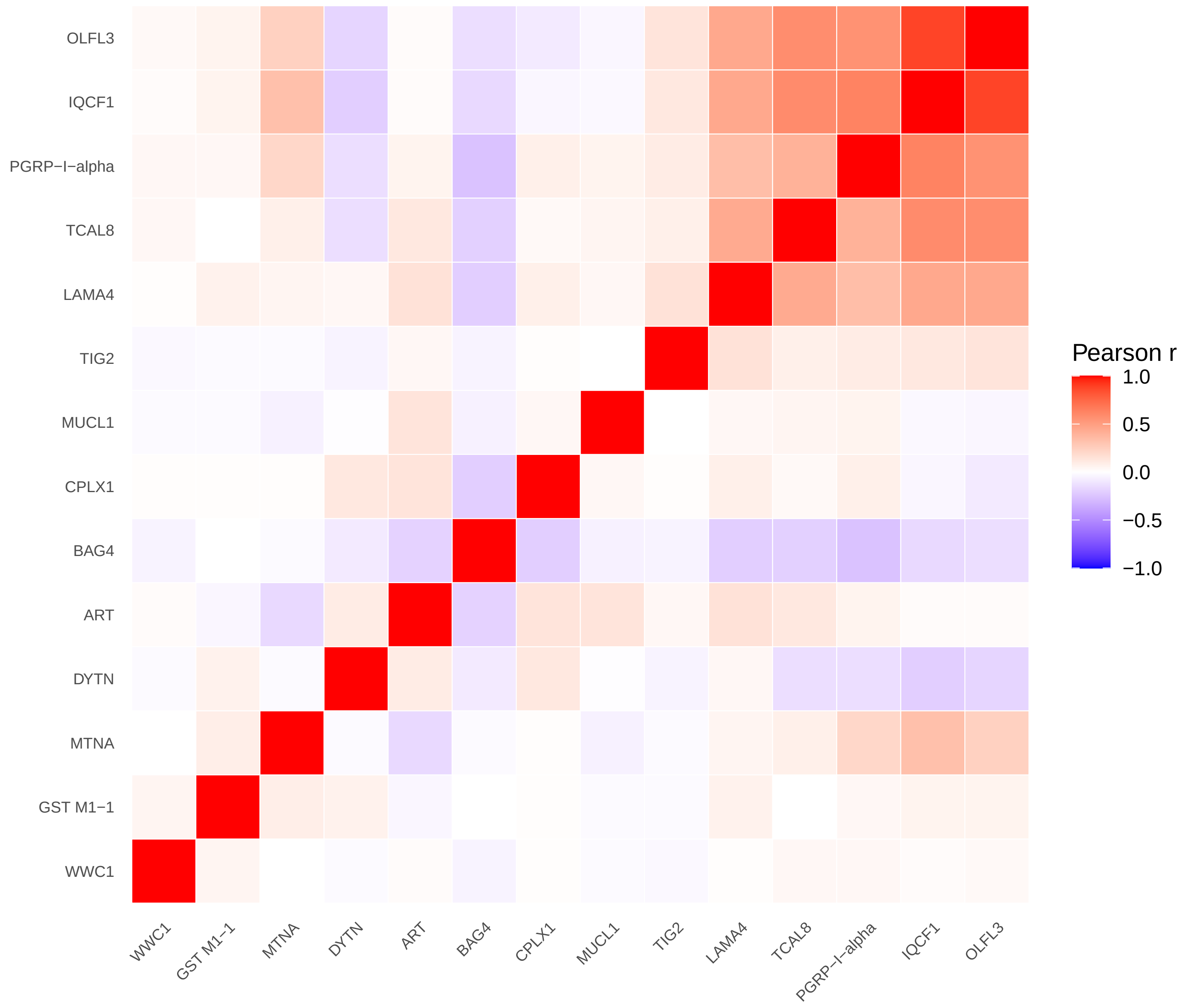

### rectum / combined − all (N=4430)

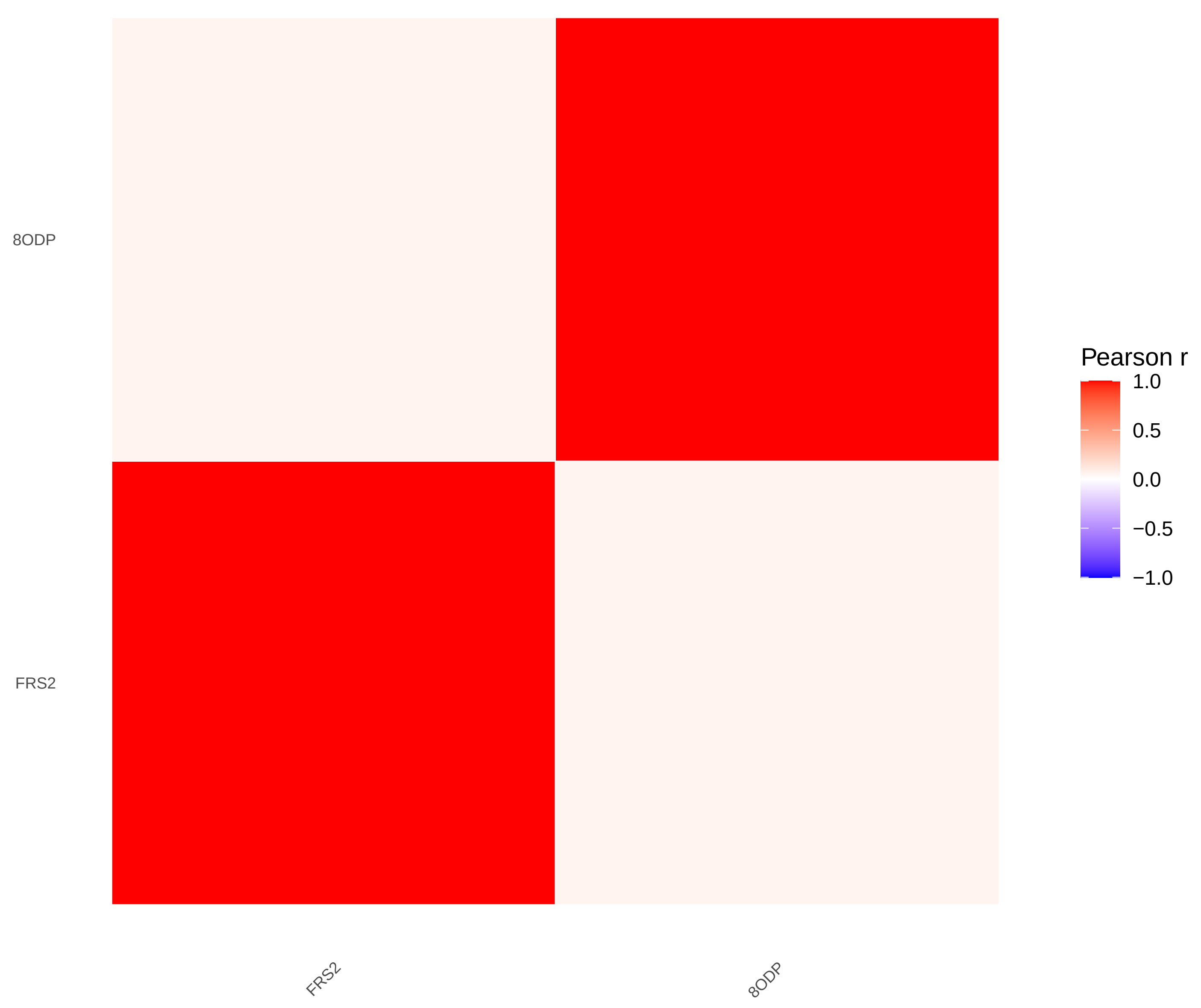


### rectum / combined − cases (N=319)

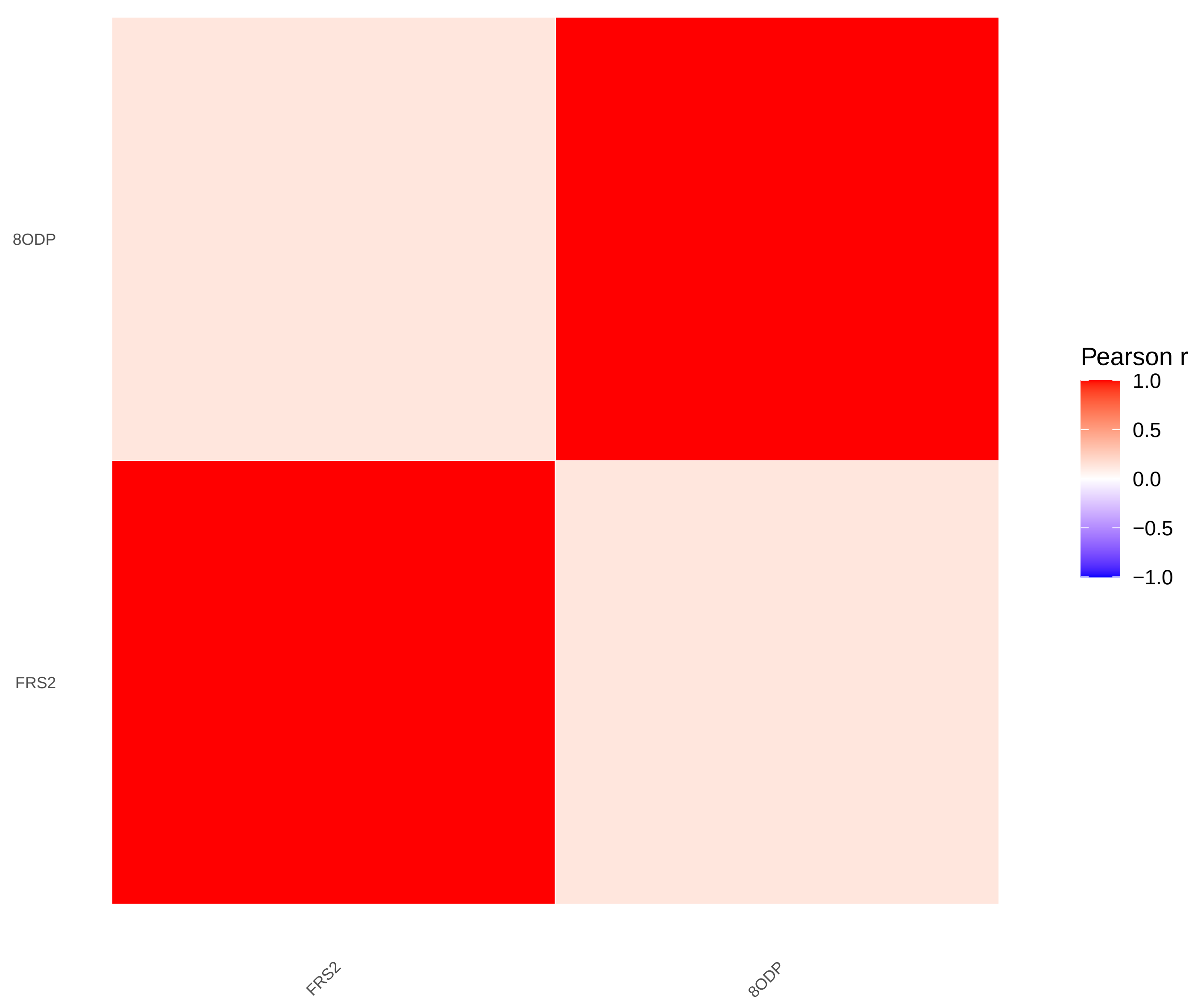


### rectum / combined − non−cases (N=4111)

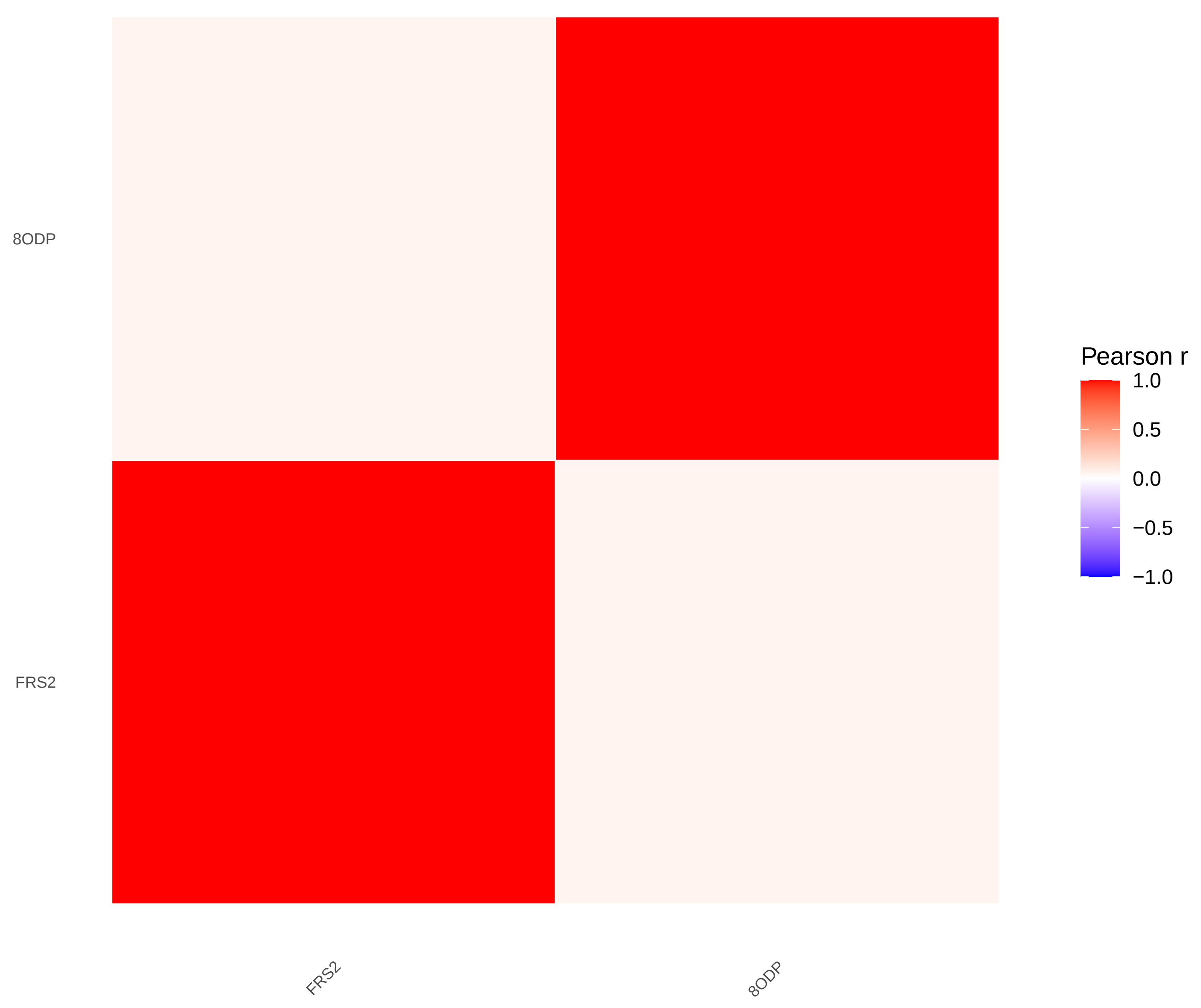

S−Figure 2 correlations: rectum / female
Not enough associated proteins (FDR < 0.05).

rectum / male − all (N=1747)

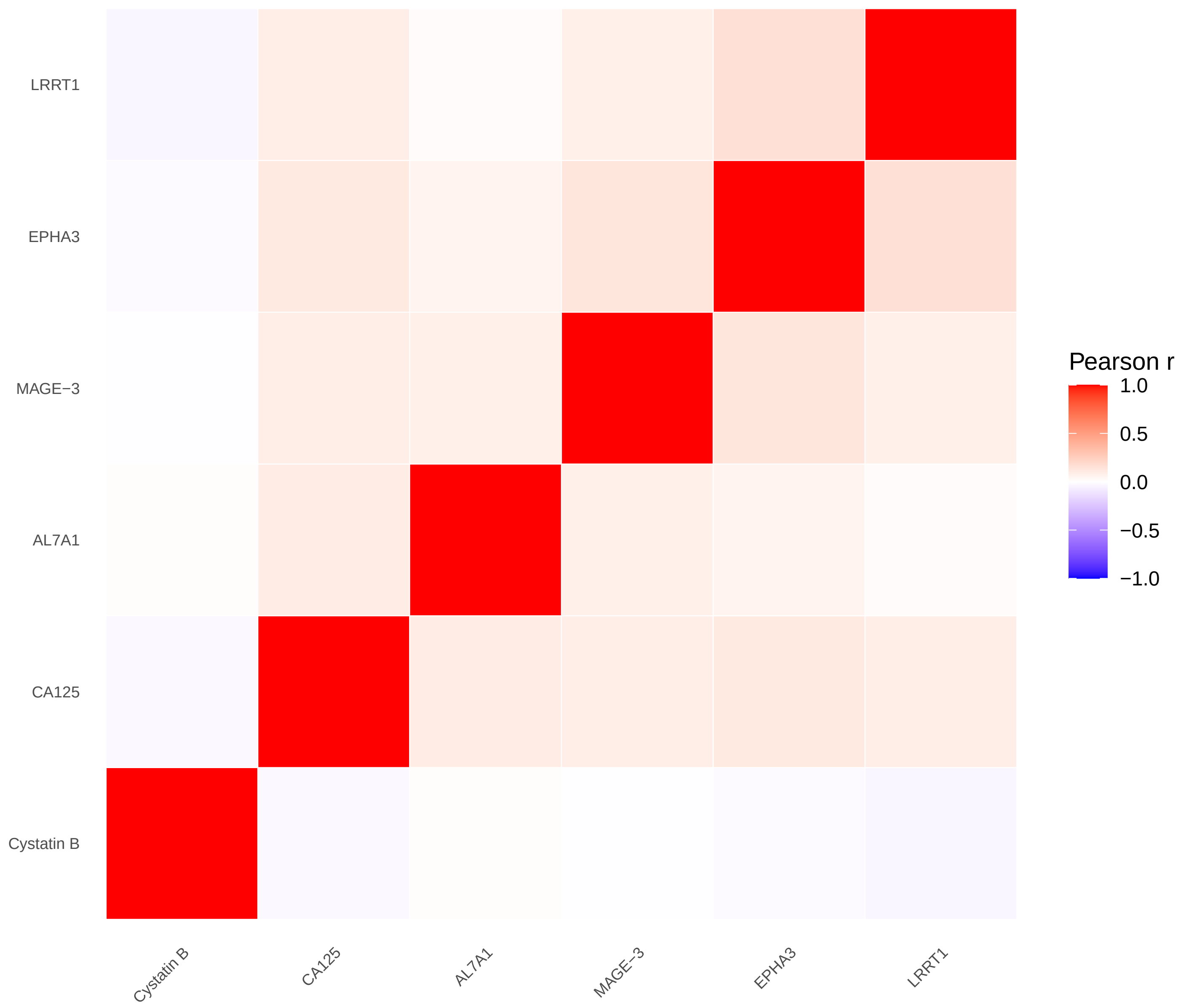


rectum / male − cases (N=177)

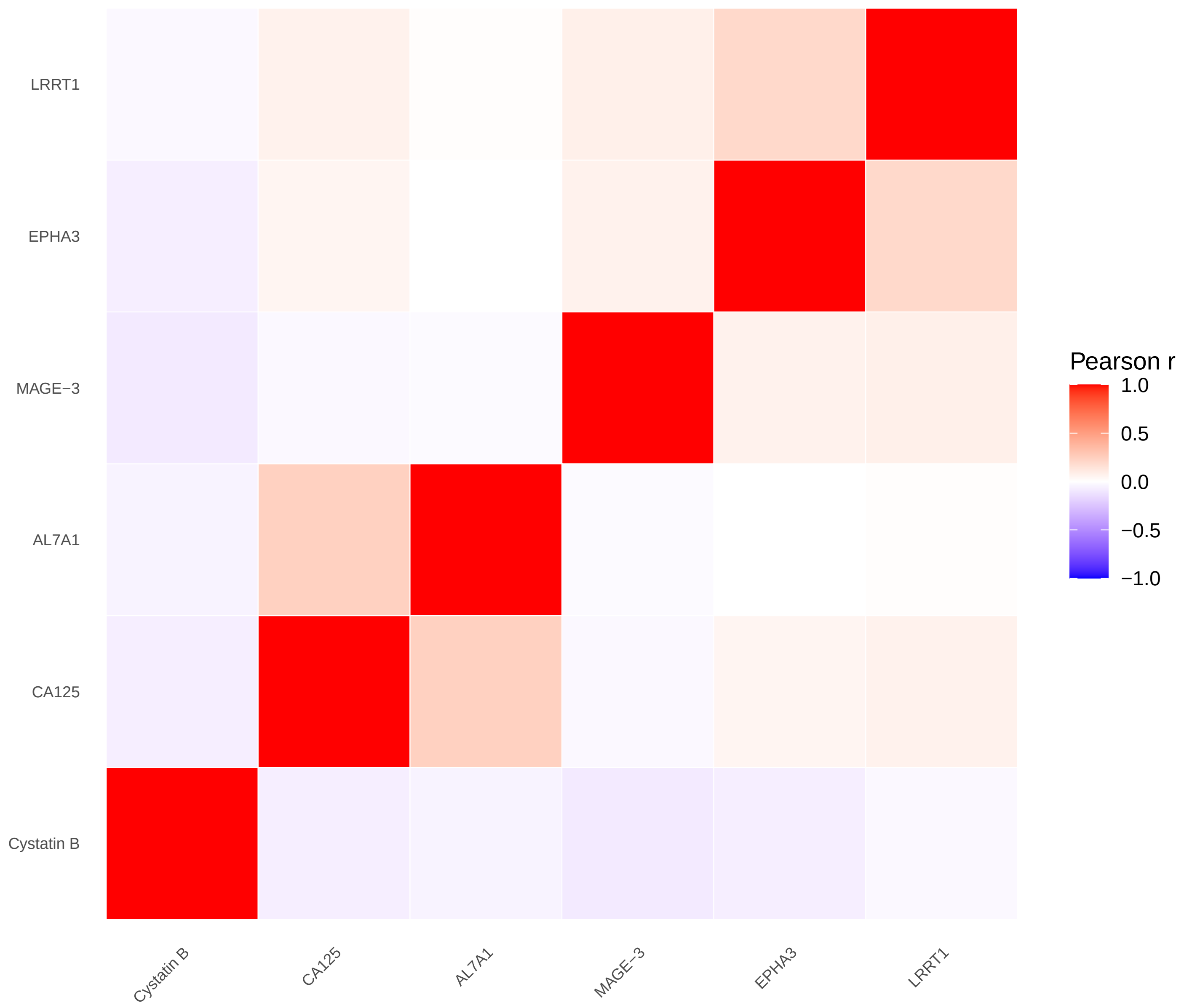


rectum / male − non−cases (N=1570)

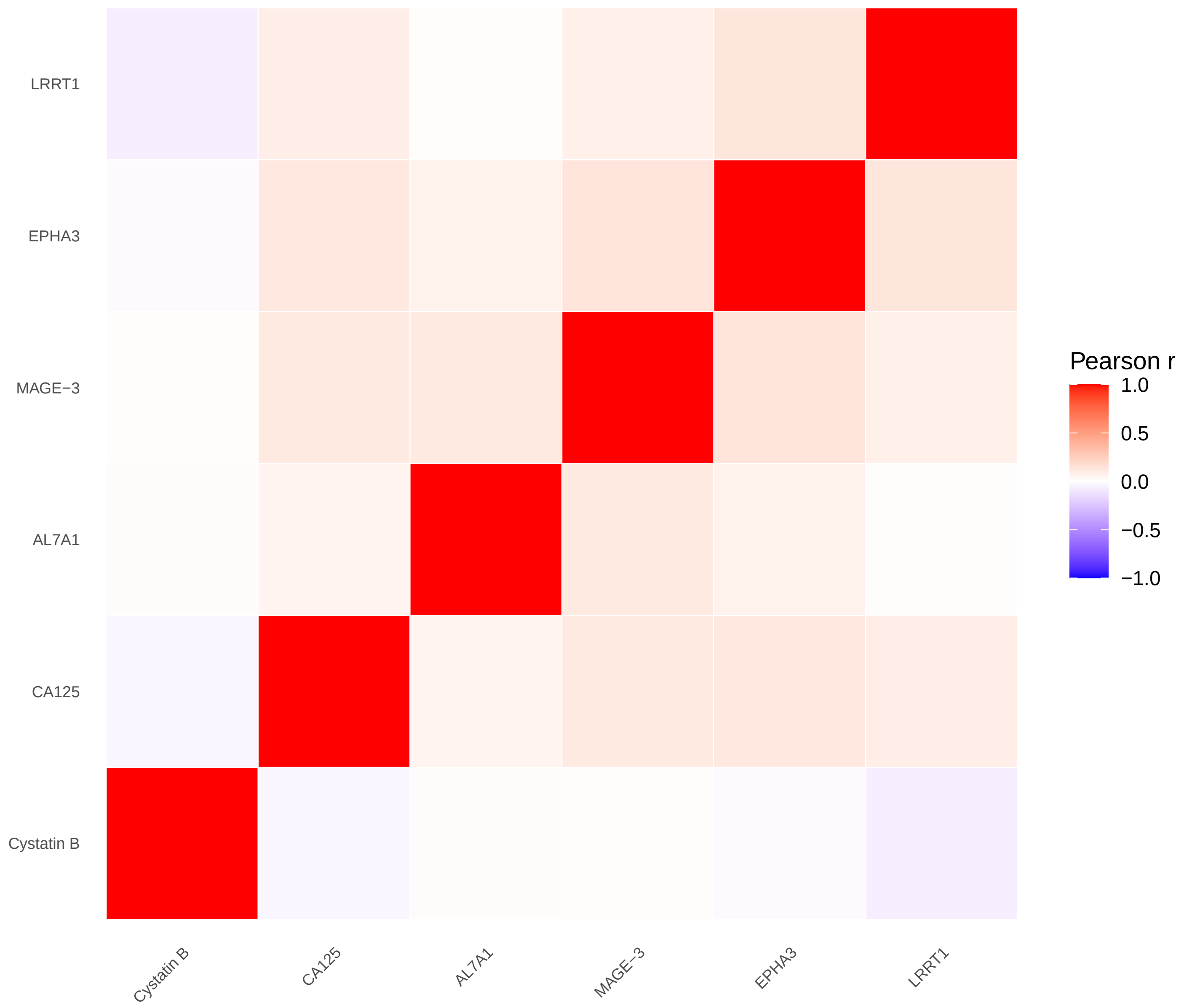

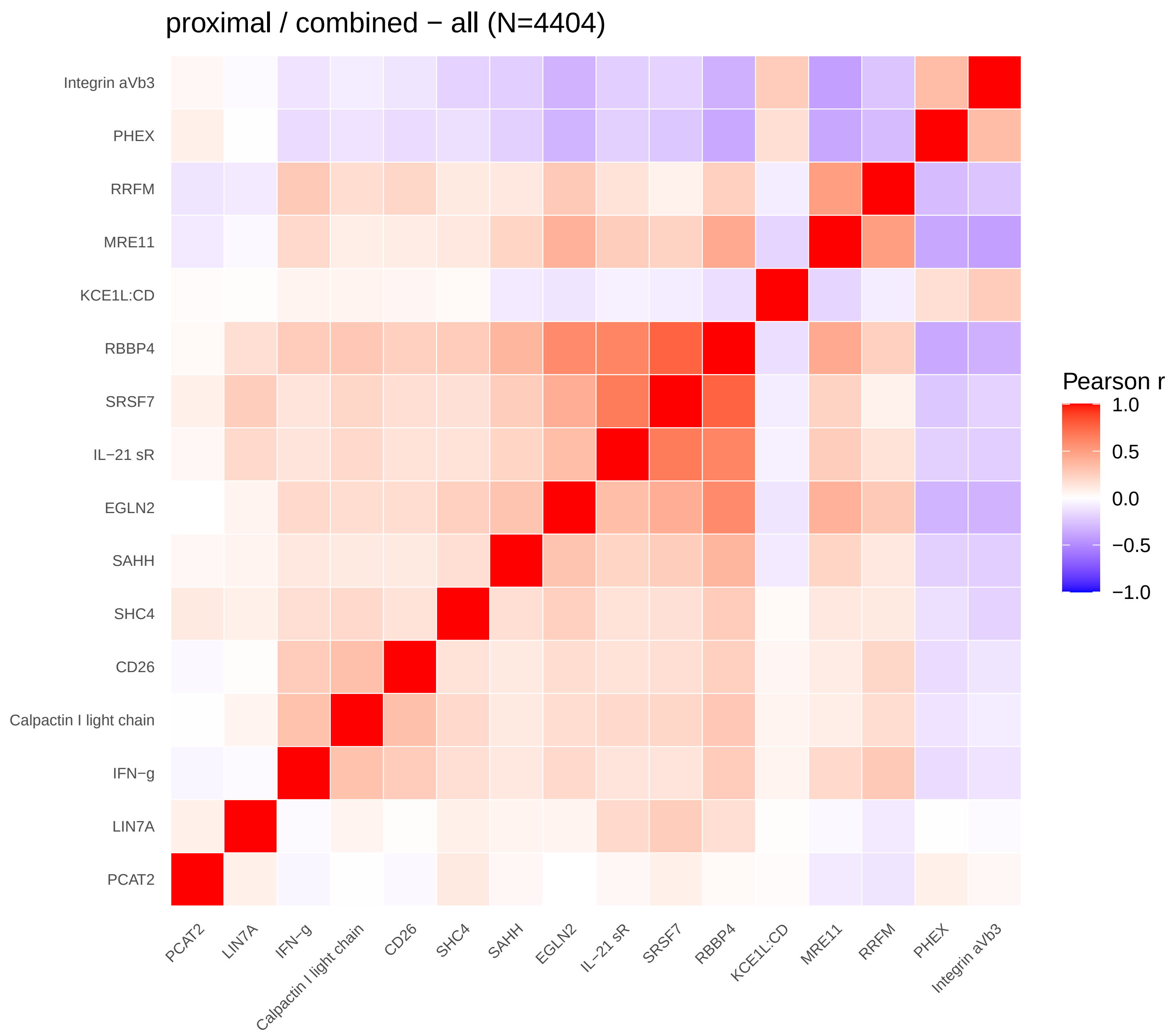
proximal / combined − all (N=4404)
Integrin aVb3
PHEX
RRFM
MRE11
KCE1L:CD
RBBP4
SRSF7
IL−21 sR
EGLN2
SAHH
SHC4
CD26
Calpactin I light chain
IFN−g
LIN7A
PCAT2
Pearson r
1.0
0.5
0.0
−0.5
−1.0

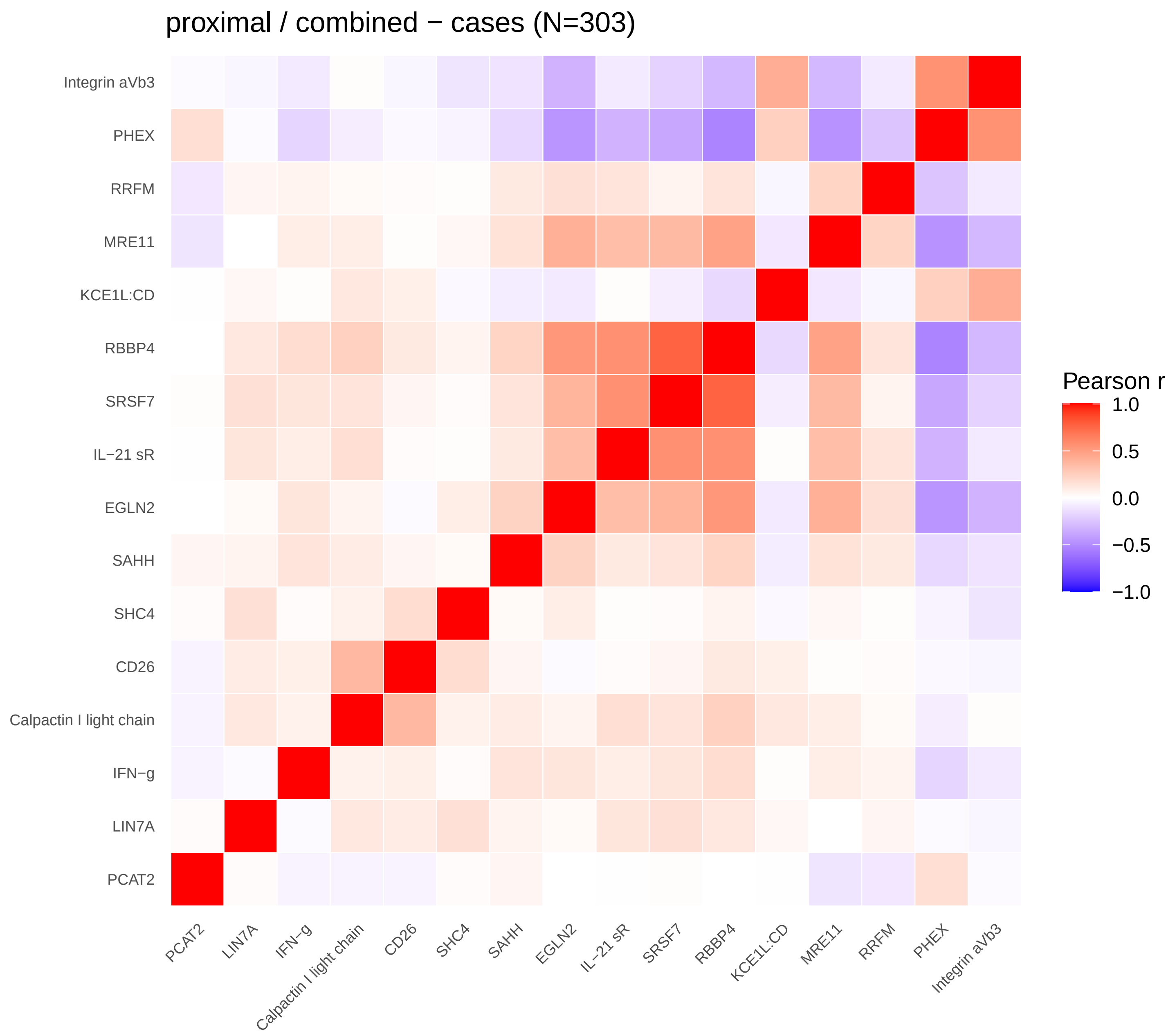
proximal / combined − cases (N=303)
Integrin aVb3
PHEX
RRFM
MRE11
KCE1L:CD
RBBP4
SRSF7
IL−21 sR
EGLN2
SAHH
SHC4
CD26
Calpactin I light chain
IFN−g
LIN7A
PCAT2
Pearson r
1.0
0.5
0.0
−0.5
−1.0

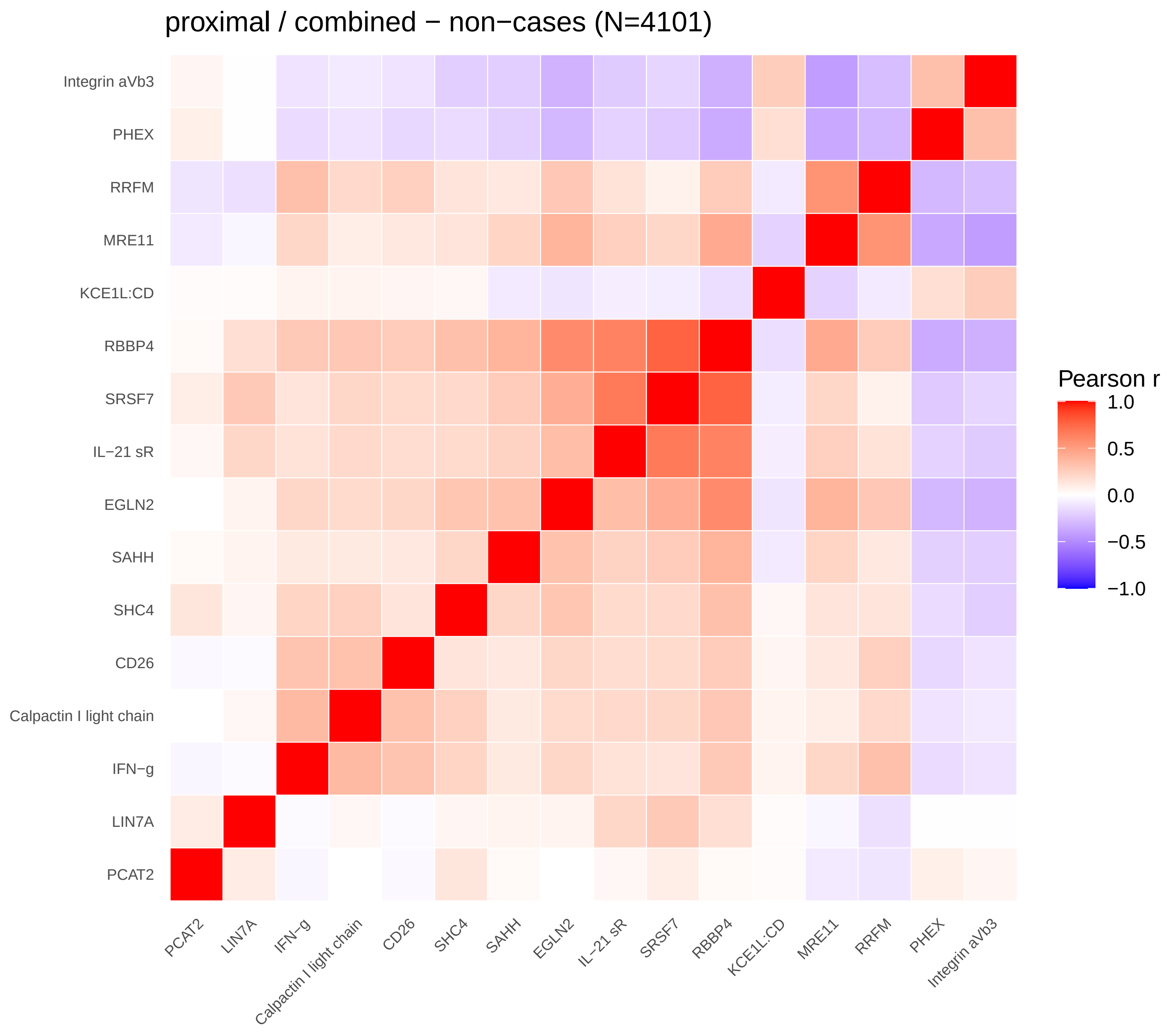
proximal / combined − non−cases (N=4101)
Integrin aVb3
PHEX
RRFM
MRE11
KCE1L:CD
RBBP4
SRSF7
IL−21 sR
EGLN2
SAHH
SHC4
CD26
Calpactin I light chain
IFN−g
LIN7A
PCAT2
Pearson r
1.0
0.5
0.0
−0.5
−1.0

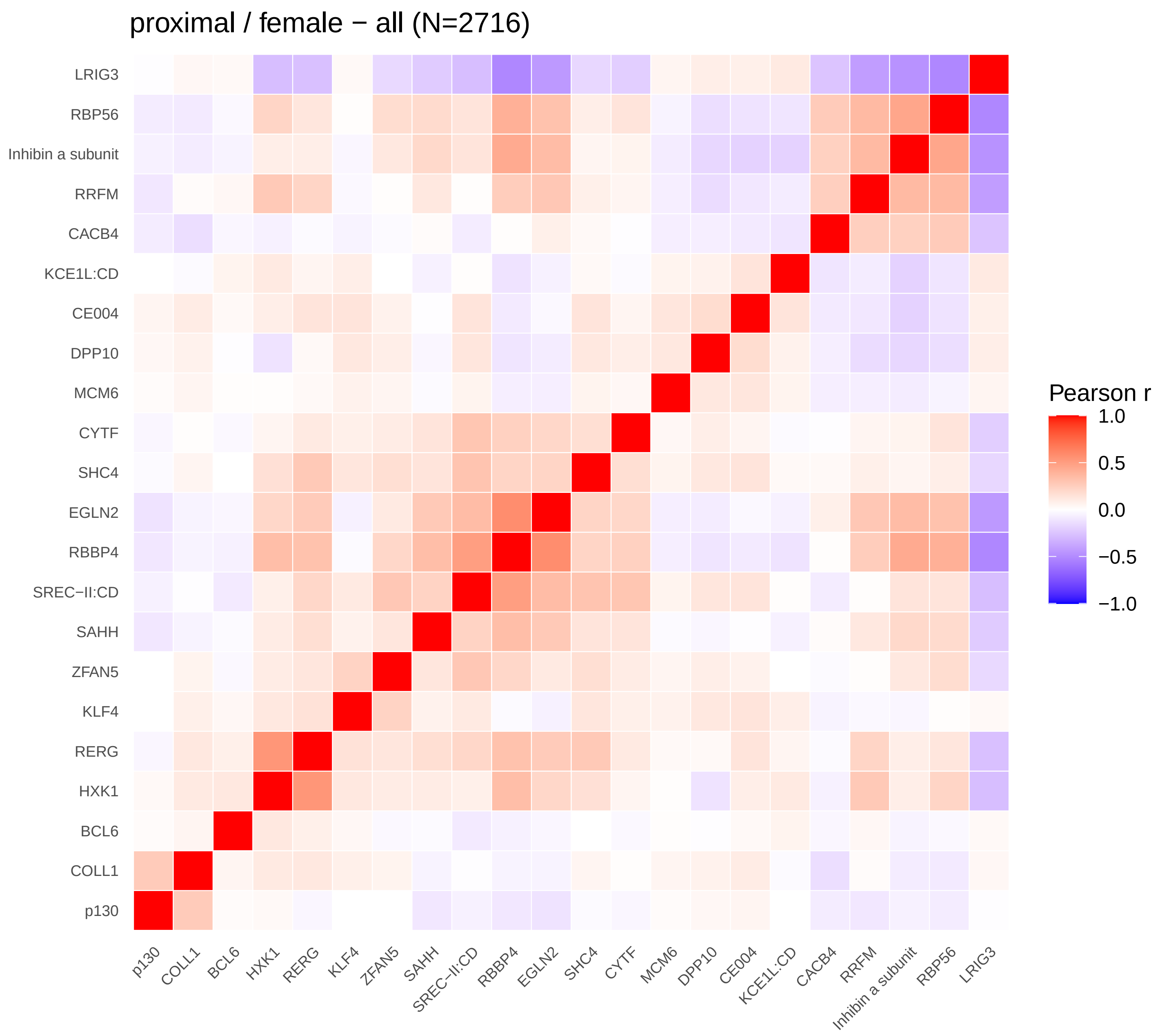
proximal / female − all (N=2716)
LRIG3
RBP56
Inhibin a subunit
RRFM
CACB4
KCE1L:CD
CE004
DPP10
MCM6
CYTF
SHC4
EGLN2
RBBP4
SREC−II:CD
SAHH
ZFAN5
KLF4
RERG
HXK1
BCL6
COLL1
p130
Pearson r
1.0
0.5
0.0
−0.5
−1.0

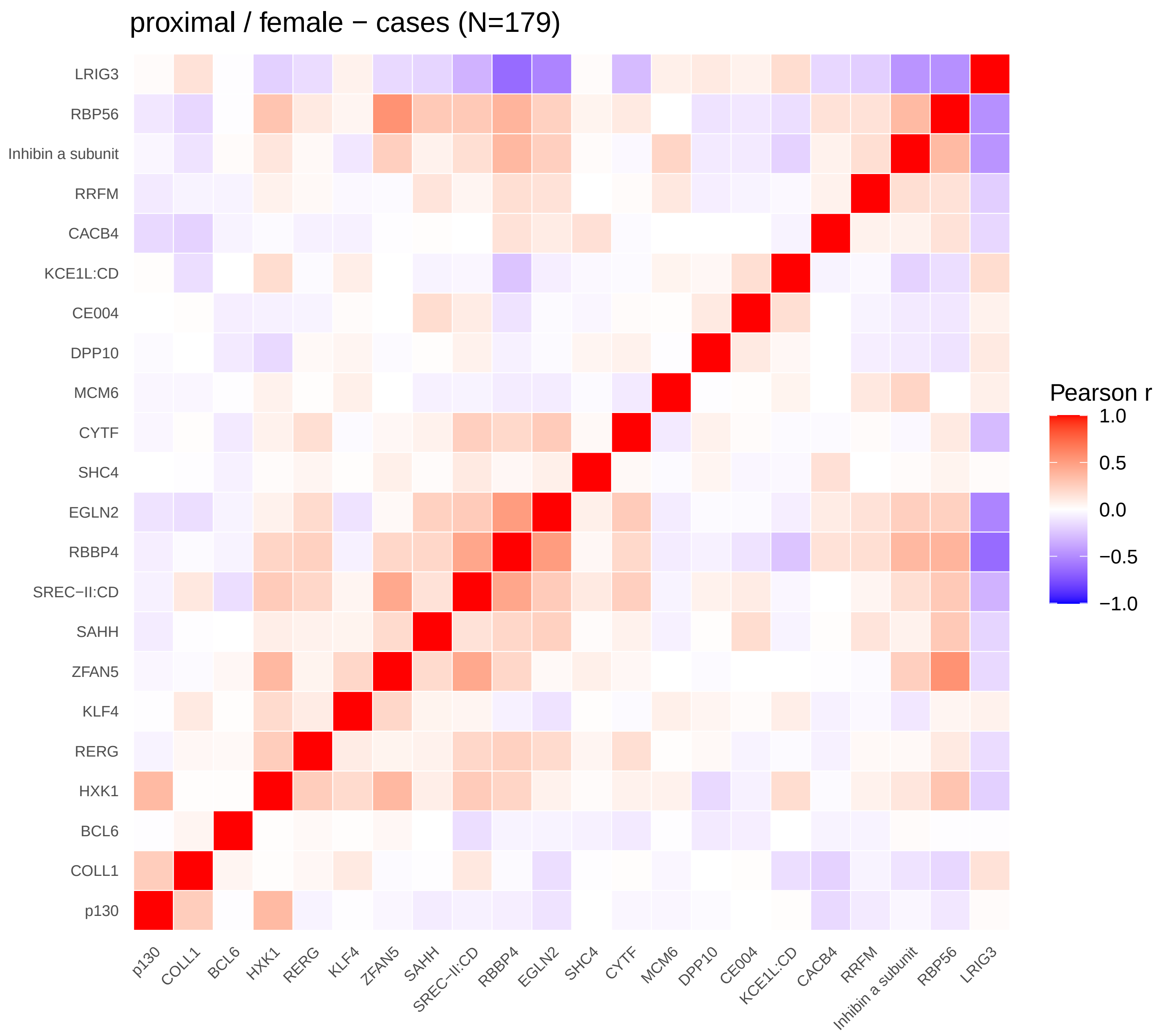
proximal / female − cases (N=179)
LRIG3
RBP56
Inhibin a subunit
RRFM
CACB4
KCE1L:CD
CE004
DPP10
MCM6
CYTF
SHC4
EGLN2
RBBP4
SREC−II:CD
SAHH
ZFAN5
KLF4
RERG
HXK1
BCL6
COLL1
p130
Pearson r
1.0
0.5
0.0
−0.5
−1.0

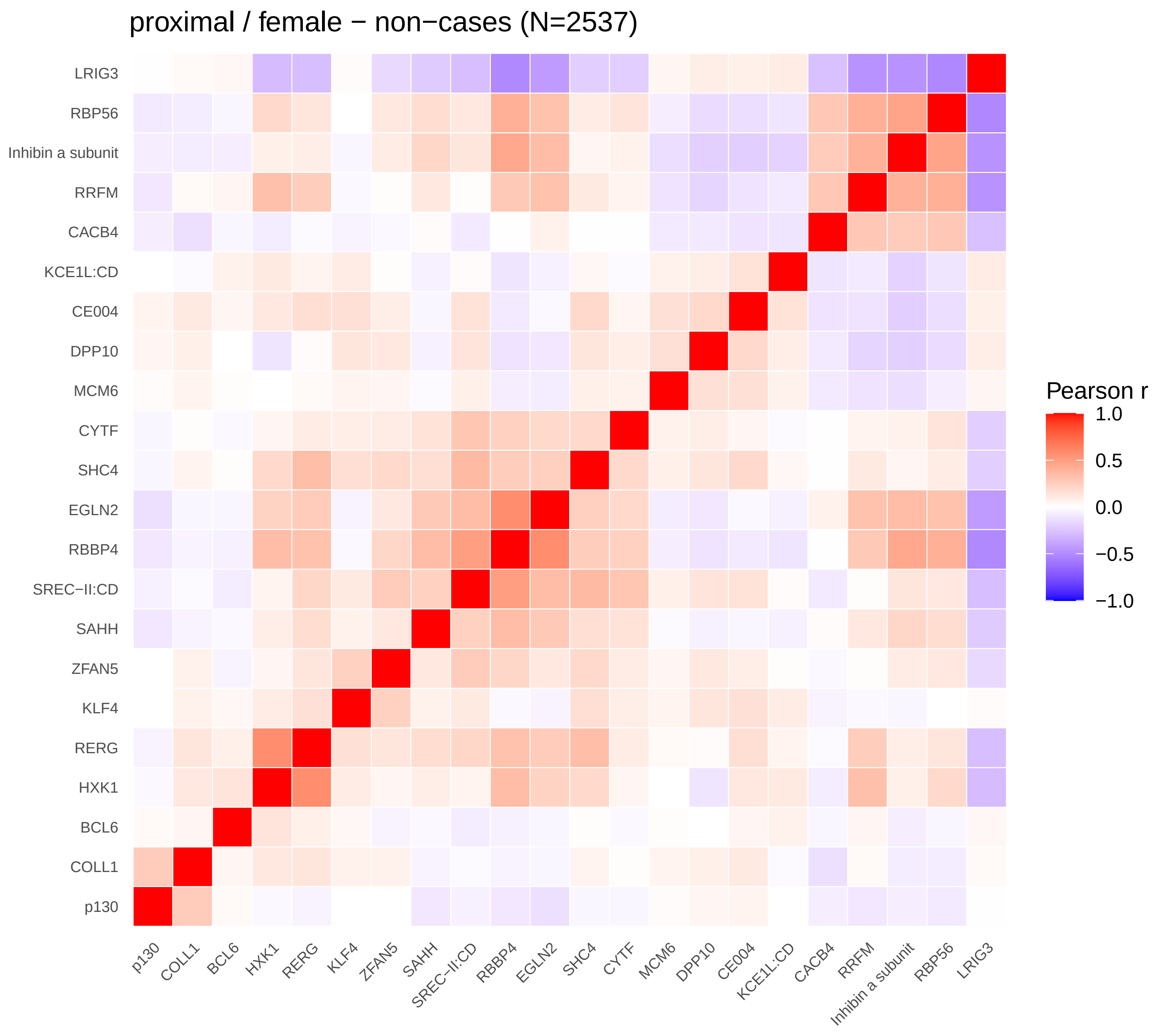
proximal / female − non−cases (N=2537)
LRIG3
RBP56
Inhibin a subunit
RRFM
CACB4
KCE1L:CD
CE004
DPP10
MCM6
CYTF
SHC4
EGLN2
RBBP4
SREC−II:CD
SAHH
ZFAN5
KLF4
RERG
HXK1
BCL6
COLL1
p130
Pearson r
1.0
0.5
0.0
−0.5
−1.0

S−Figure 2 correlations: proximal / male
Not enough associated proteins (FDR < 0.05).

distal / combined − all (N=4405)

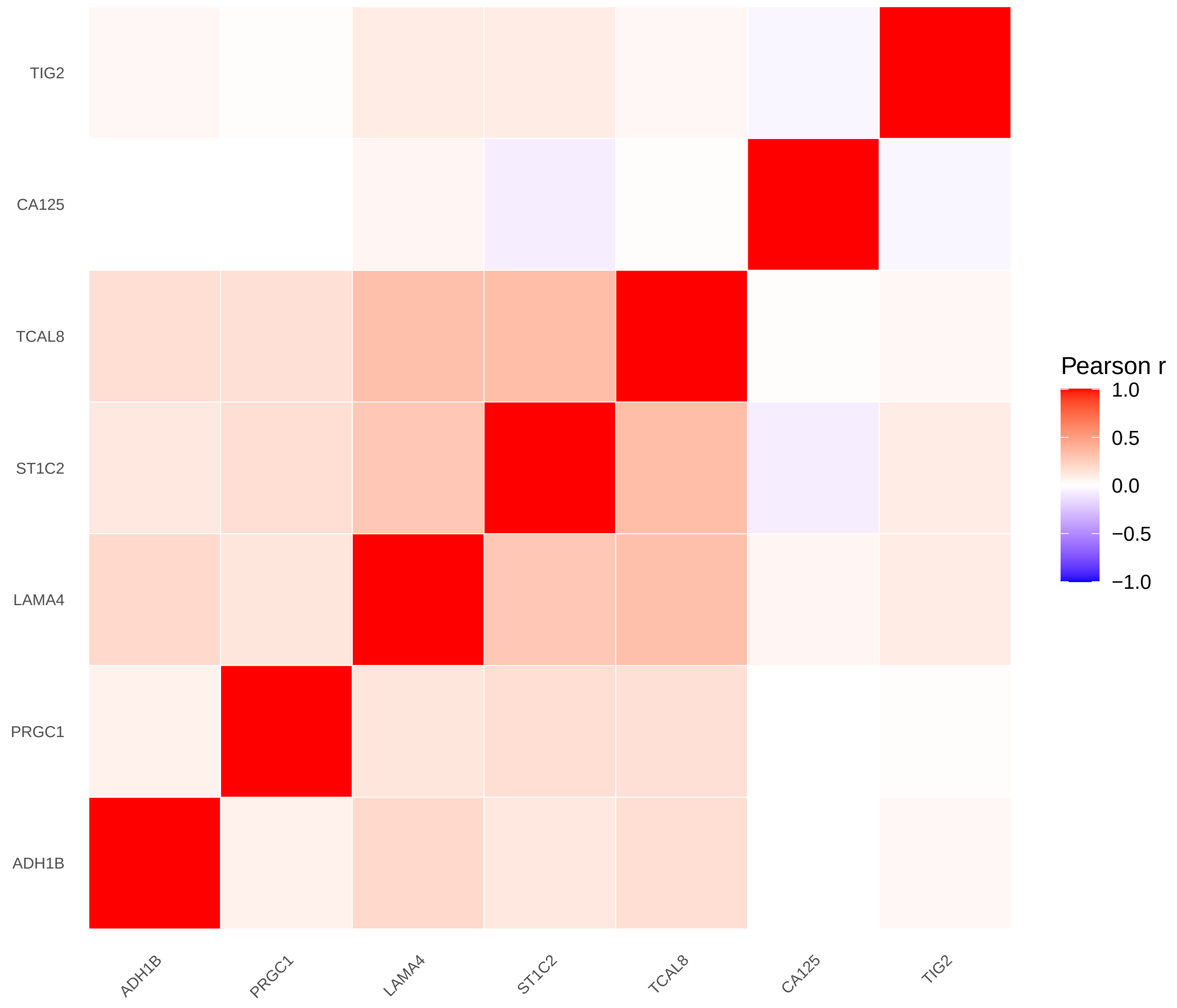


distal / combined − cases (N=304)

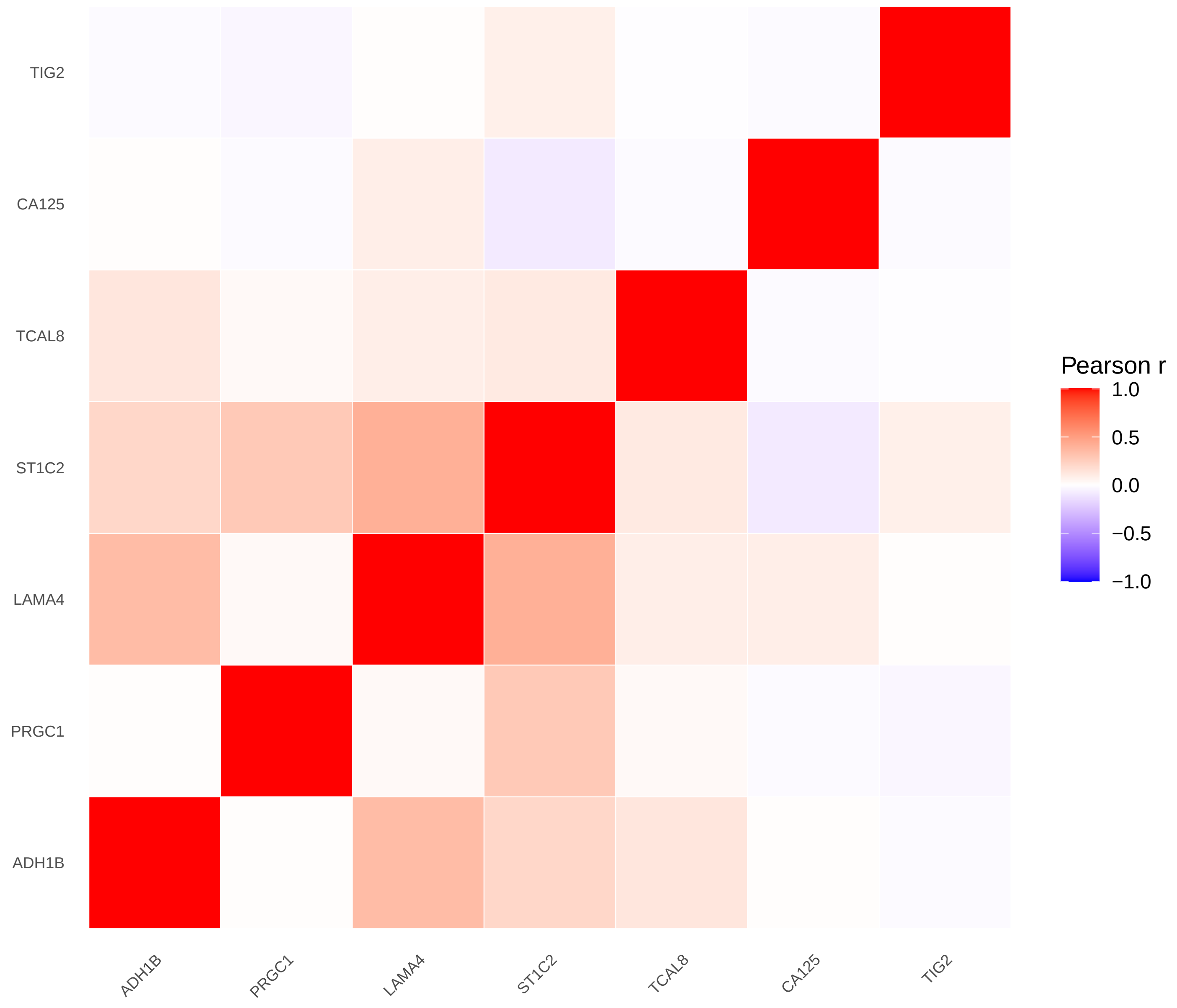


distal / combined − non−cases (N=4101)

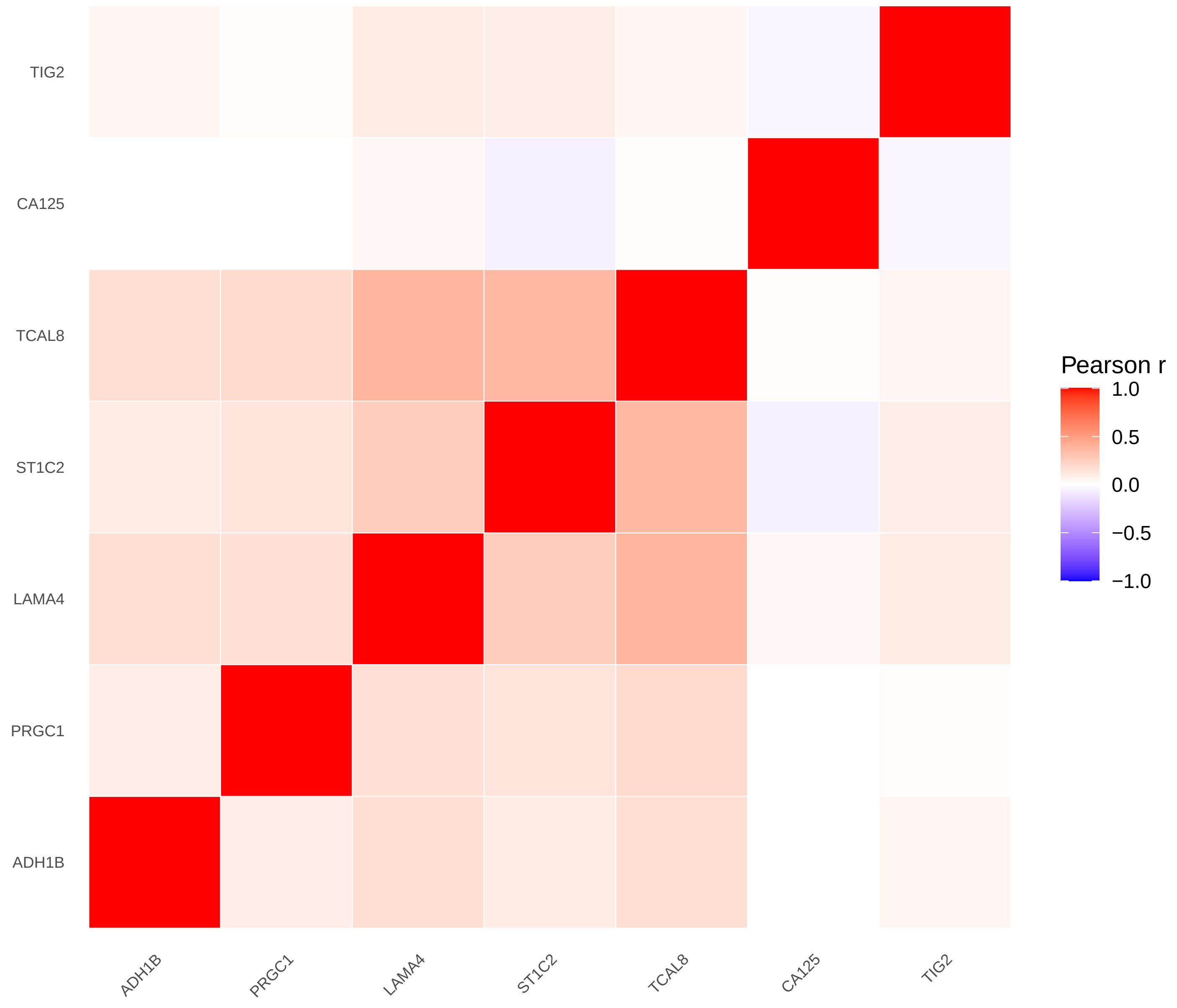

distal / female − all (N=2705)

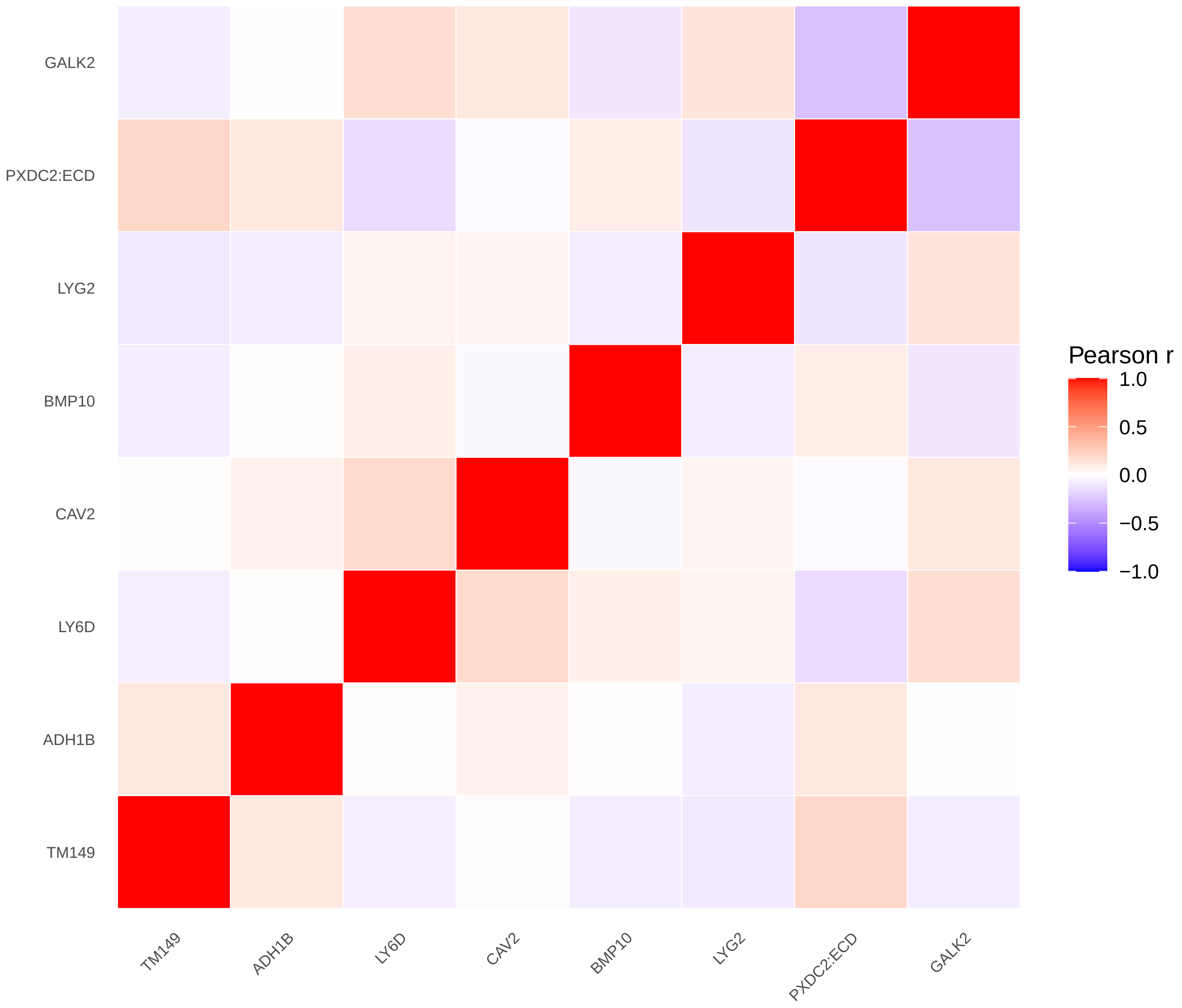


distal / female − cases (N=170)

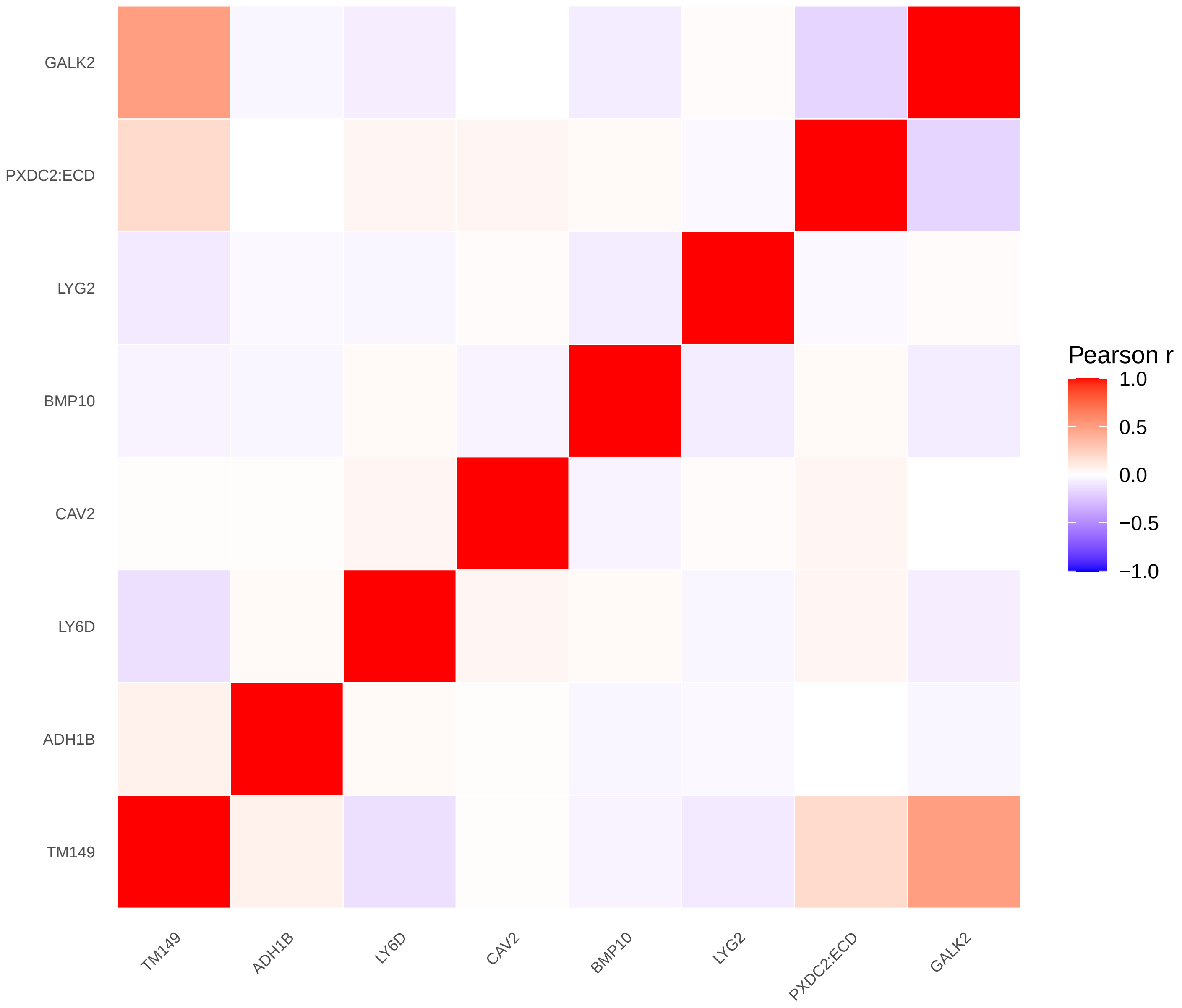


distal / female − non−cases (N=2535)

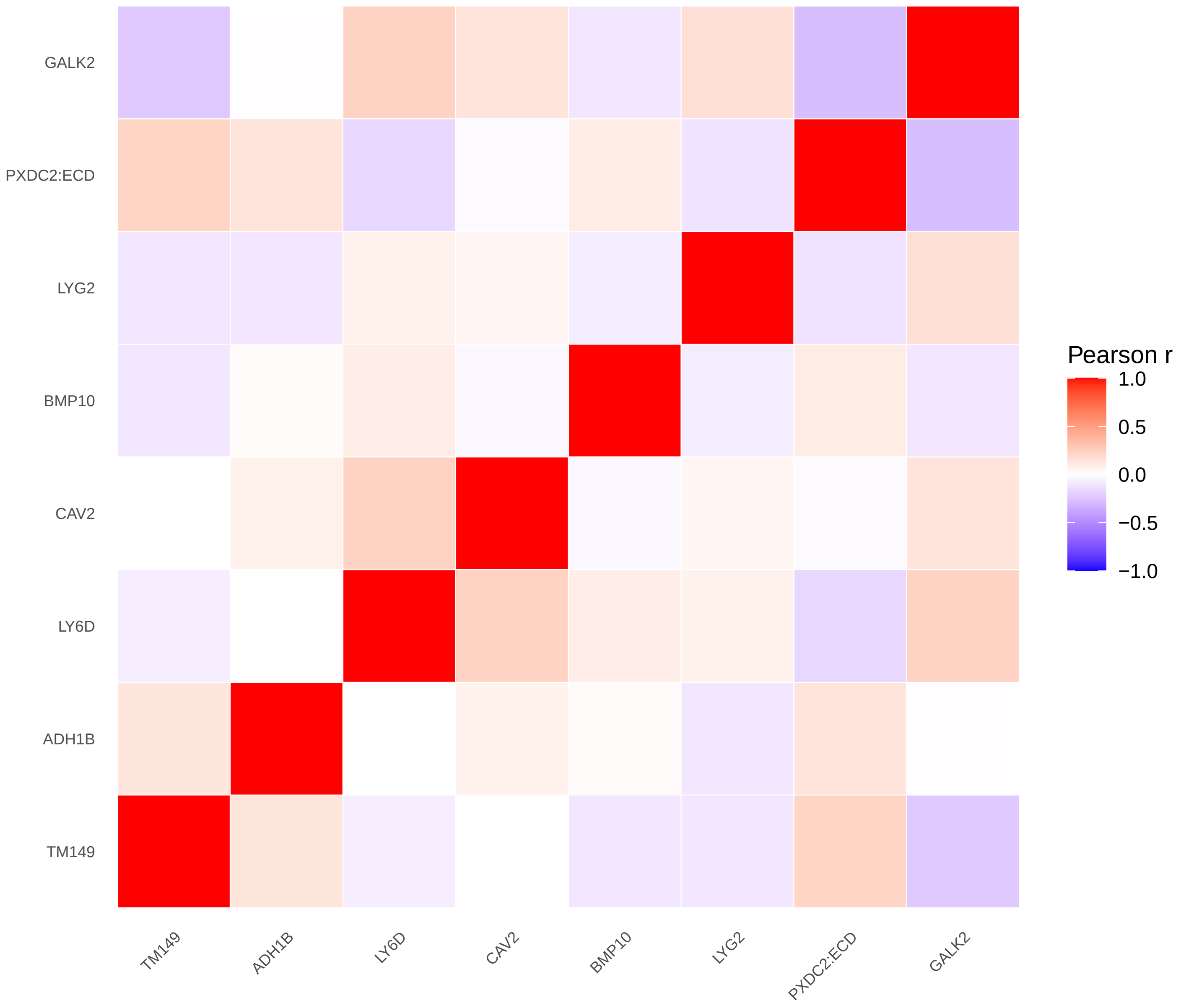

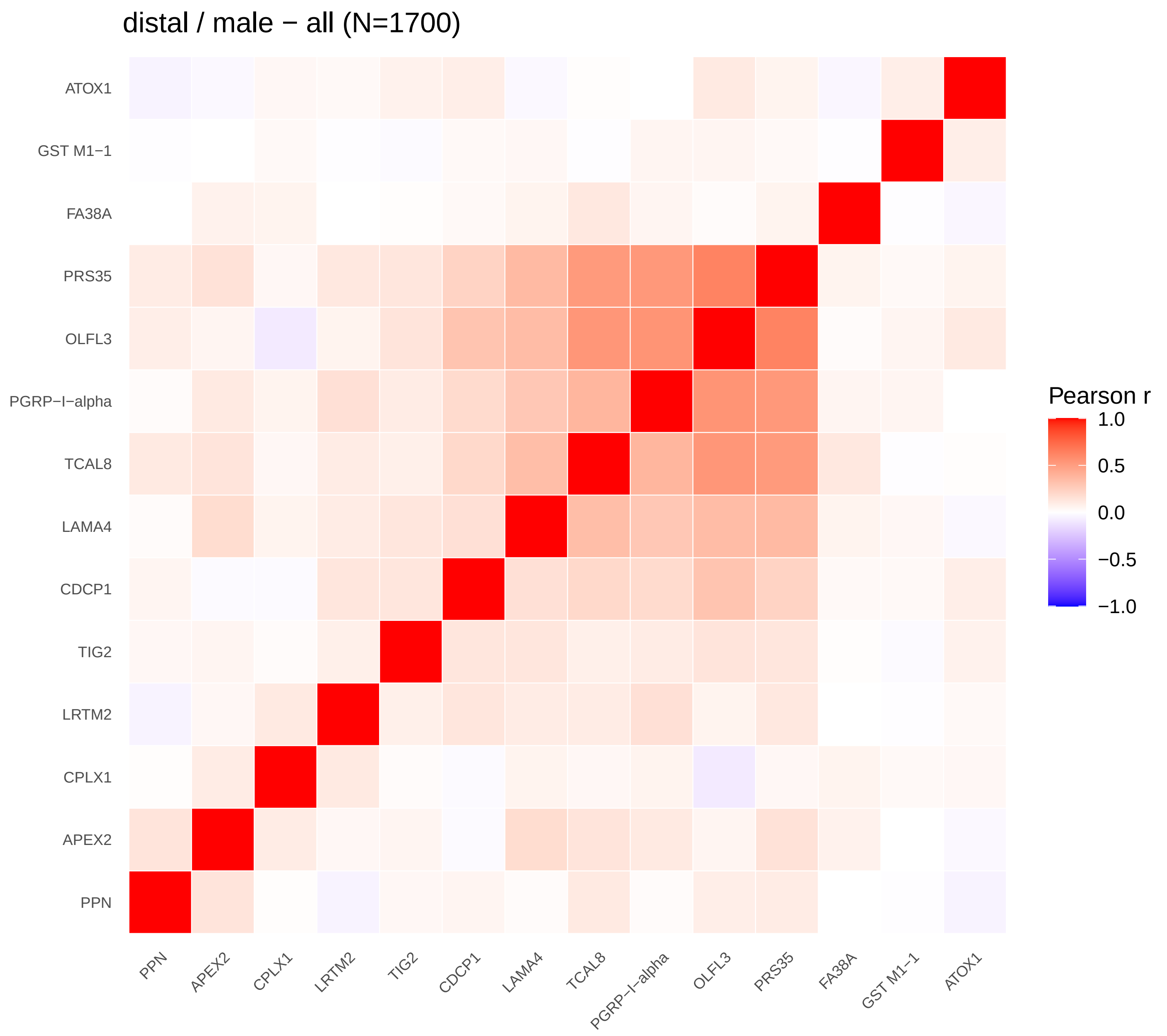
distal / male − all (N=1700)
ATOX1
GST M1−1
FA38A
PRS35
OLFL3
PGRP−I−alpha
TCAL8
LAMA4
CDCP1
TIG2
LRTM2
CPLX1
APEX2
PPN
Pearson r
1.0
0.5
0.0
−0.5
−1.0

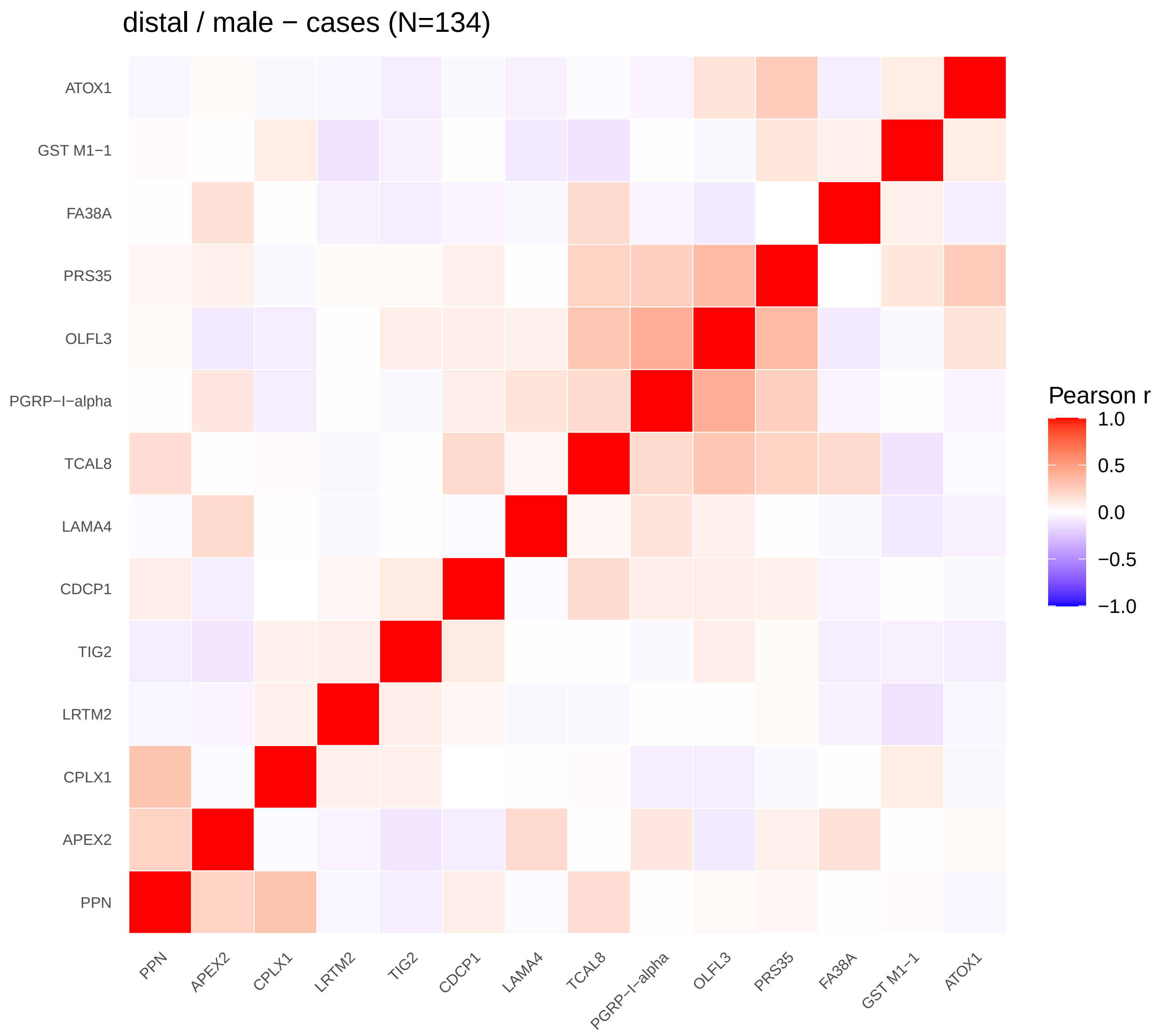
distal / male − cases (N=134)
ATOX1
GST M1−1
FA38A
PRS35
OLFL3
PGRP−I−alpha
TCAL8
LAMA4
CDCP1
TIG2
LRTM2
CPLX1
APEX2
PPN
Pearson r
1.0
0.5
0.0
−0.5
−1.0

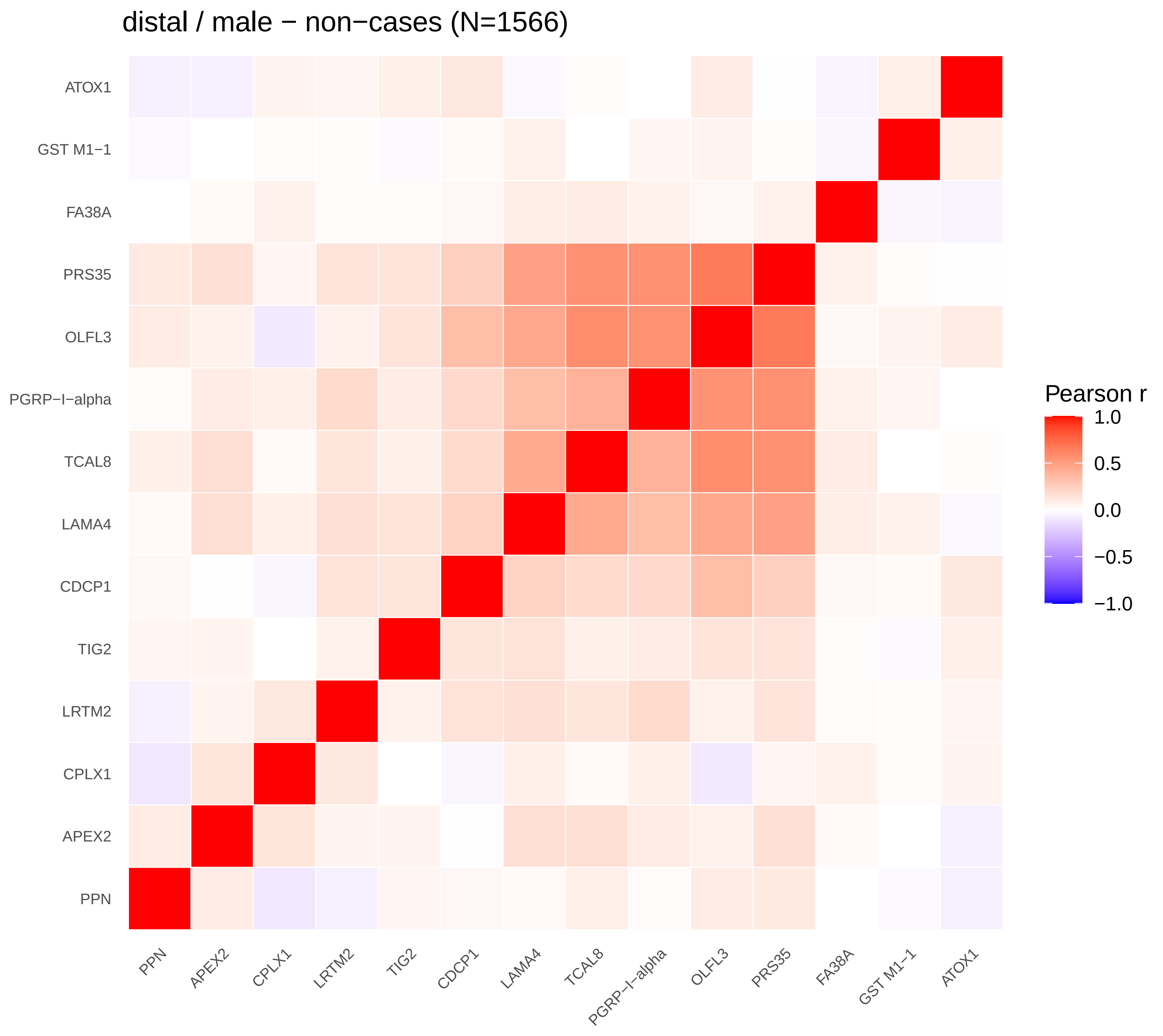
distal / male − non−cases (N=1566)
ATOX1
GST M1−1
FA38A
PRS35
OLFL3
PGRP−I−alpha
TCAL8
LAMA4
CDCP1
TIG2
LRTM2
CPLX1
APEX2
PPN
Pearson r
1.0
0.5
0.0
−0.5
−1.0
