## Supplementary Figure 3 for "Proteomic markers of colorectal cancer risk: results from the European Prospective Investigation into Cancer and Nutrition"

colorectal / combined − all

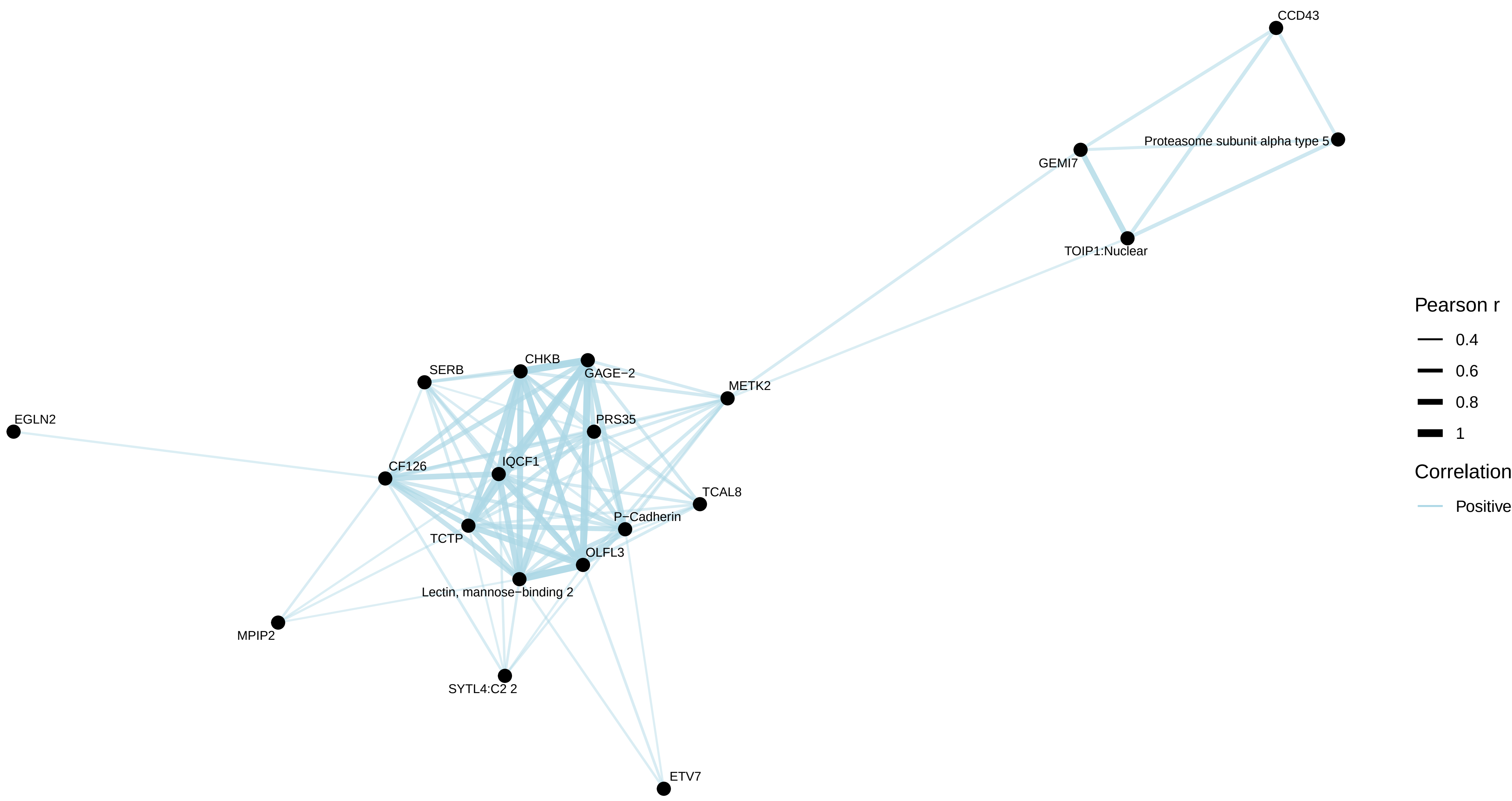


colorectal / combined − cases

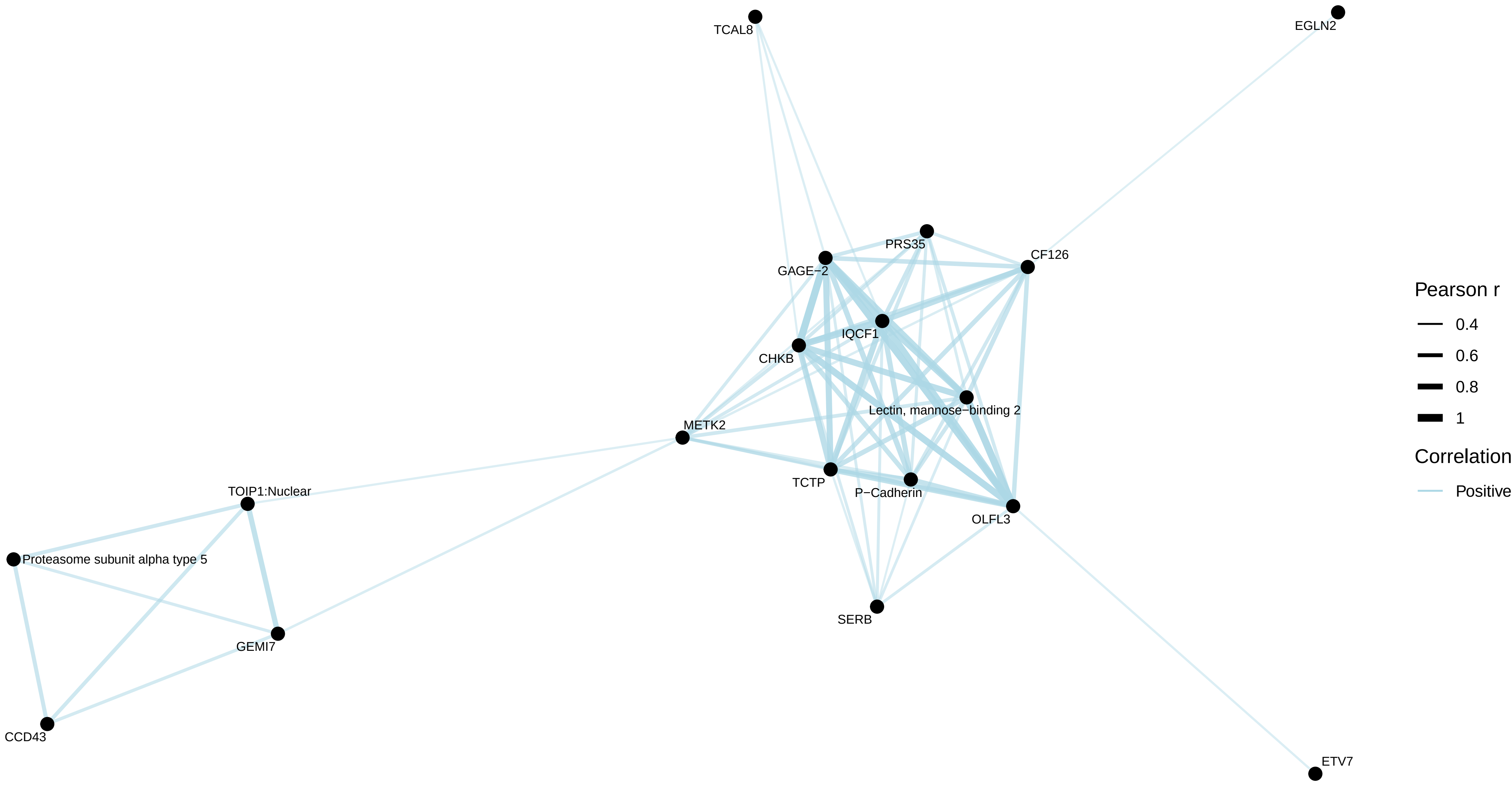


colorectal / combined − non−cases

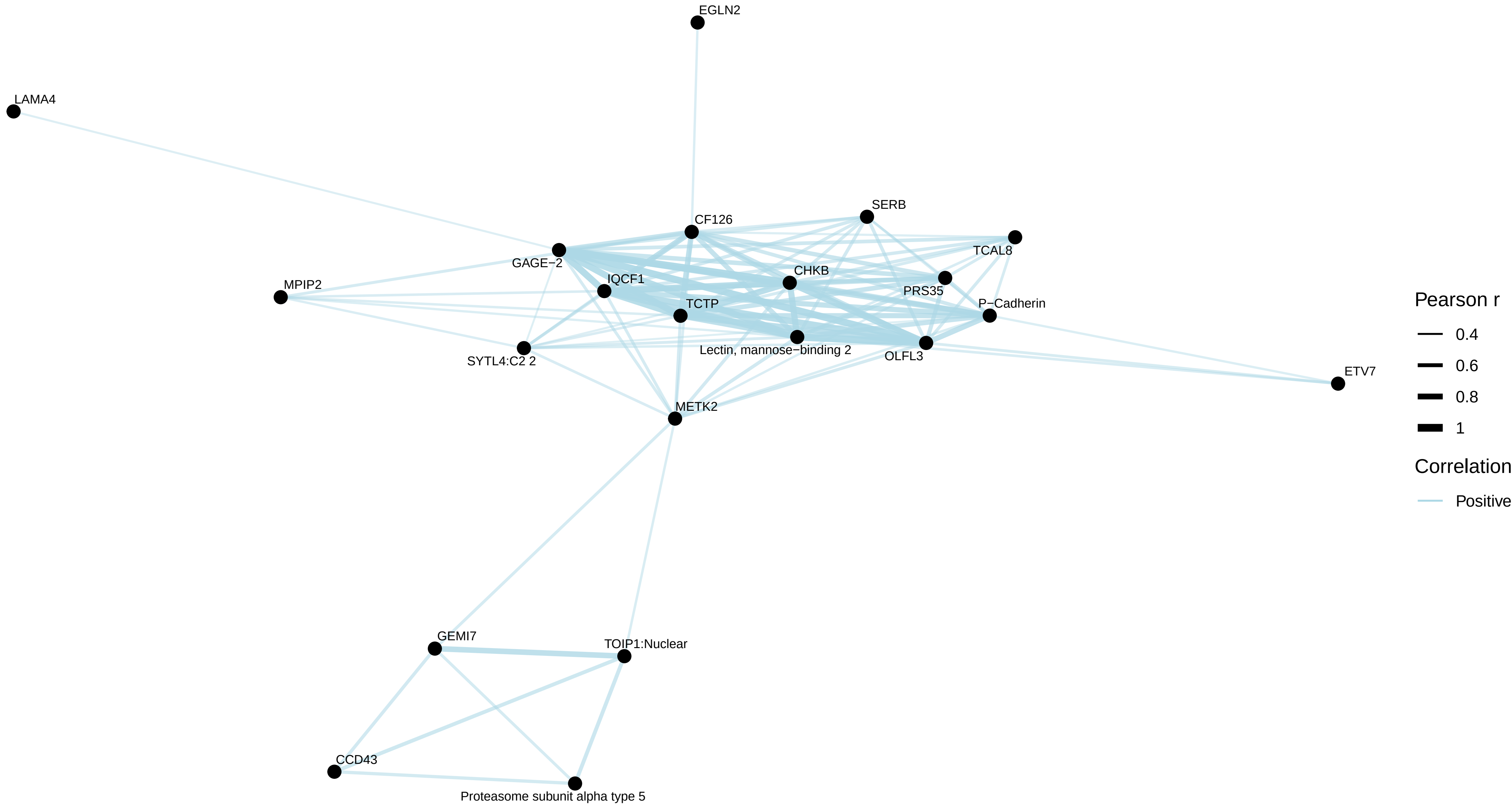

colorectal / female − all

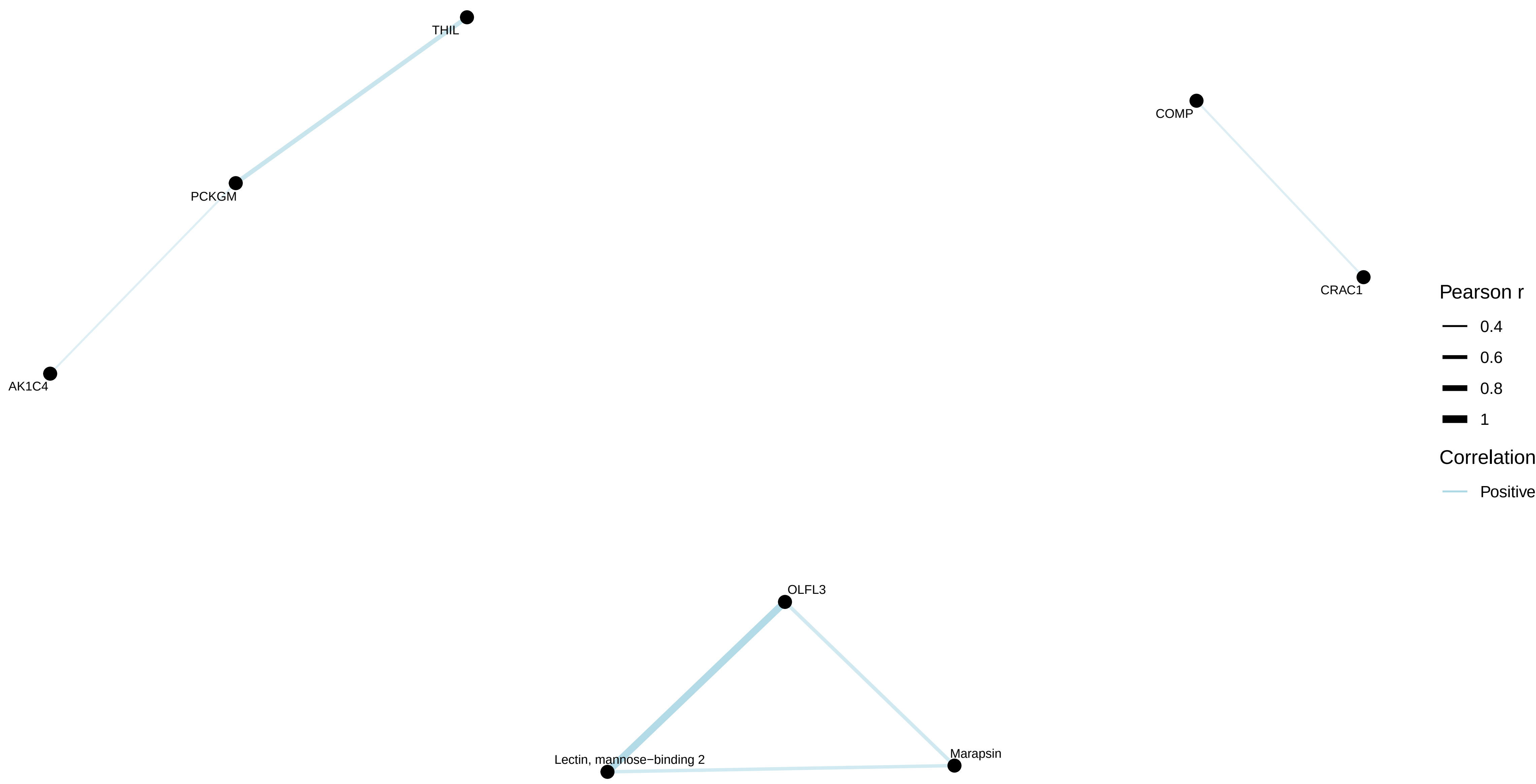


colorectal / female − cases

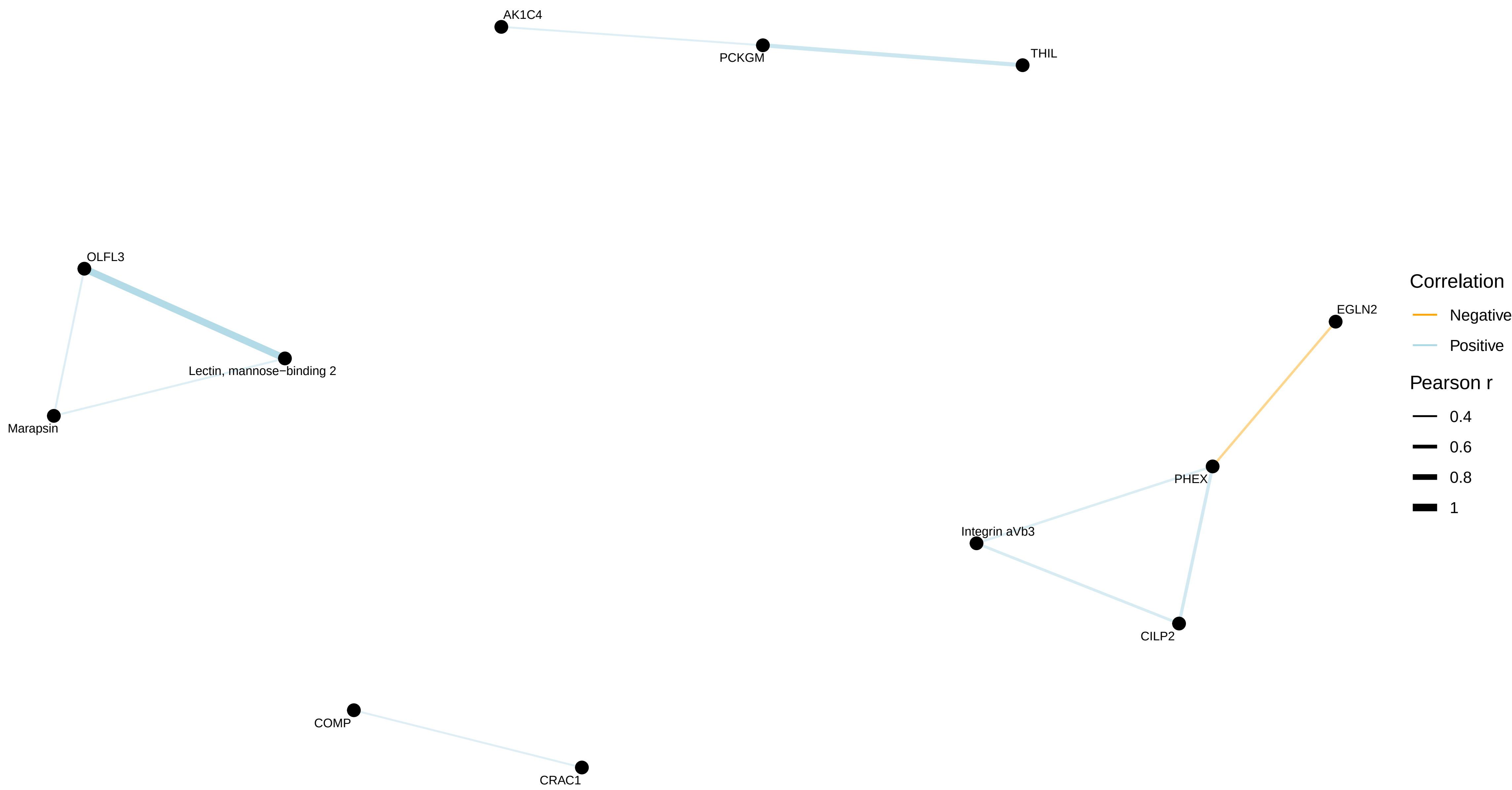


colorectal / female − non−cases

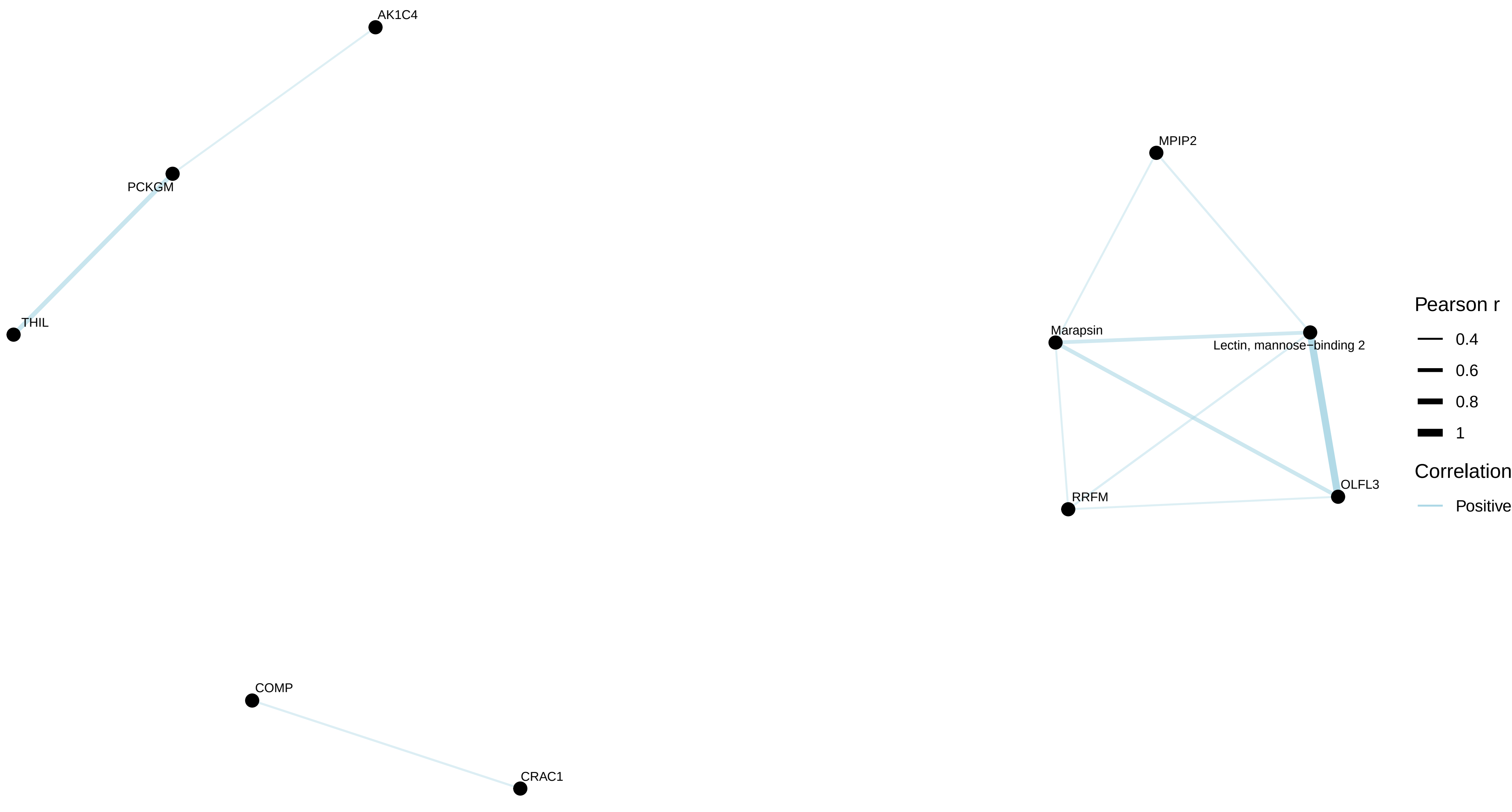

colorectal / male − all
No correlations with |r| > 0.4

colorectal / male − cases
No correlations with |r| > 0.4

colorectal / male − non−cases

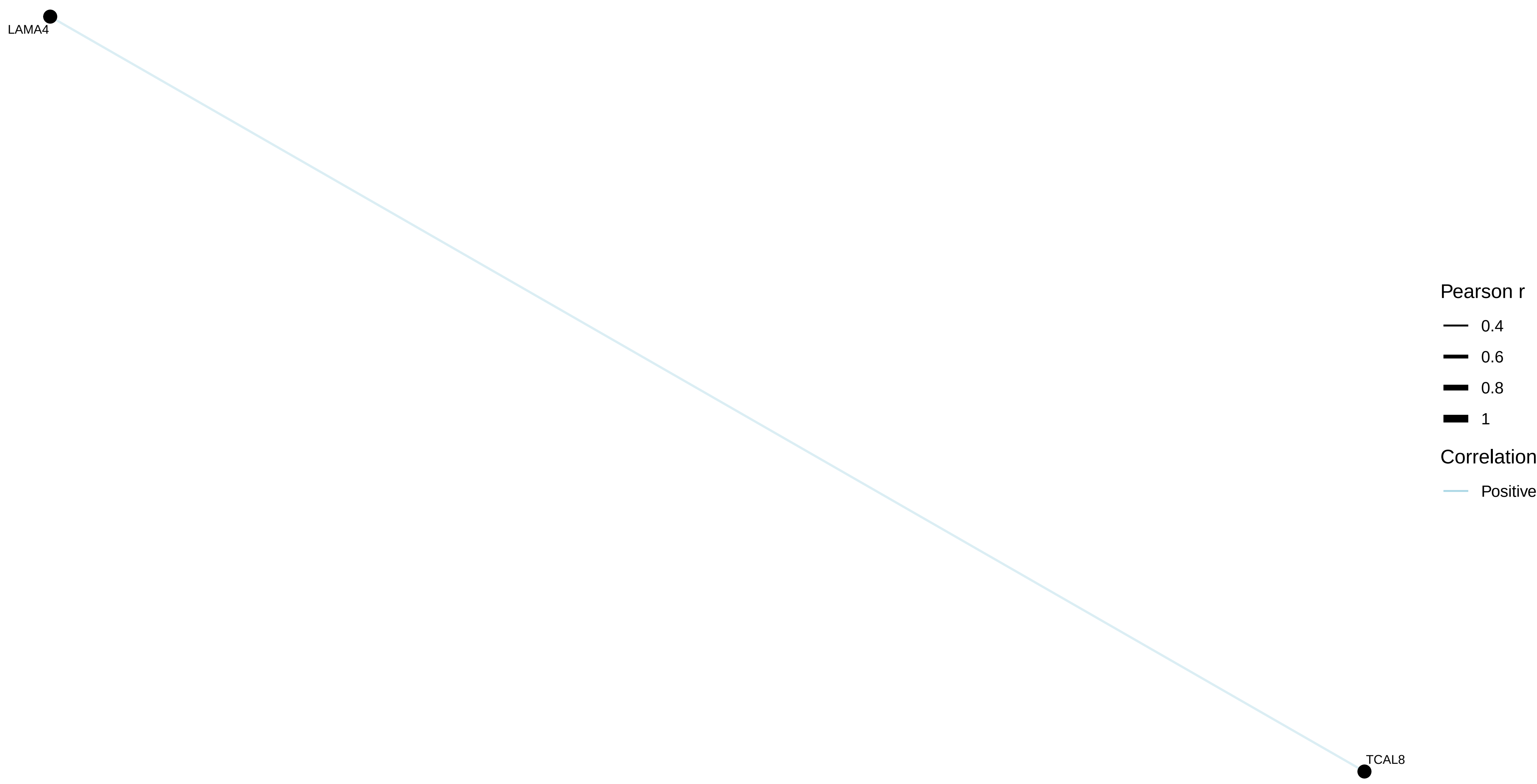

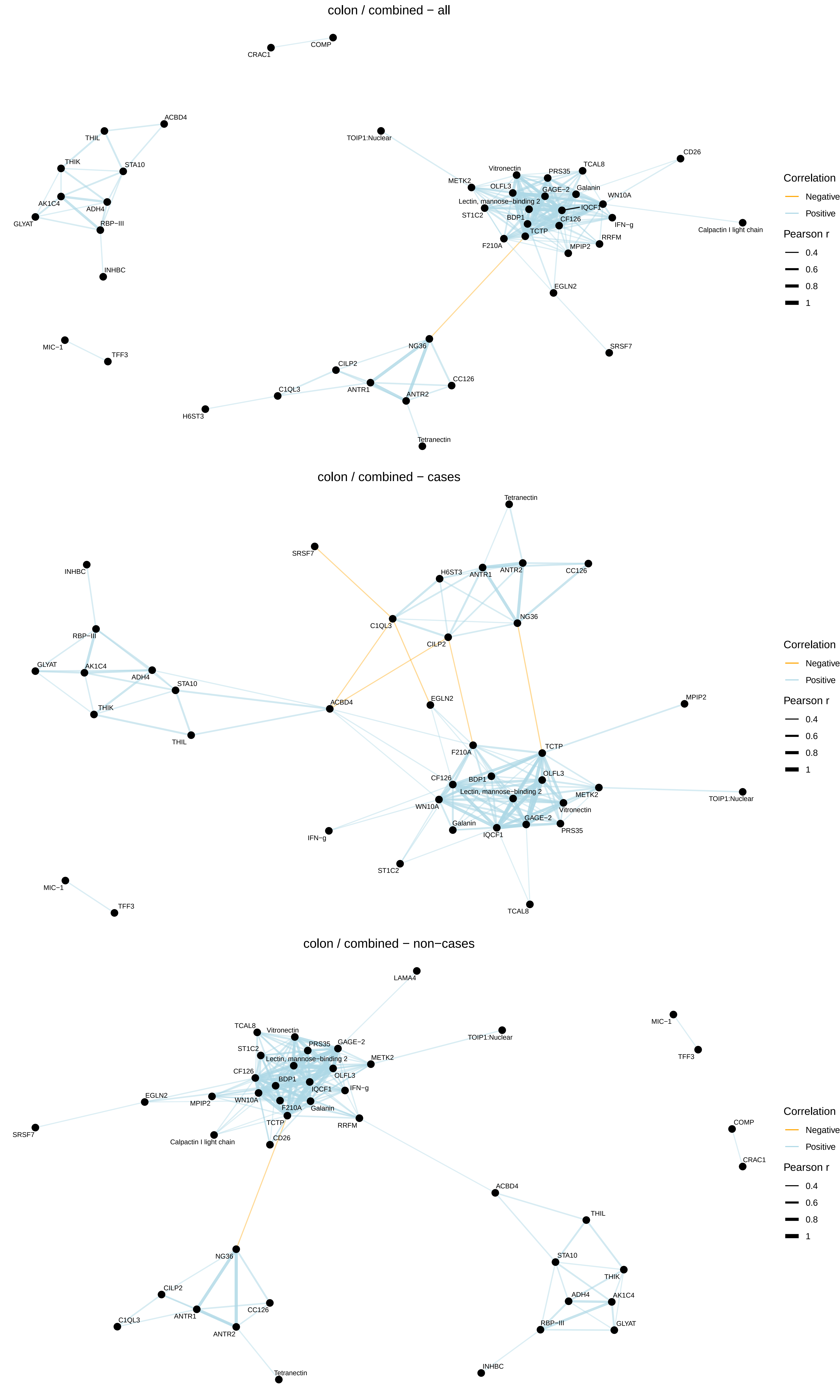
colon / combined − all
CRAC1
COMP
ACBD4
THIL
THIK
STA10
AK1C4
ADH4
GLYAT
RBP−III
INHBC
MIC−1
TFF3
TOIP1:Nuclear
CD26
Vitronectin
PRS35
TCAL8
METK2
OLFL3
GAGE−2
Galanin
WN10A
Lectin, mannose−binding 2
IQCF1
ST1C2
BDP1
CF126
IFN−g
Calpactin I light chain
TCTP
RRFM
F210A
MPIP2
EGLN2
SRSF7
NG36
CILP2
C1QL3
ANTR1
CC126
ANTR2
H6ST3
Tetranectin
Correlation
Negative
Positive
Pearson r
0.4
0.6
0.8
1
colon / combined − cases
Tetranectin
SRSF7
INHBC
H6ST3
ANTR1
ANTR2
CC126
NG36
C1QL3
CILP2
RBP−III
GLYAT
AK1C4
ADH4
STA10
ACBD4
EGLN2
THIK
THIL
MPIP2
F210A
TCTP
OLFL3
CF126
BDP1
Lectin, mannose−binding 2
METK2
TOIP1:Nuclear
WN10A
Vitronectin
GAGE−2
Galanin
IQCF1
PRS35
IFN−g
ST1C2
MIC−1
TFF3
TCAL8
Correlation
Negative
Positive
Pearson r
0.4
0.6
0.8
1
colon / combined − non−cases
LAMA4
TCAL8
Vitronectin
TOIP1:Nuclear
MIC−1
PRS35
GAGE−2
ST1C2
Lectin, mannose−binding 2
METK2
TFF3
CF126
OLFL3
BDP1
IQCF1
IFN−g
EGLN2
MPIP2
WN10A
F210A
Galanin
TCTP
RRFM
SRSF7
Calpactin I light chain
CD26
COMP
CRAC1
ACBD4
THIL
STA10
THIK
NG36
ADH4
AK1C4
CILP2
CC126
RBP−III
GLYAT
C1QL3
ANTR1
ANTR2
INHBC
Tetranectin
Correlation
Negative
Positive
Pearson r
0.4
0.6
0.8
1

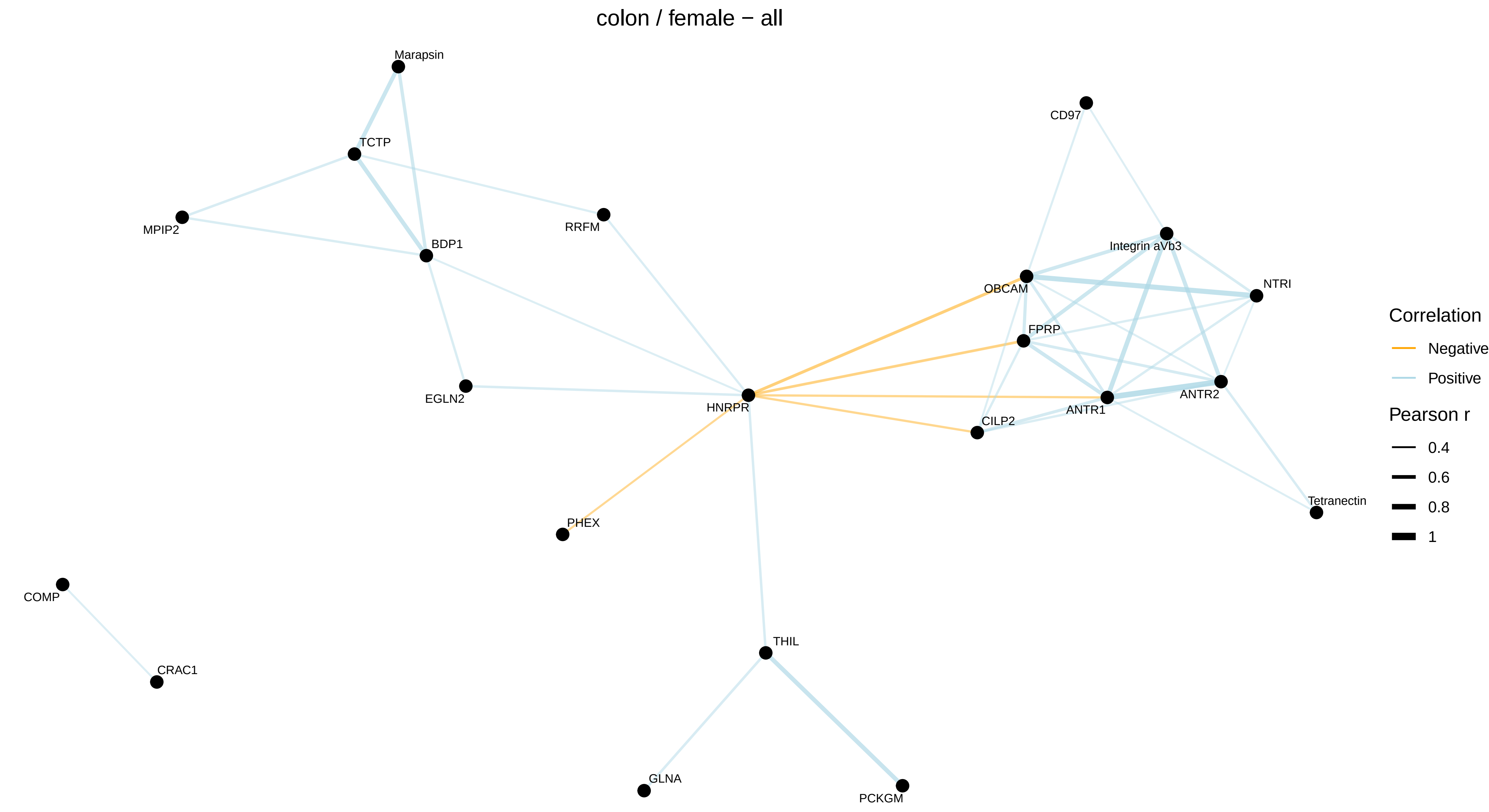
colon / female − all
Marapsin
TCTP
MPIP2
RRFM
BDP1
EGLN2
HNRPR
PHEX
CD97
Integrin aVb3
OBCAM
NTRI
FPRP
ANTR1
ANTR2
CILP2
Tetranectin
COMP
CRAC1
THIL
GLNA
PCKGM
Correlation
Negative
Positive
Pearson r
0.4
0.6
0.8
1

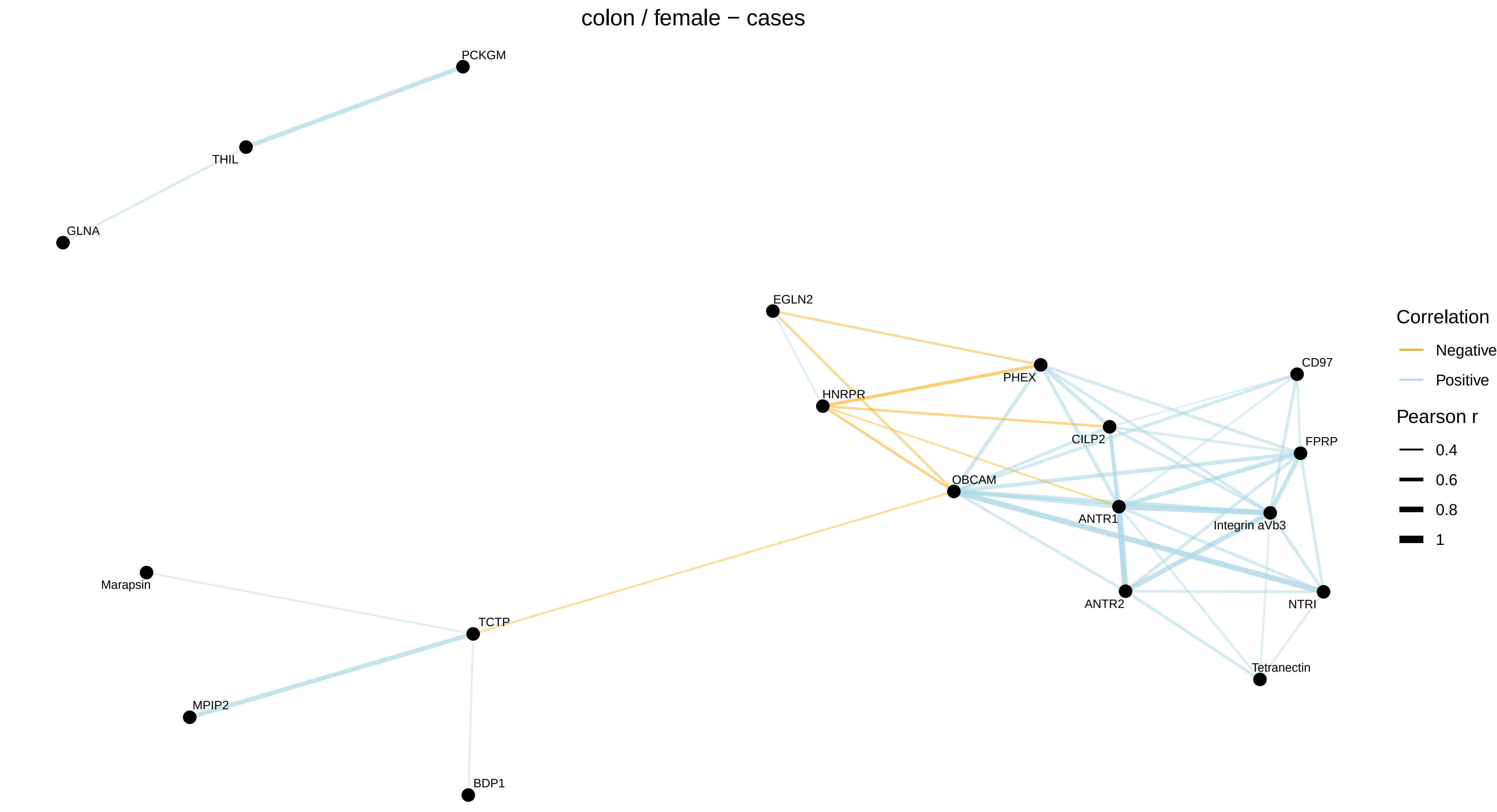
colon / female − cases
PCKGM
THIL
GLNA
EGLN2
HNRPR
PHEX
CD97
CILP2
FPRP
OBCAM
ANTR1
Integrin aVb3
ANTR2
NTRI
Tetranectin
Marapsin
TCTP
MPIP2
BDP1
Correlation
Negative
Positive
Pearson r
0.4
0.6
0.8
1

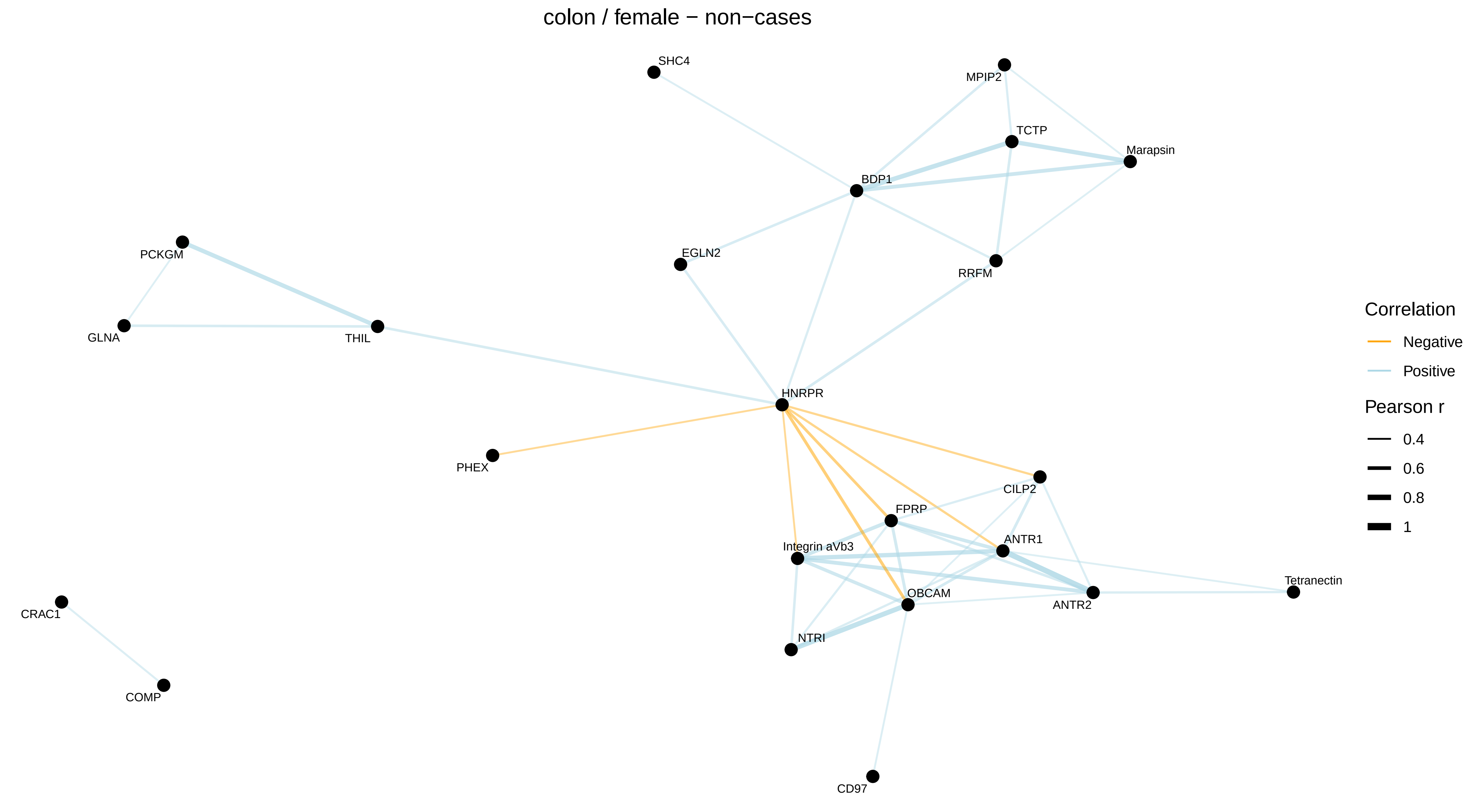
colon / female − non−cases
SHC4
MPIP2
TCTP
Marapsin
BDP1
EGLN2
RRFM
PCKGM
GLNA
THIL
HNRPR
PHEX
CILP2
FPRP
ANTR1
Integrin aVb3
OBCAM
ANTR2
Tetranectin
NTRI
CRAC1
COMP
CD97
Correlation
Negative
Positive
Pearson r
0.4
0.6
0.8
1

colon / male − all

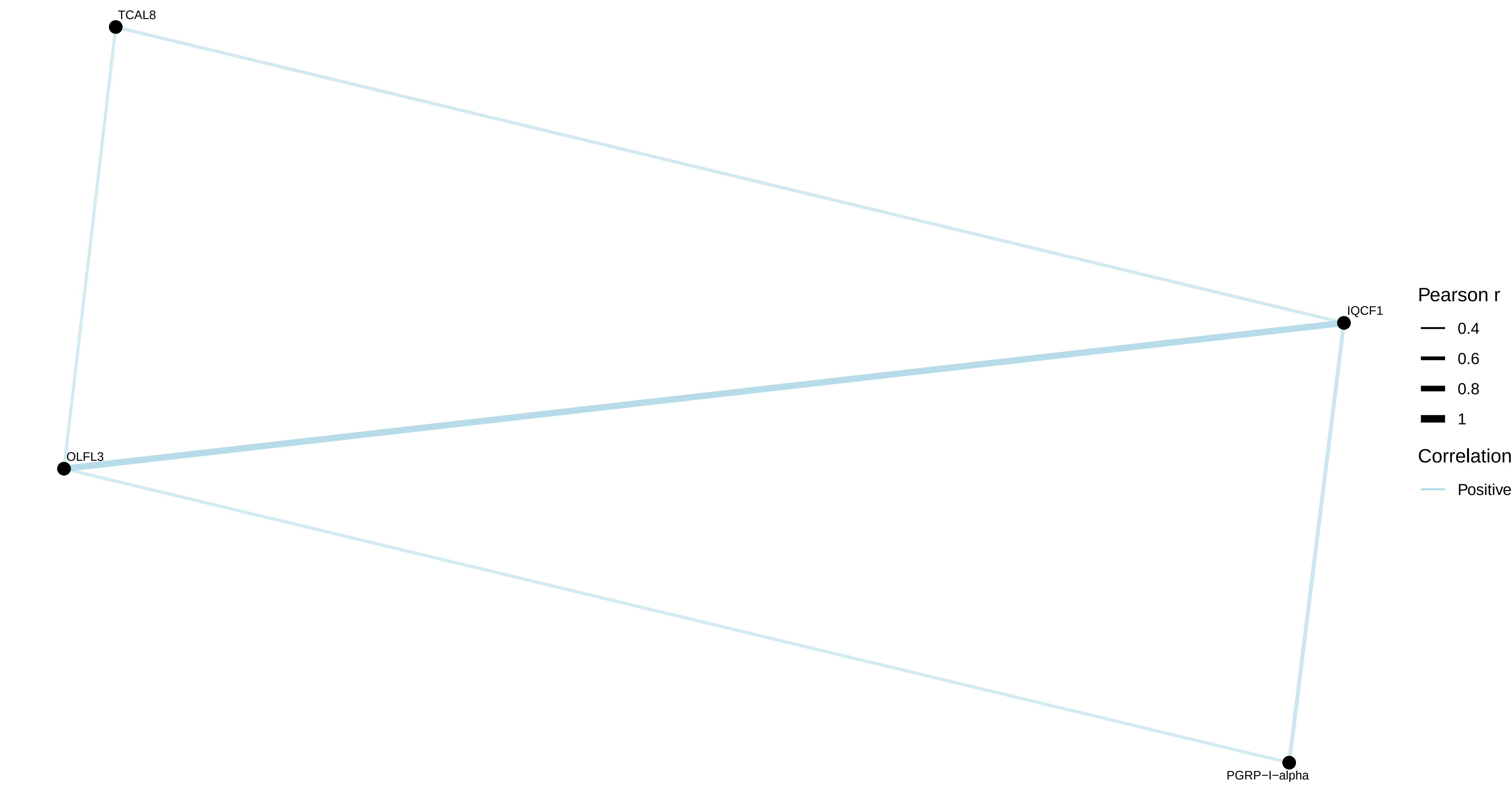


colon / male − cases

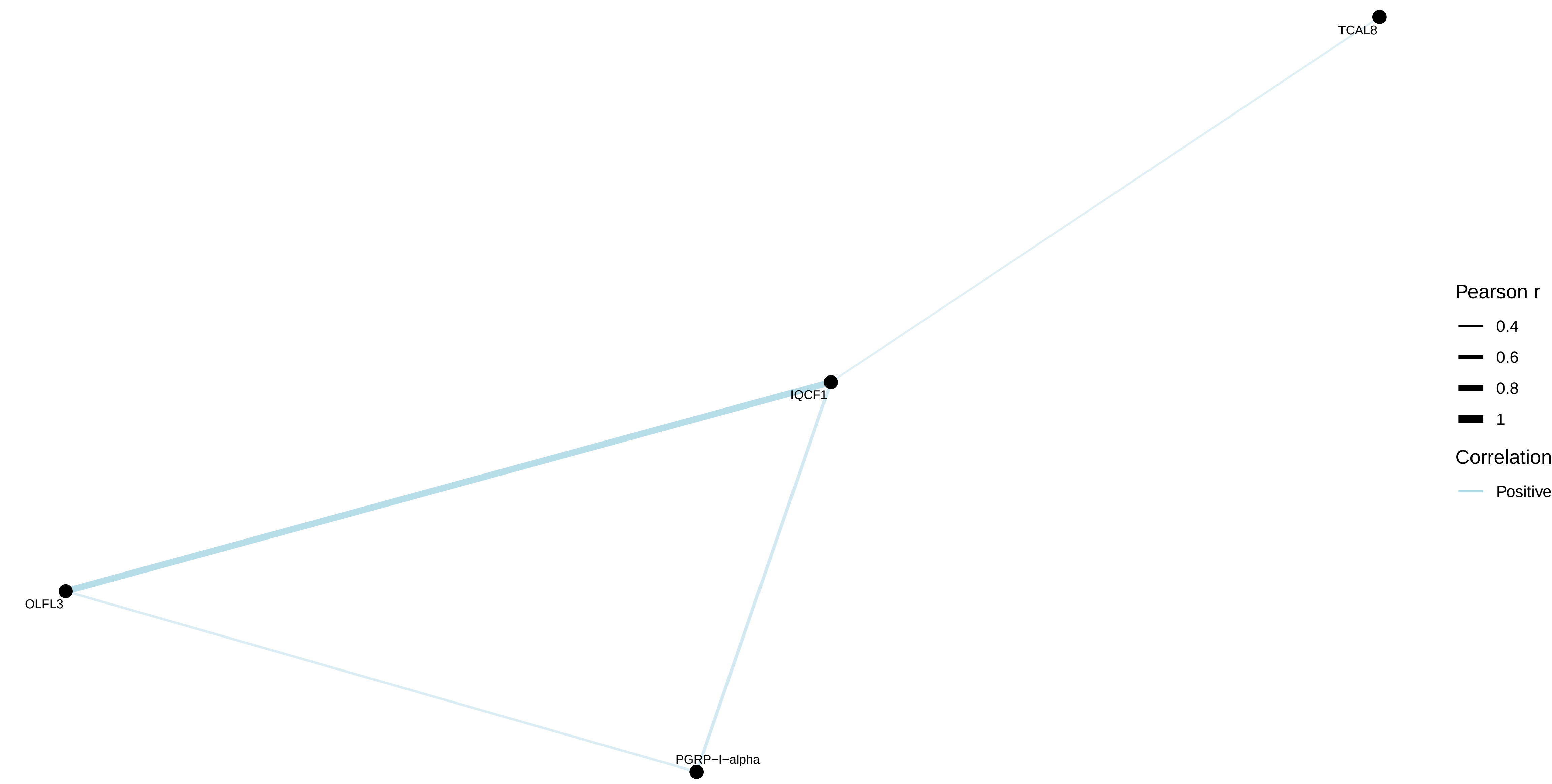


colon / male − non−cases

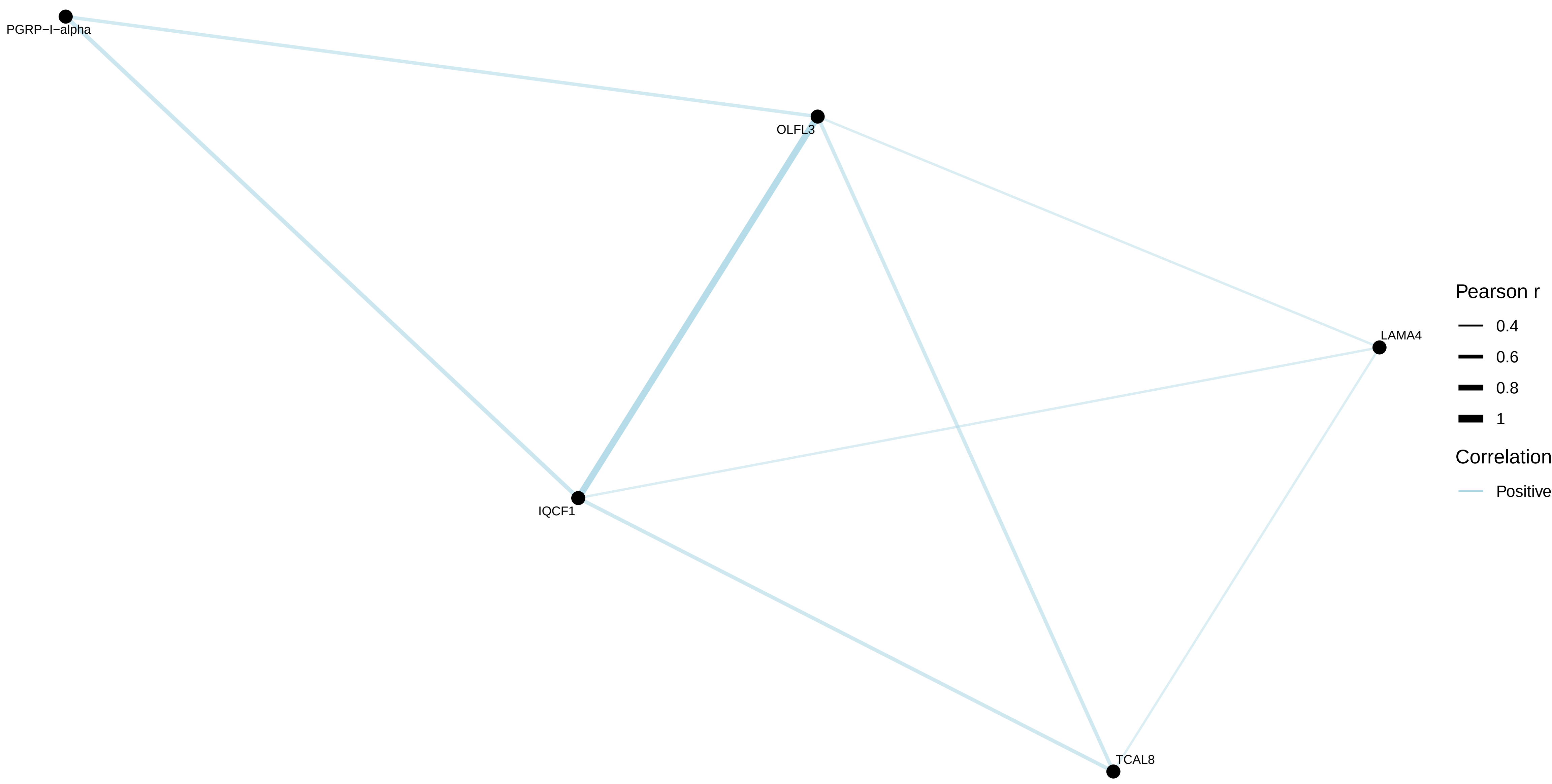

rectum / combined − all
No correlations with |r| > 0.4

rectum / combined − cases
No correlations with |r| > 0.4

rectum / combined − non−cases
No correlations with |r| > 0.4

rectum / male − all
No correlations with |r| > 0.4

rectum / male − cases
No correlations with |r| > 0.4

rectum / male − non−cases
No correlations with |r| > 0.4

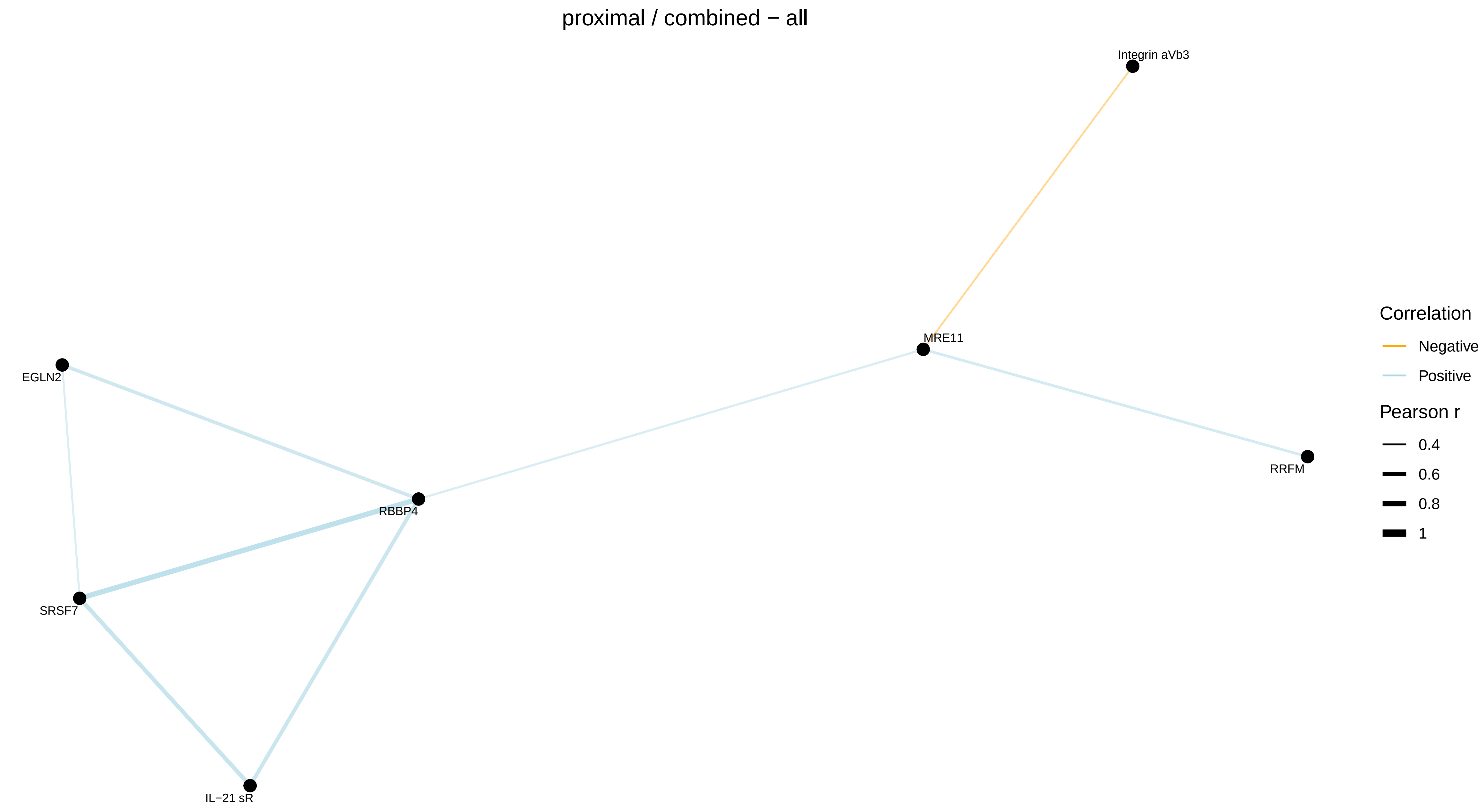
proximal / combined − all
Integrin aVb3
MRE11
EGLN2
RRFM
RBBP4
SRSF7
IL−21 sR
Correlation
Negative
Positive
Pearson r
0.4
0.6
0.8
1

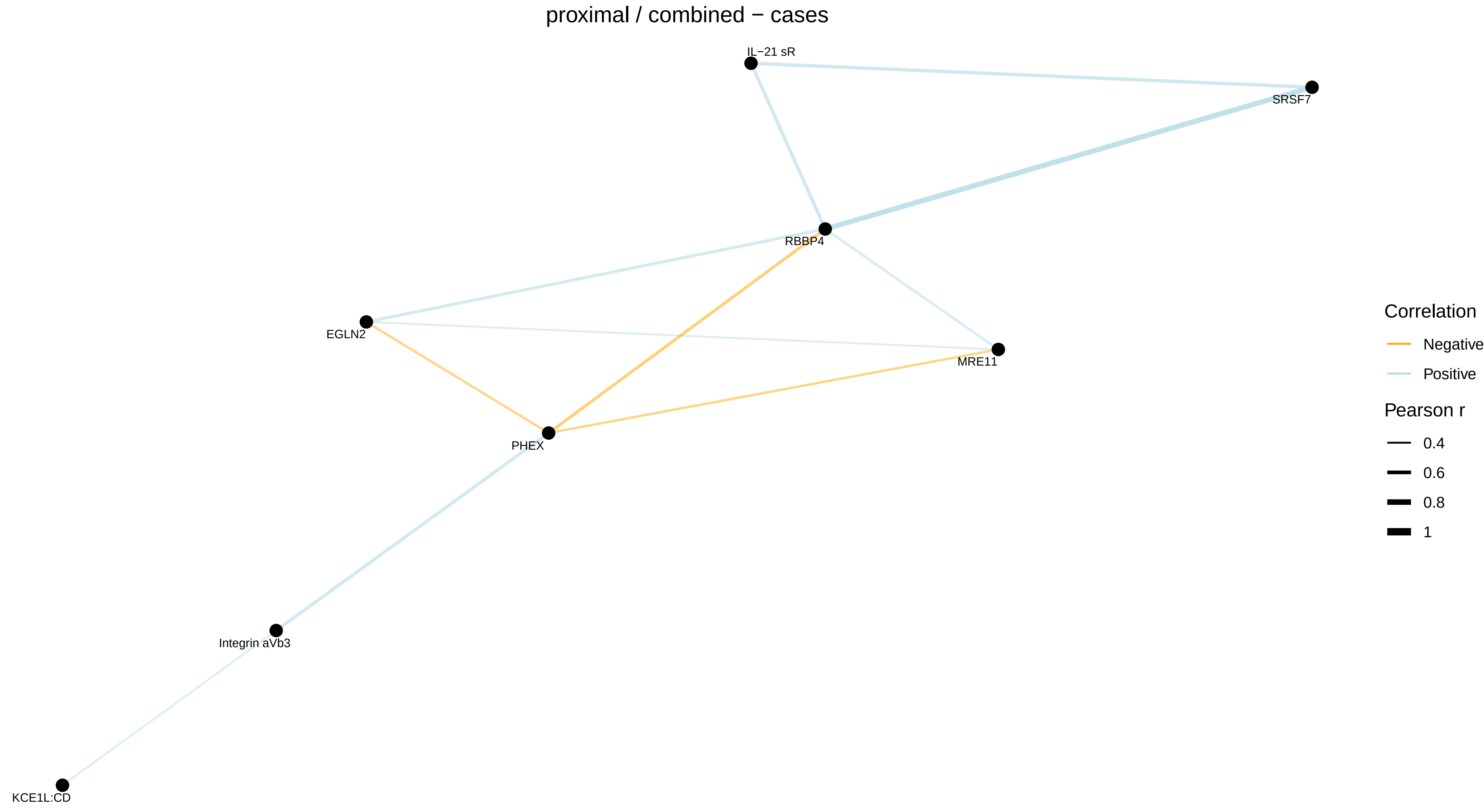
proximal / combined − cases
IL−21 sR
SRSF7
RBBP4
EGLN2
MRE11
PHEX
Integrin aVb3
KCE1L:CD
Correlation
Negative
Positive
Pearson r
0.4
0.6
0.8
1

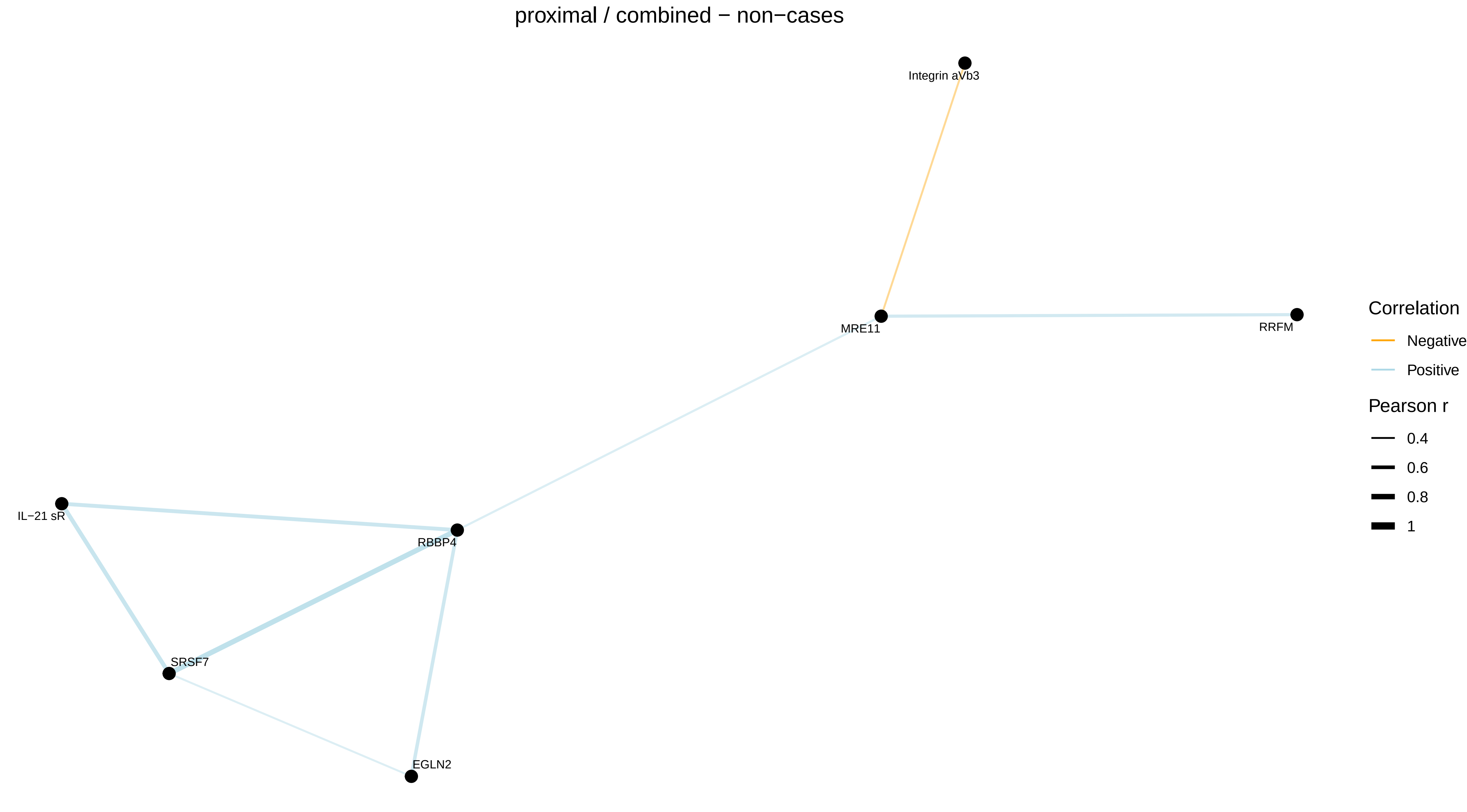
proximal / combined − non−cases
Integrin aVb3
MRE11
RRFM
IL−21 sR
RBBP4
SRSF7
EGLN2
Correlation
Negative
Positive
Pearson r
0.4
0.6
0.8
1

proximal / female − all

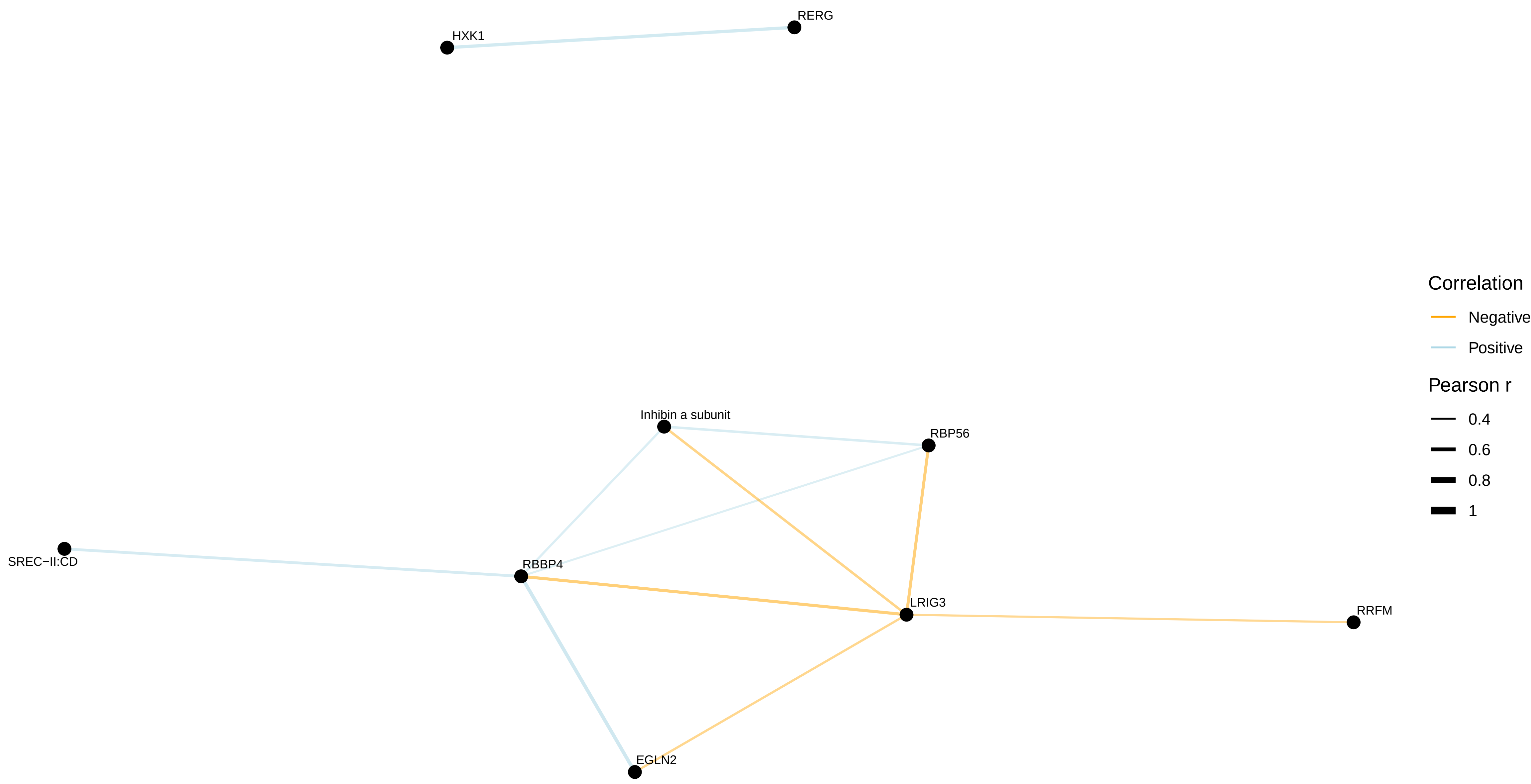


proximal / female − cases

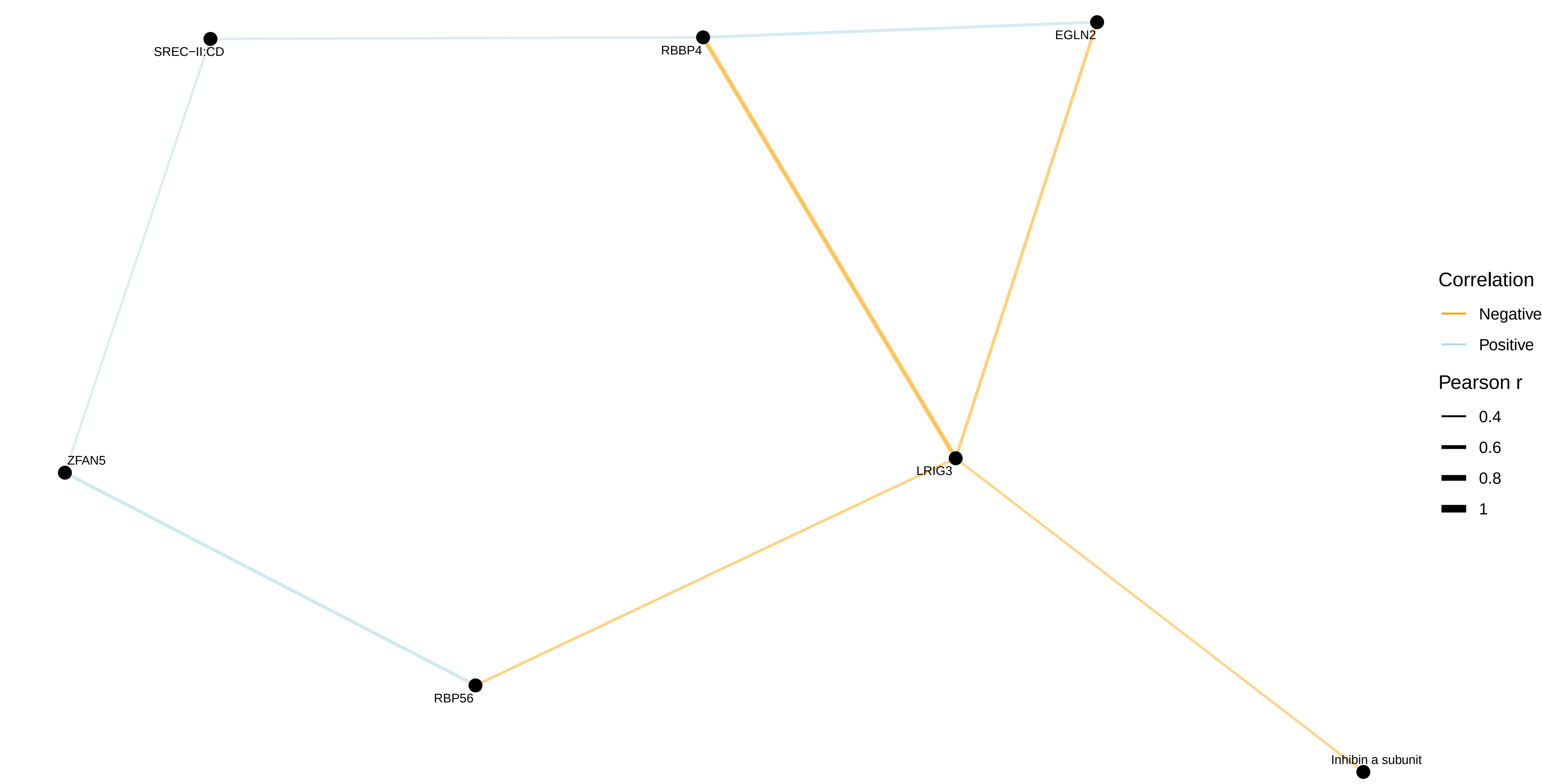


proximal / female − non−cases

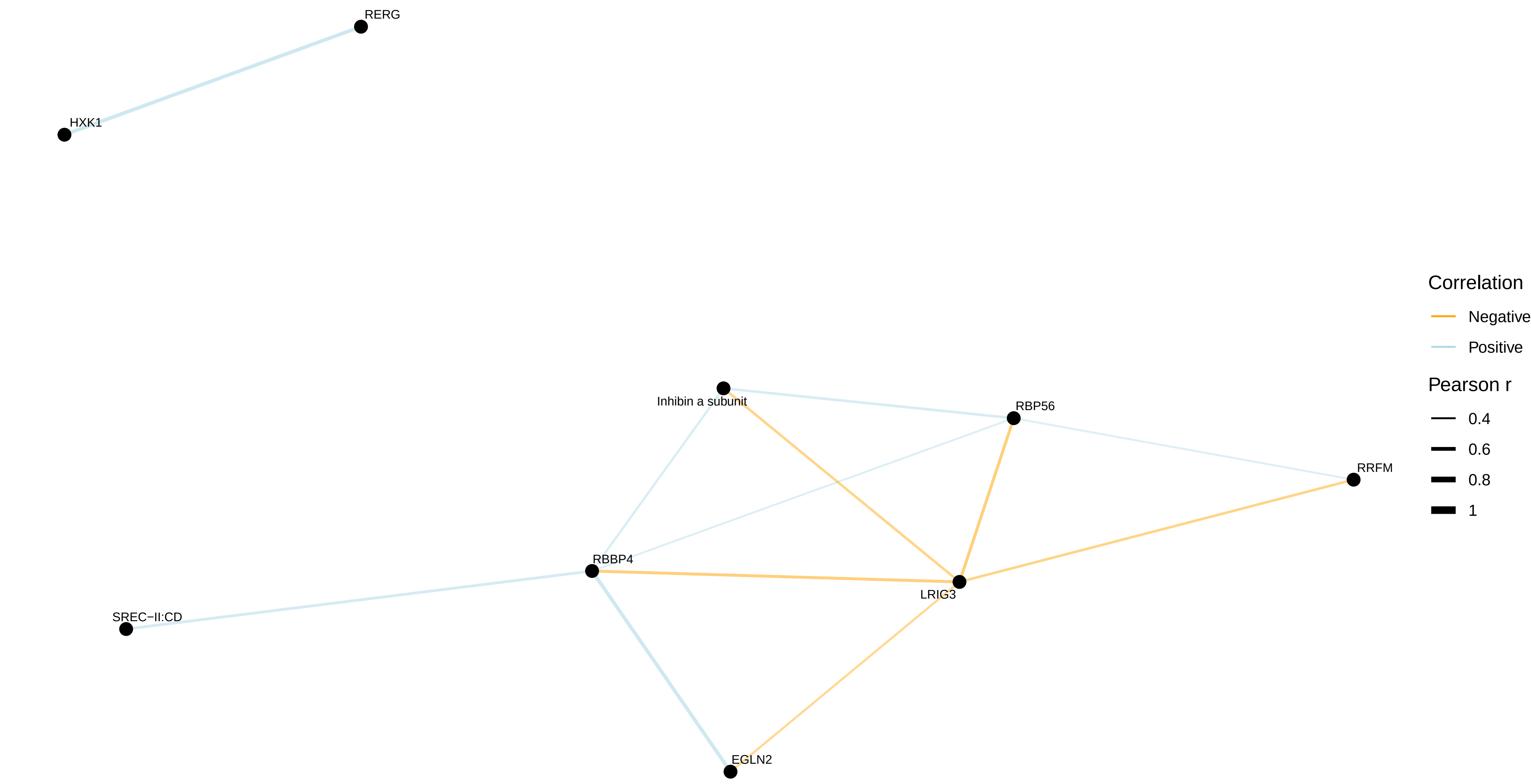

distal / combined − all
No correlations with |r| > 0.4

distal / combined − cases

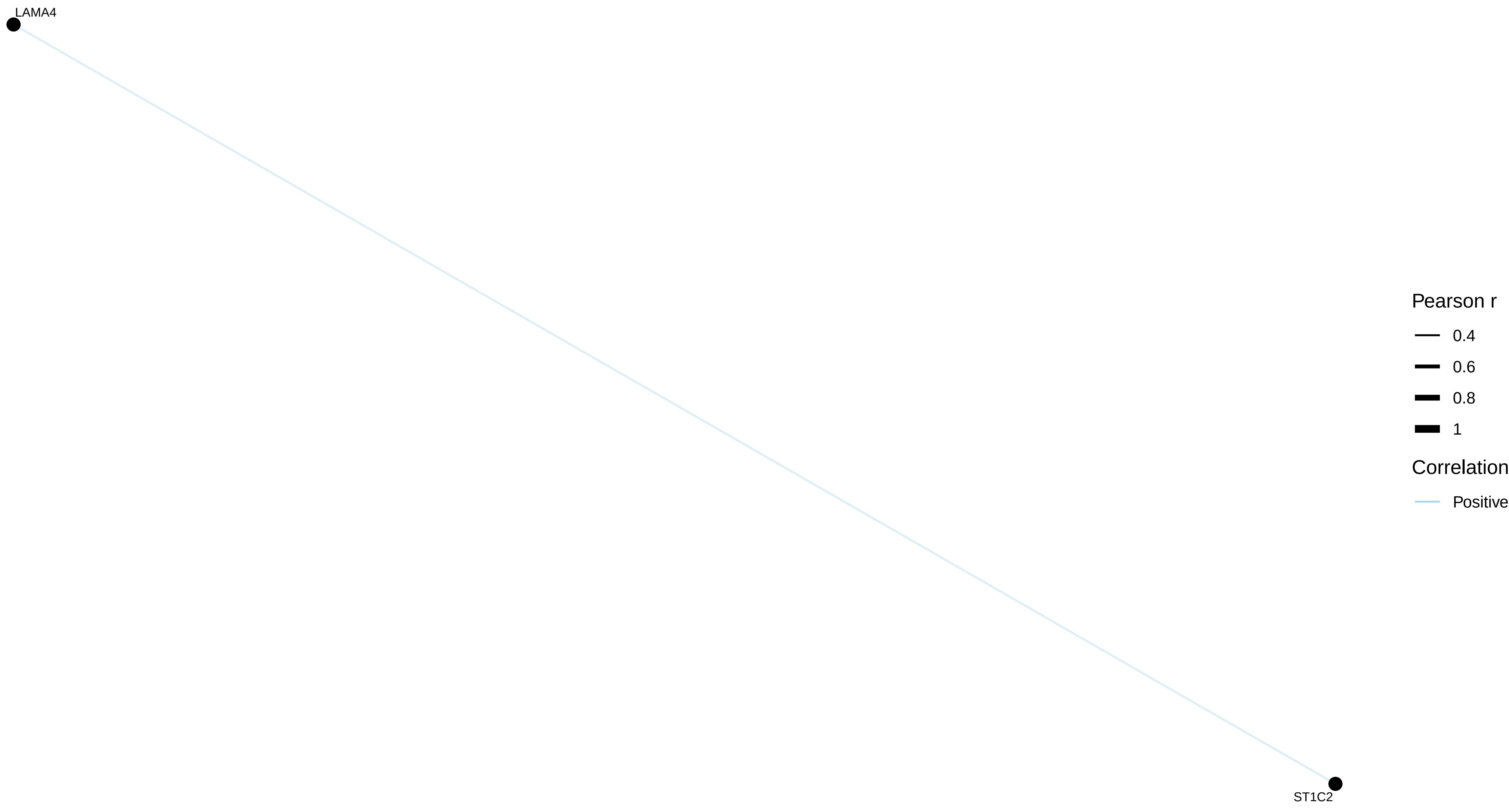


distal / combined − non−cases
No correlations with |r| > 0.4

distal / female − all
No correlations with |r| > 0.4

distal / female − cases

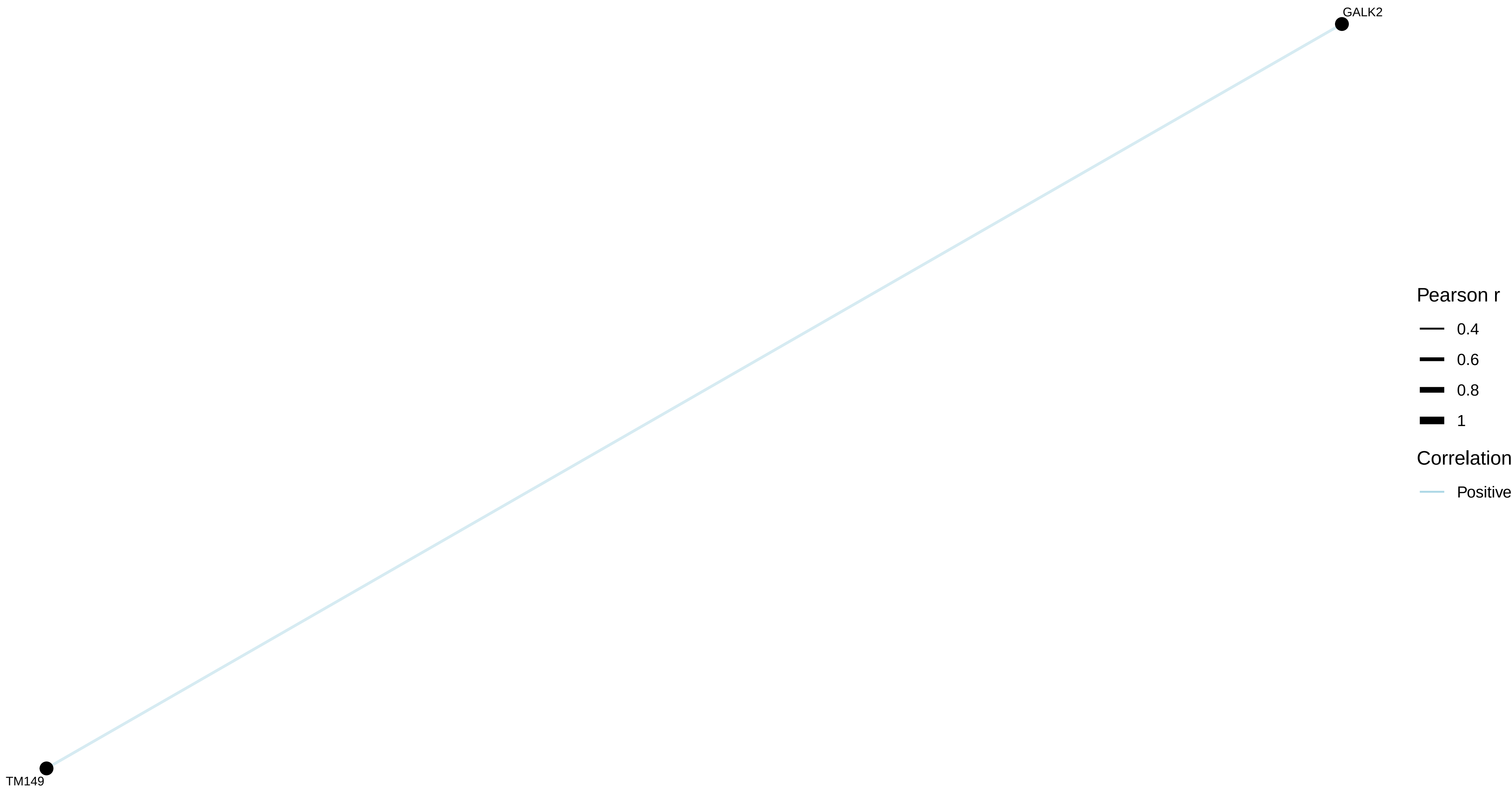


distal / female − non−cases
No correlations with |r| > 0.4

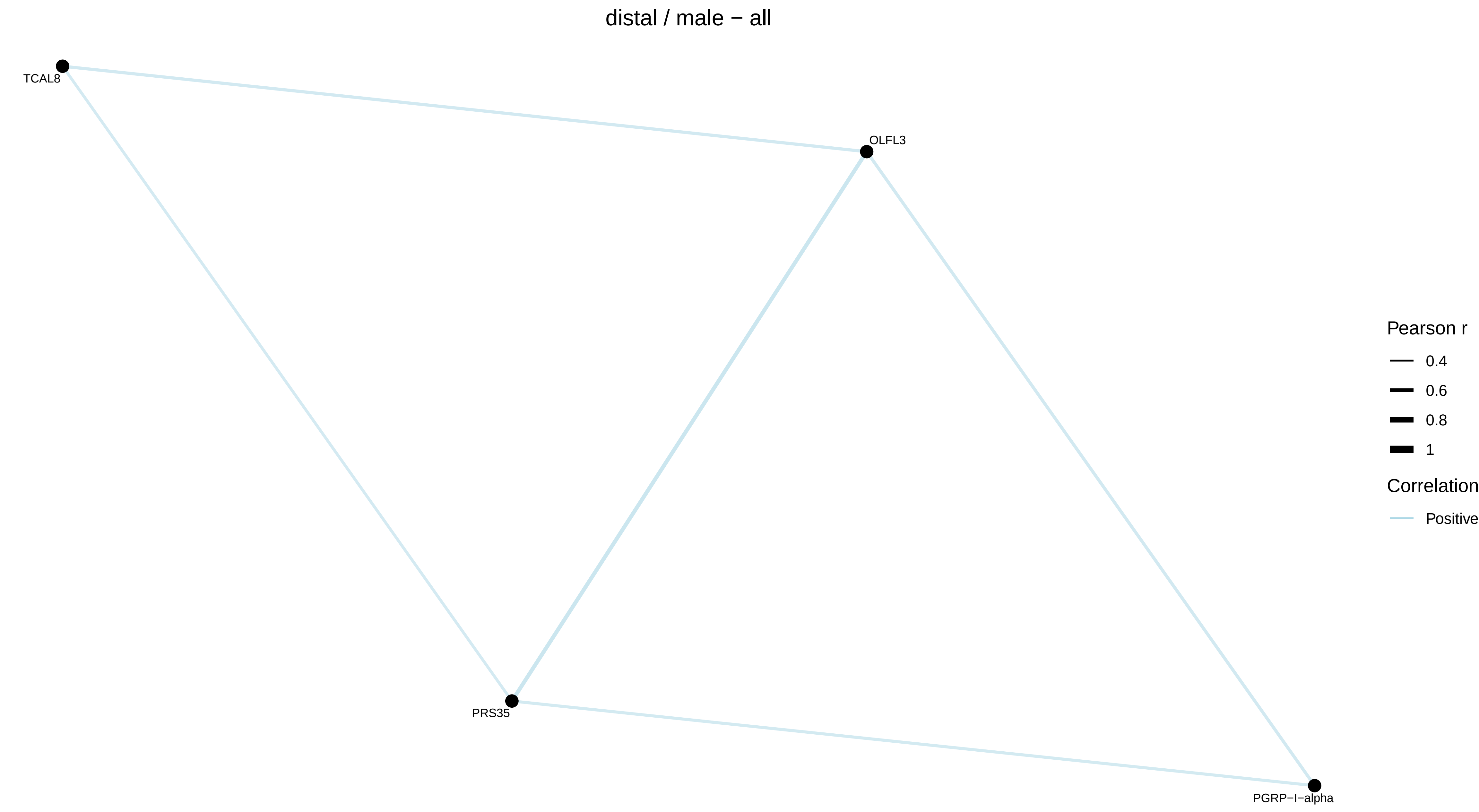
distal / male − all
TCAL8
OLFL3
PRS35
PGRP−I−alpha
Pearson r
0.4
0.6
0.8
1
Correlation
Positive

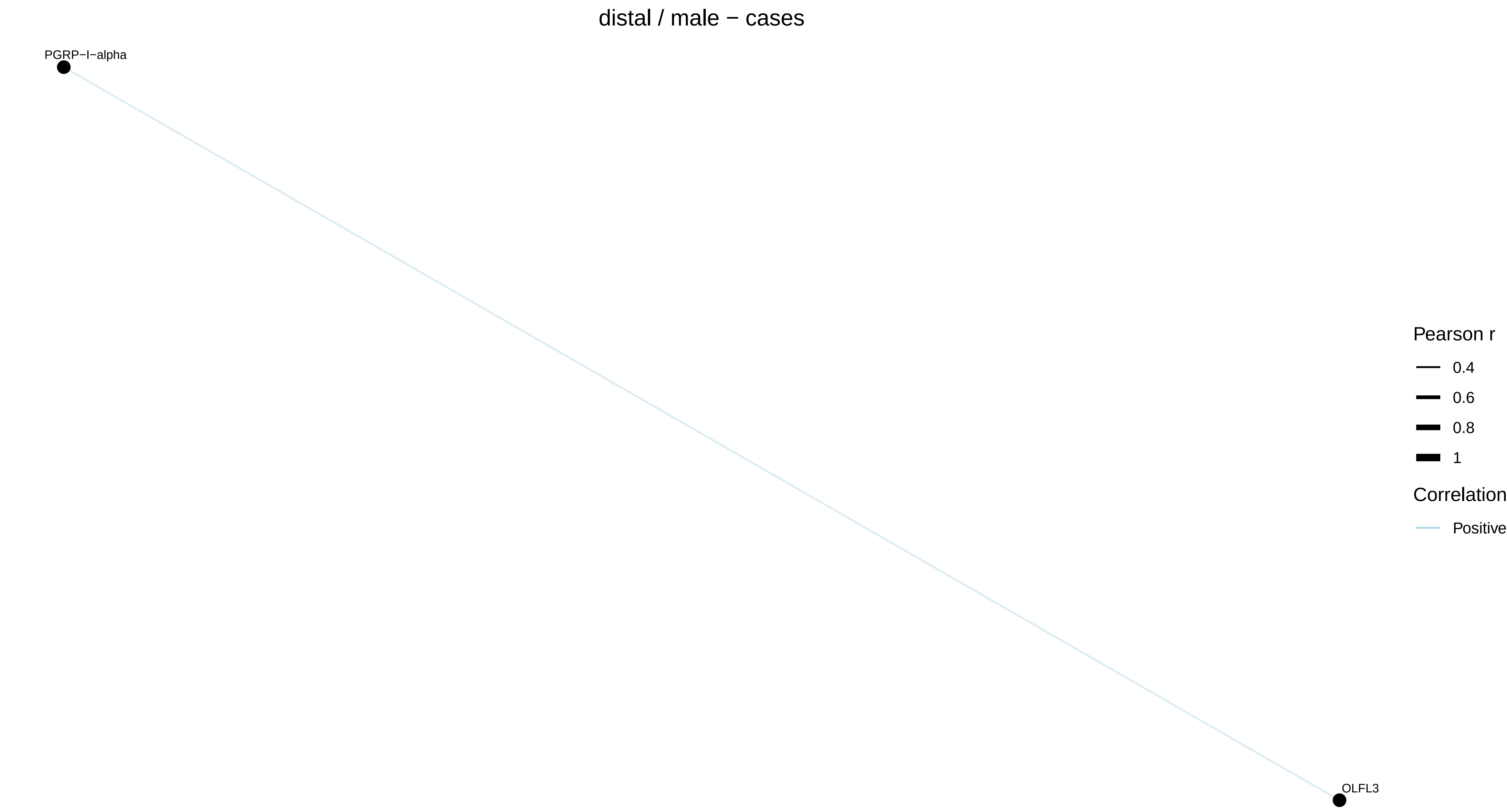
distal / male − cases
PGRP−I−alpha
OLFL3
Pearson r
0.4
0.6
0.8
1
Correlation
Positive

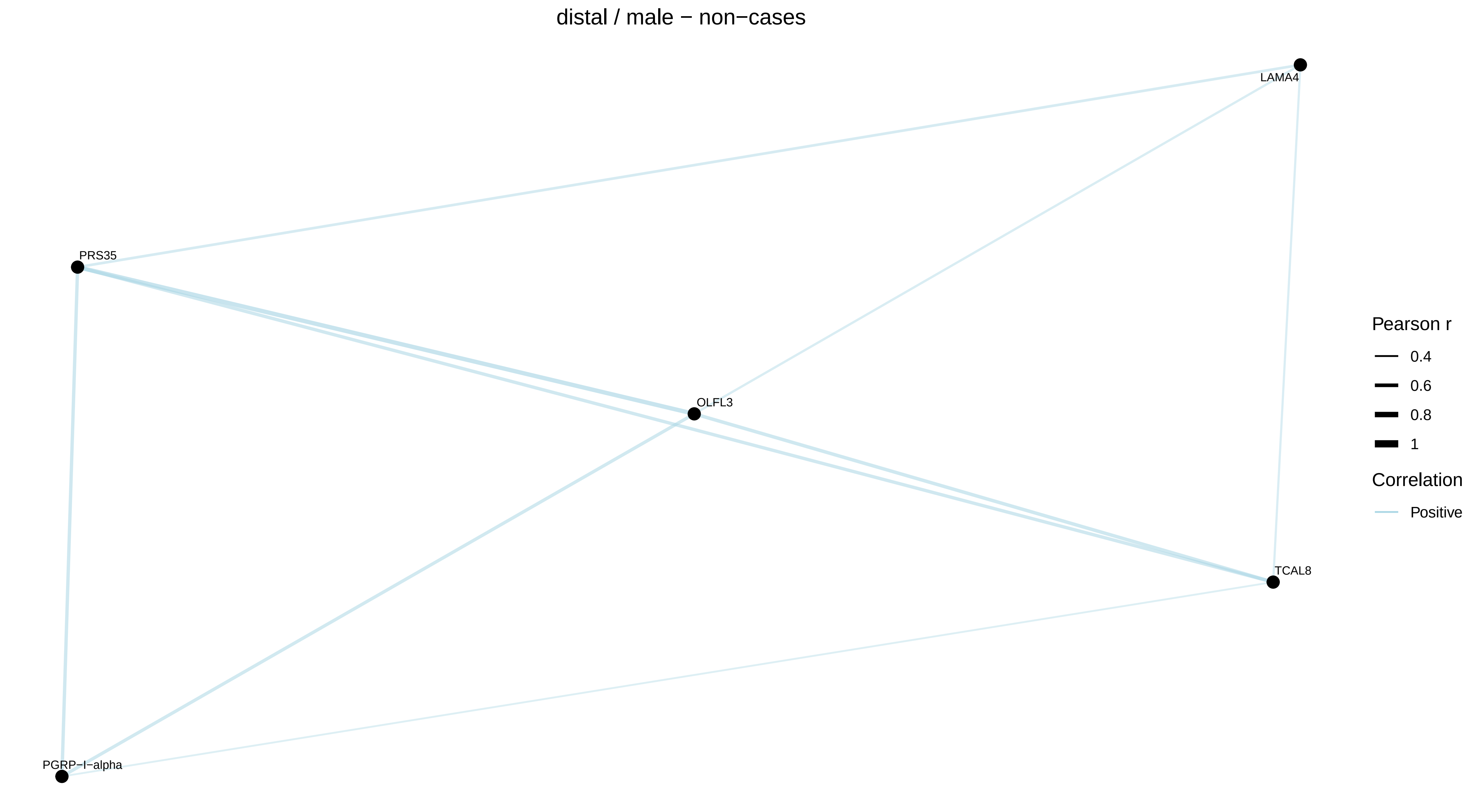
distal / male − non−cases
LAMA4
PRS35
OLFL3
TCAL8
PGRP−I−alpha
Pearson r
0.4
0.6
0.8
1
Correlation
Positive
