## Supplementary Figure 4 for "Proteomic markers of colorectal cancer risk: results from the European Prospective Investigation into Cancer and Nutrition"

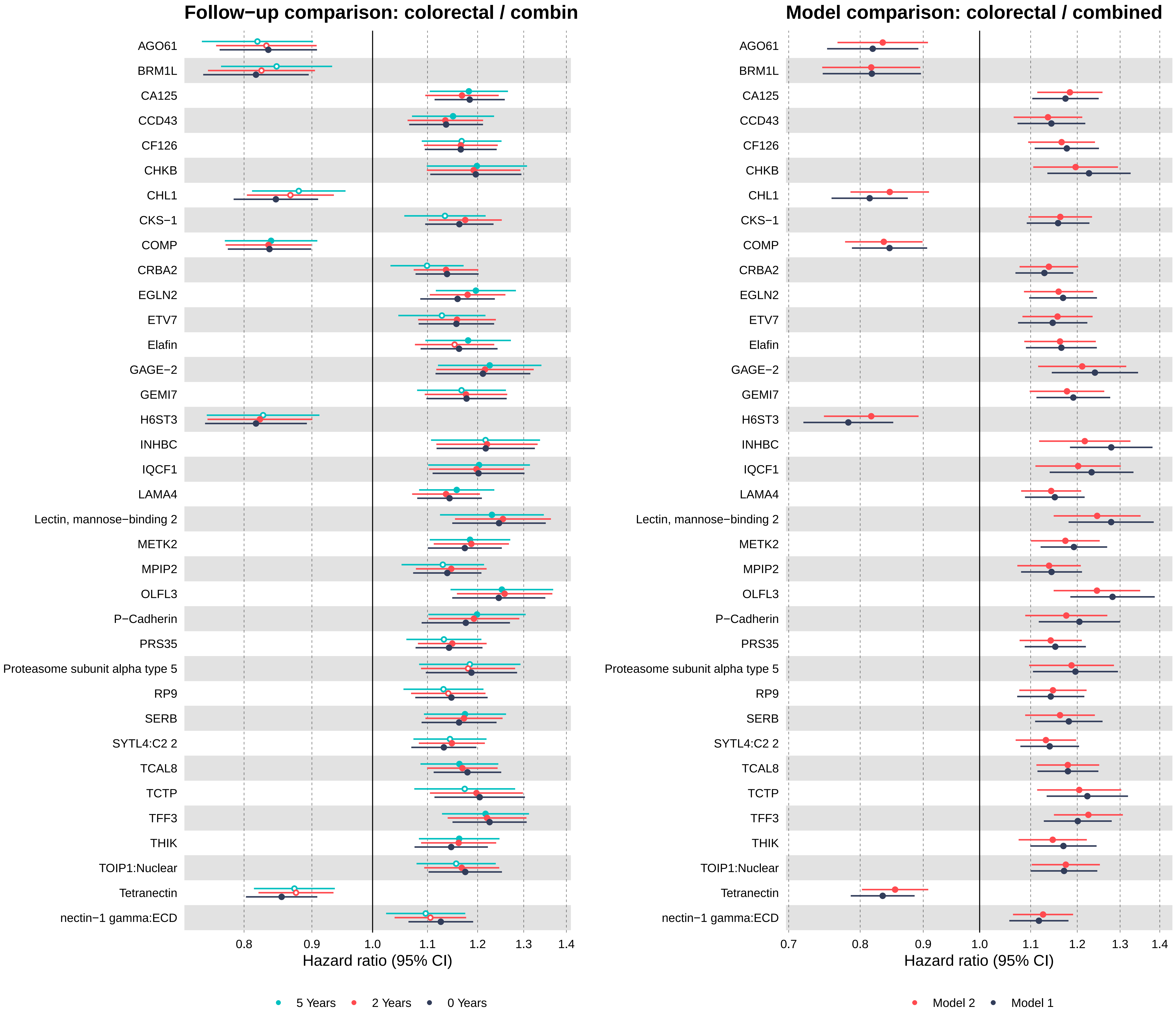
Follow−up comparison: colorectal / combin
AGO61
BRM1L
CA125
CCD43
CF126
CHKB
CHL1
CKS−1
COMP
CRBA2
EGLN2
ETV7
Elafin
GAGE−2
GEMI7
H6ST3
INHBC
IQCF1
LAMA4
Lectin, mannose−binding 2
METK2
MPIP2
OLFL3
P−Cadherin
PRS35
Proteasome subunit alpha type 5
RP9
SERB
SYTL4:C2 2
TCAL8
TCTP
TFF3
THIK
TOIP1:Nuclear
Tetranectin
nectin−1 gamma:ECD
0.8
0.9
1.0
1.1
1.2
1.3
1.4
Hazard ratio (95% CI)
5 Years
2 Years
0 Years
Model comparison: colorectal / combined
AGO61
BRM1L
CA125
CCD43
CF126
CHKB
CHL1
CKS−1
COMP
CRBA2
EGLN2
ETV7
Elafin
GAGE−2
GEMI7
H6ST3
INHBC
IQCF1
LAMA4
Lectin, mannose−binding 2
METK2
MPIP2
OLFL3
P−Cadherin
PRS35
Proteasome subunit alpha type 5
RP9
SERB
SYTL4:C2 2
TCAL8
TCTP
TFF3
THIK
TOIP1:Nuclear
Tetranectin
nectin−1 gamma:ECD
0.7
0.8
0.9
1.0
1.1
1.2
1.3
1.4
Hazard ratio (95% CI)
Model 2
Model 1

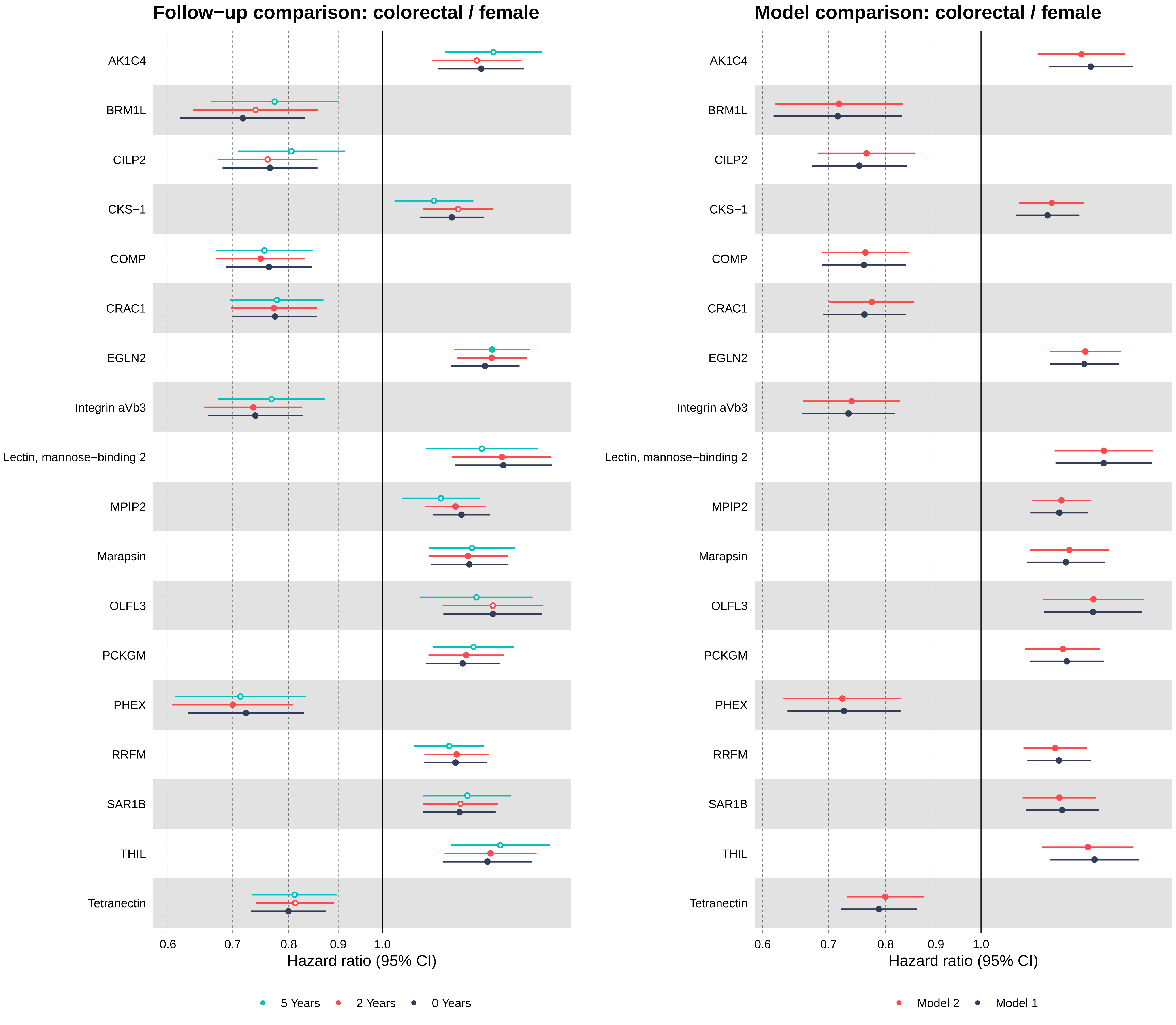
Follow−up comparison: colorectal / female
AK1C4
BRM1L
CILP2
CKS−1
COMP
CRAC1
EGLN2
Integrin aVb3
Lectin, mannose−binding 2
MPIP2
Marapsin
OLFL3
PCKGM
PHEX
RRFM
SAR1B
THIL
Tetranectin
0.6
0.7
0.8
0.9
1.0
Hazard ratio (95% CI)
5 Years
2 Years
0 Years
Model comparison: colorectal / female
AK1C4
BRM1L
CILP2
CKS−1
COMP
CRAC1
EGLN2
Integrin aVb3
Lectin, mannose−binding 2
MPIP2
Marapsin
OLFL3
PCKGM
PHEX
RRFM
SAR1B
THIL
Tetranectin
0.6
0.7
0.8
0.9
1.0
Hazard ratio (95% CI)
Model 2
Model 1

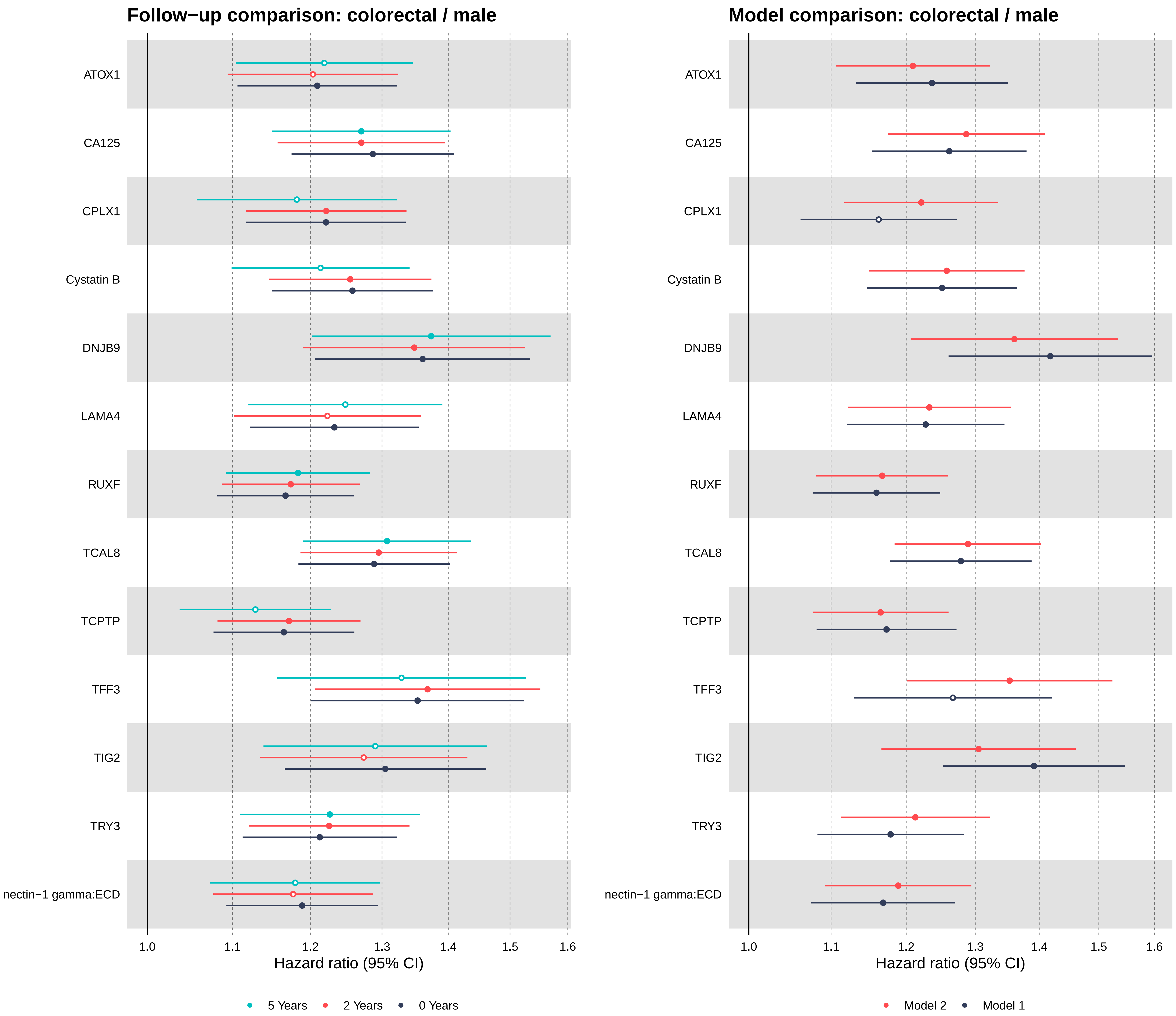

Follow−up comparison: colorectal / male
Model comparison: colorectal / male
ATOX1
CA125
CPLX1
Cystatin B
DNJB9
LAMA4
RUXF
TCAL8
TCPTP
TFF3
TIG2
TRY3
nectin−1 gamma:ECD
1.0
1.1
1.2
1.3
1.4
1.5
1.6
Hazard ratio (95% CI)
5 Years
2 Years
0 Years
Model 2
Model 1

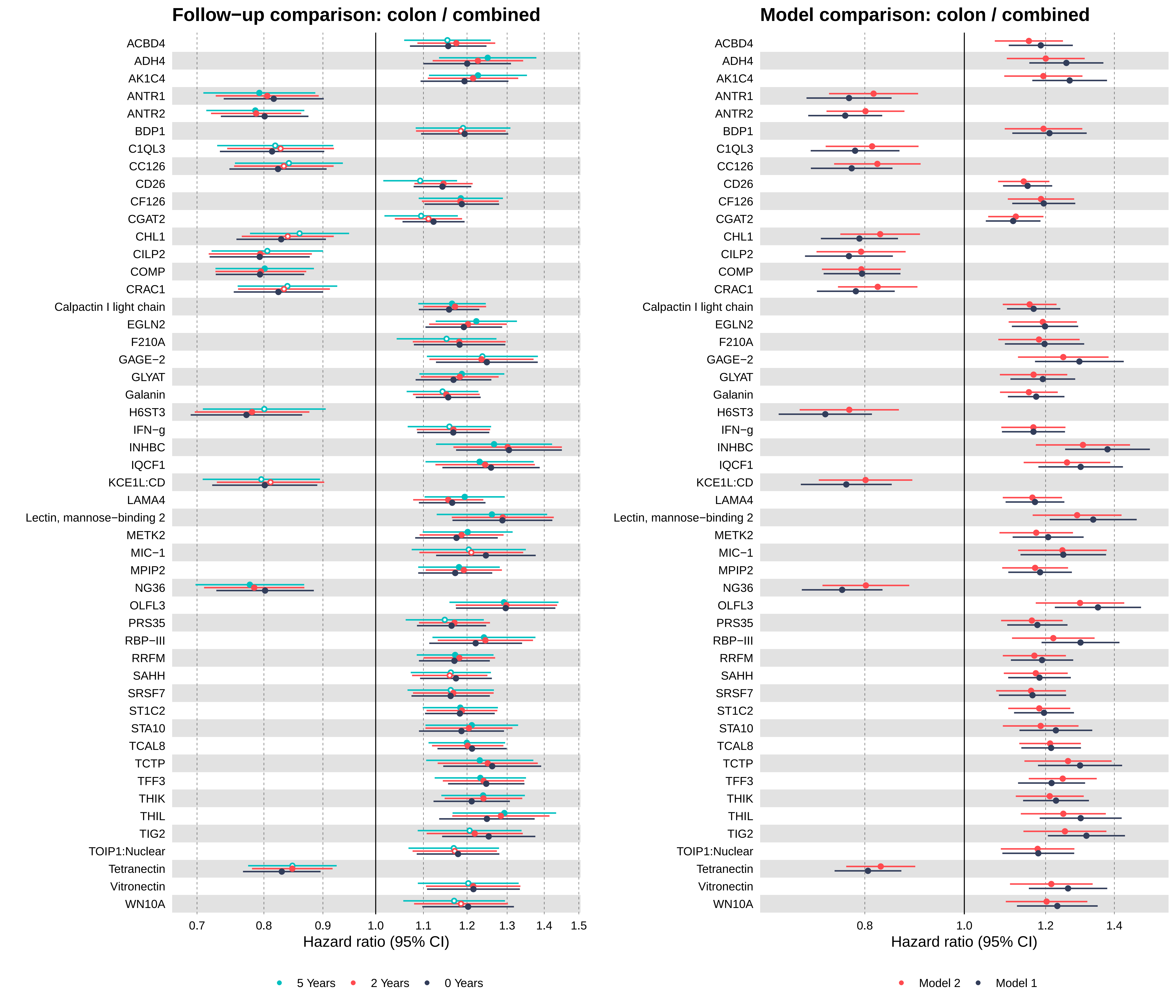

Follow−up comparison: colon / combined
Model comparison: colon / combined
ACBD4
ADH4
AK1C4
ANTR1
ANTR2
BDP1
C1QL3
CC126
CD26
CF126
CGAT2
CHL1
CILP2
COMP
CRAC1
Calpactin I light chain
EGLN2
F210A
GAGE−2
GLYAT
Galanin
H6ST3
IFN−g
INHBC
IQCF1
KCE1L:CD
LAMA4
Lectin, mannose−binding 2
METK2
MIC−1
MPIP2
NG36
OLFL3
PRS35
RBP−III
RRFM
SAHH
SRSF7
ST1C2
STA10
TCAL8
TCTP
TFF3
THIK
THIL
TIG2
TOIP1:Nuclear
Tetranectin
Vitronectin
WN10A
0.7
0.8
0.9
1.0
1.1
1.2
1.3
1.4
1.5
Hazard ratio (95% CI)
5 Years
2 Years
0 Years
0.8
1.0
1.2
1.4
Hazard ratio (95% CI)
Model 2
Model 1

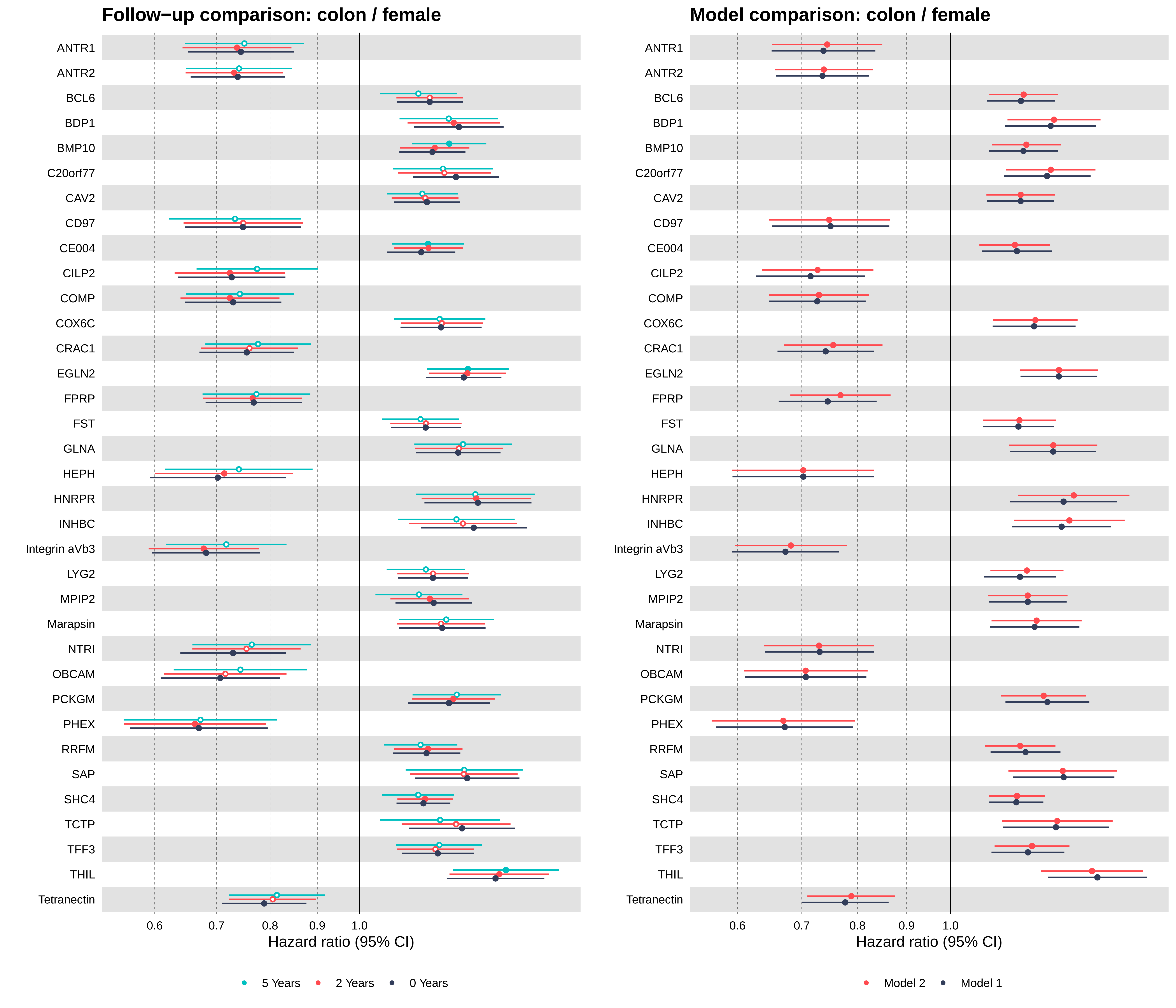
Follow−up comparison: colon / female
Model comparison: colon / female
ANTR1
ANTR2
BCL6
BDP1
BMP10
C20orf77
CAV2
CD97
CE004
CILP2
COMP
COX6C
CRAC1
EGLN2
FPRP
FST
GLNA
HEPH
HNRPR
INHBC
Integrin aVb3
LYG2
MPIP2
Marapsin
NTRI
OBCAM
PCKGM
PHEX
RRFM
SAP
SHC4
TCTP
TFF3
THIL
Tetranectin
0.6
0.7
0.8
0.9
1.0
Hazard ratio (95% CI)
5 Years
2 Years
0 Years
Model 2
Model 1

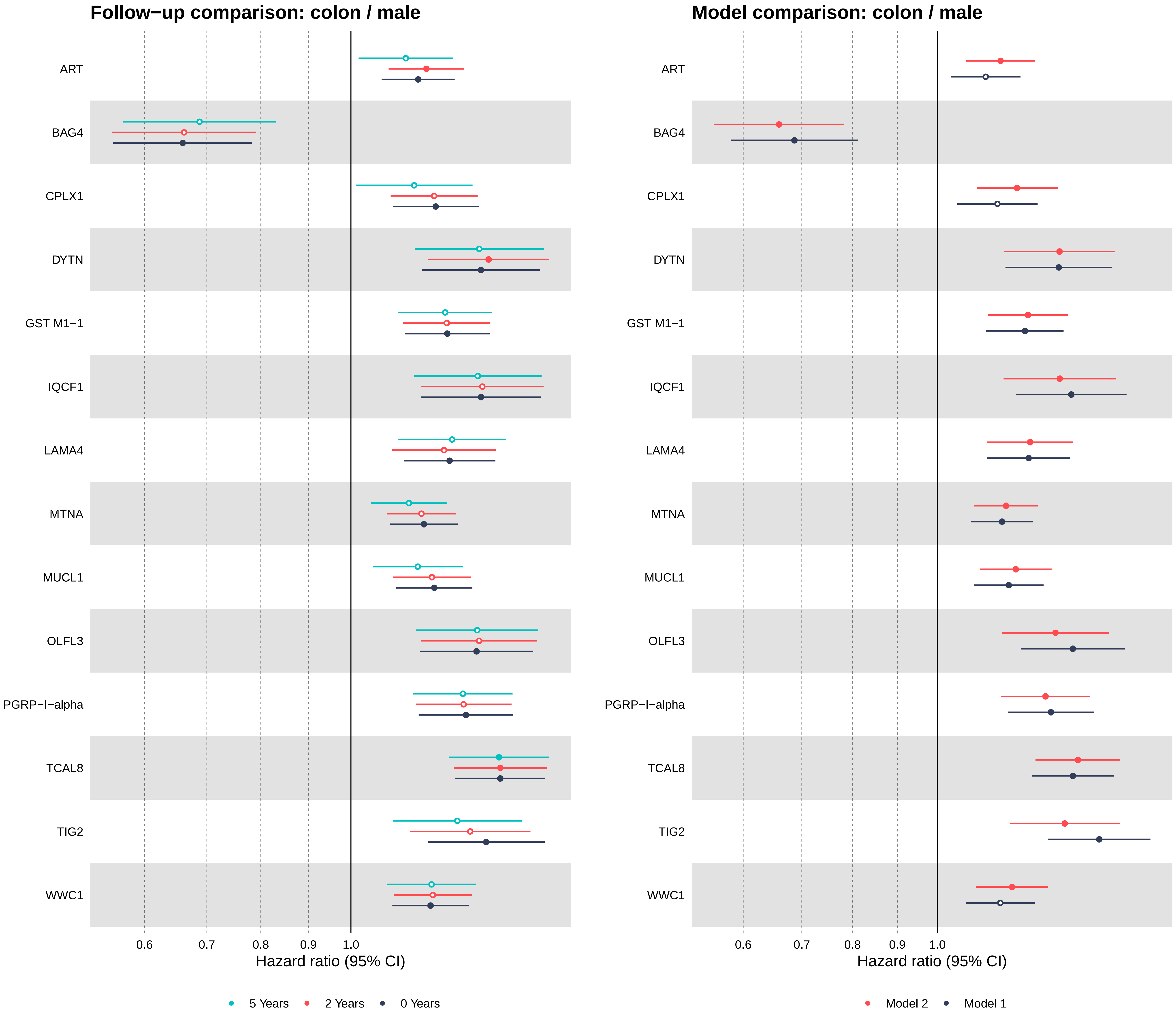

Follow−up comparison: colon / male
Model comparison: colon / male
ART
BAG4
CPLX1
DYTN
GST M1−1
IQCF1
LAMA4
MTNA
MUCL1
OLFL3
PGRP−I−alpha
TCAL8
TIG2
WWC1
0.6
0.7
0.8
0.9
1.0
Hazard ratio (95% CI)
5 Years
2 Years
0 Years
Model 2
Model 1

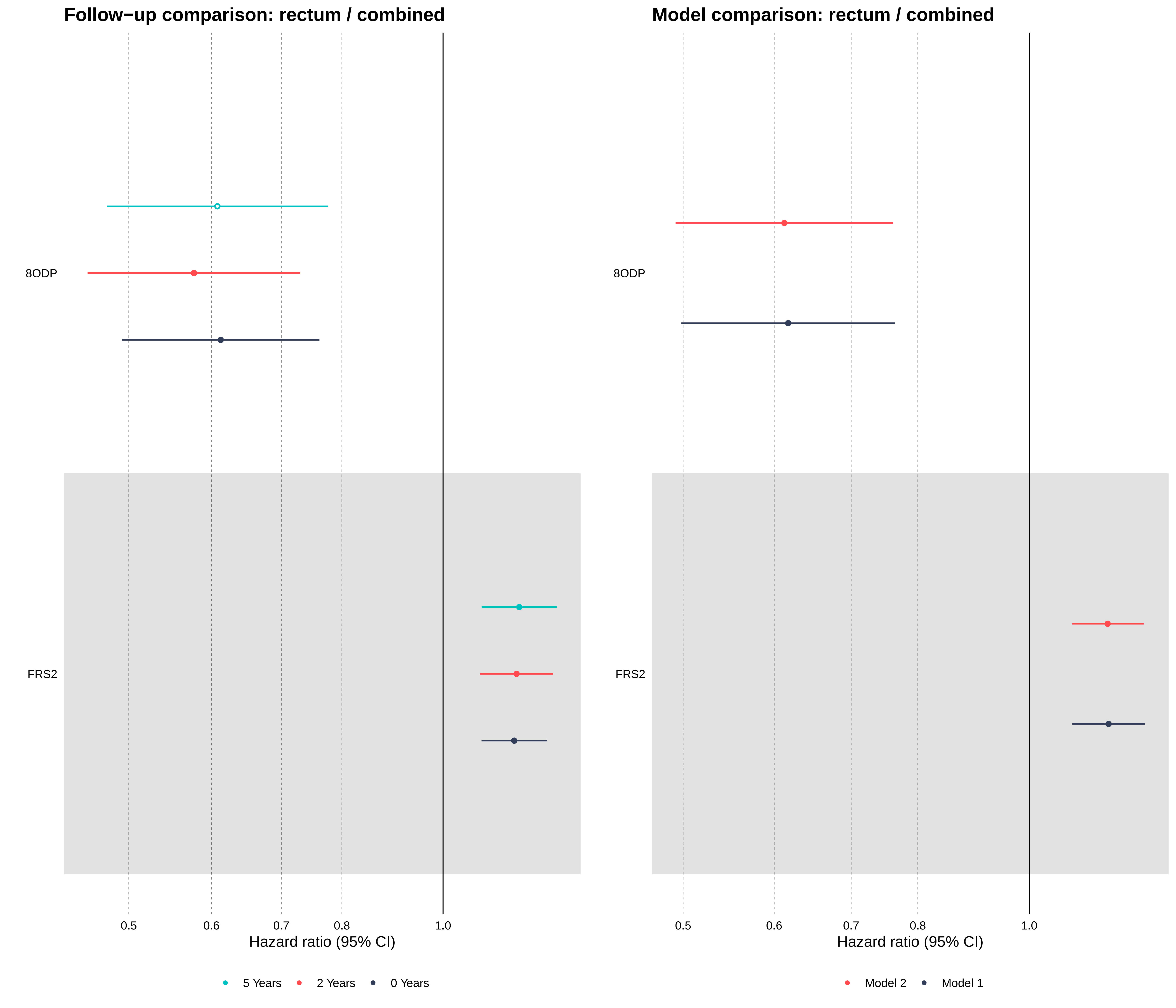
Follow−up comparison: rectum / combined
Model comparison: rectum / combined
8ODP
FRS2
8ODP
FRS2
0.5
0.6
0.7
0.8
1.0
Hazard ratio (95% CI)
0.5
0.6
0.7
0.8
1.0
Hazard ratio (95% CI)
5 Years
2 Years
0 Years
Model 2
Model 1

No FDR−significant proteins: rectum − female

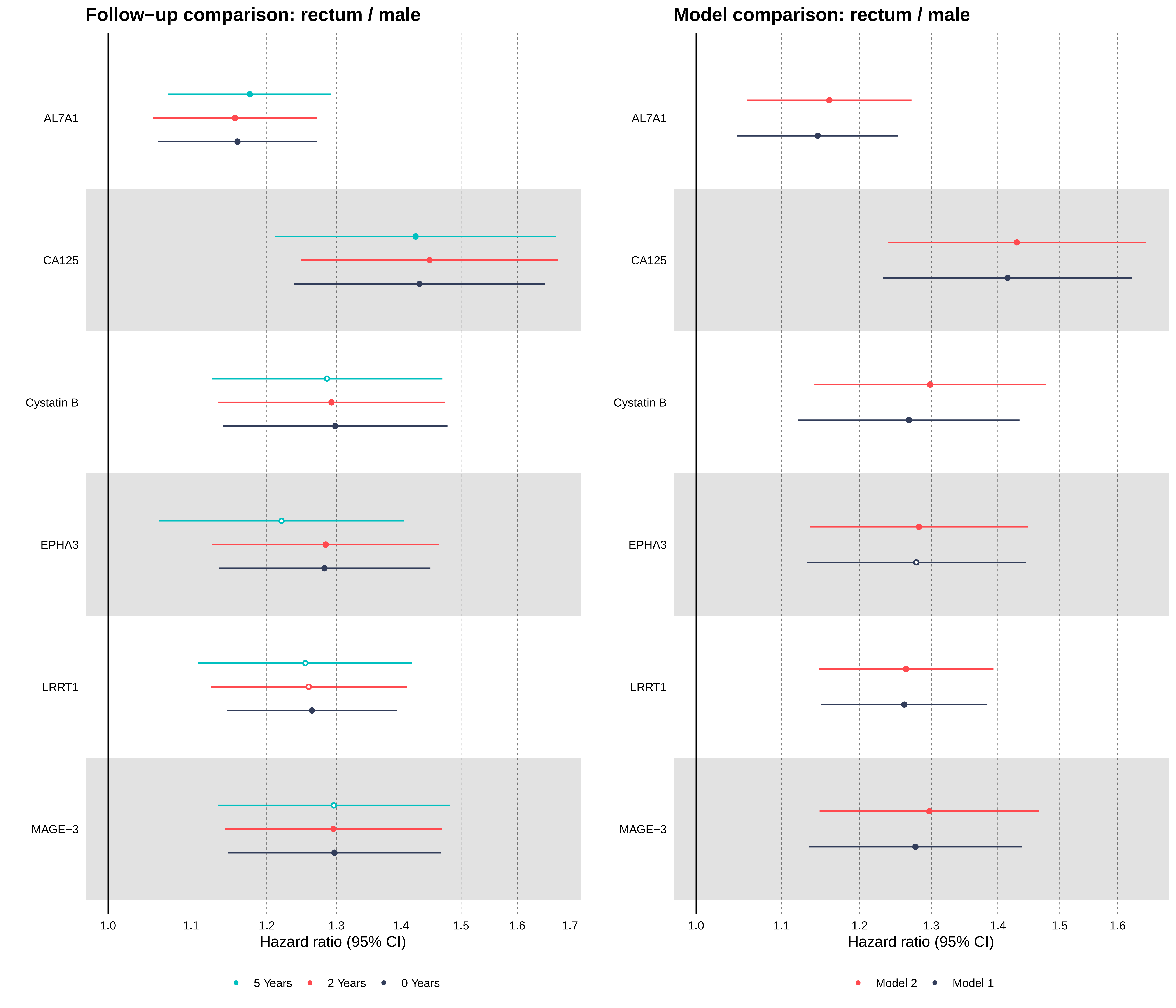
Follow−up comparison: rectum / male
AL7A1
CA125
Cystatin B
EPHA3
LRRT1
MAGE−3
1.0
1.1
1.2
1.3
1.4
1.5
1.6
1.7
Hazard ratio (95% CI)
5 Years
2 Years
0 Years
Model comparison: rectum / male
AL7A1
CA125
Cystatin B
EPHA3
LRRT1
MAGE−3
1.0
1.1
1.2
1.3
1.4
1.5
1.6
Hazard ratio (95% CI)
Model 2
Model 1

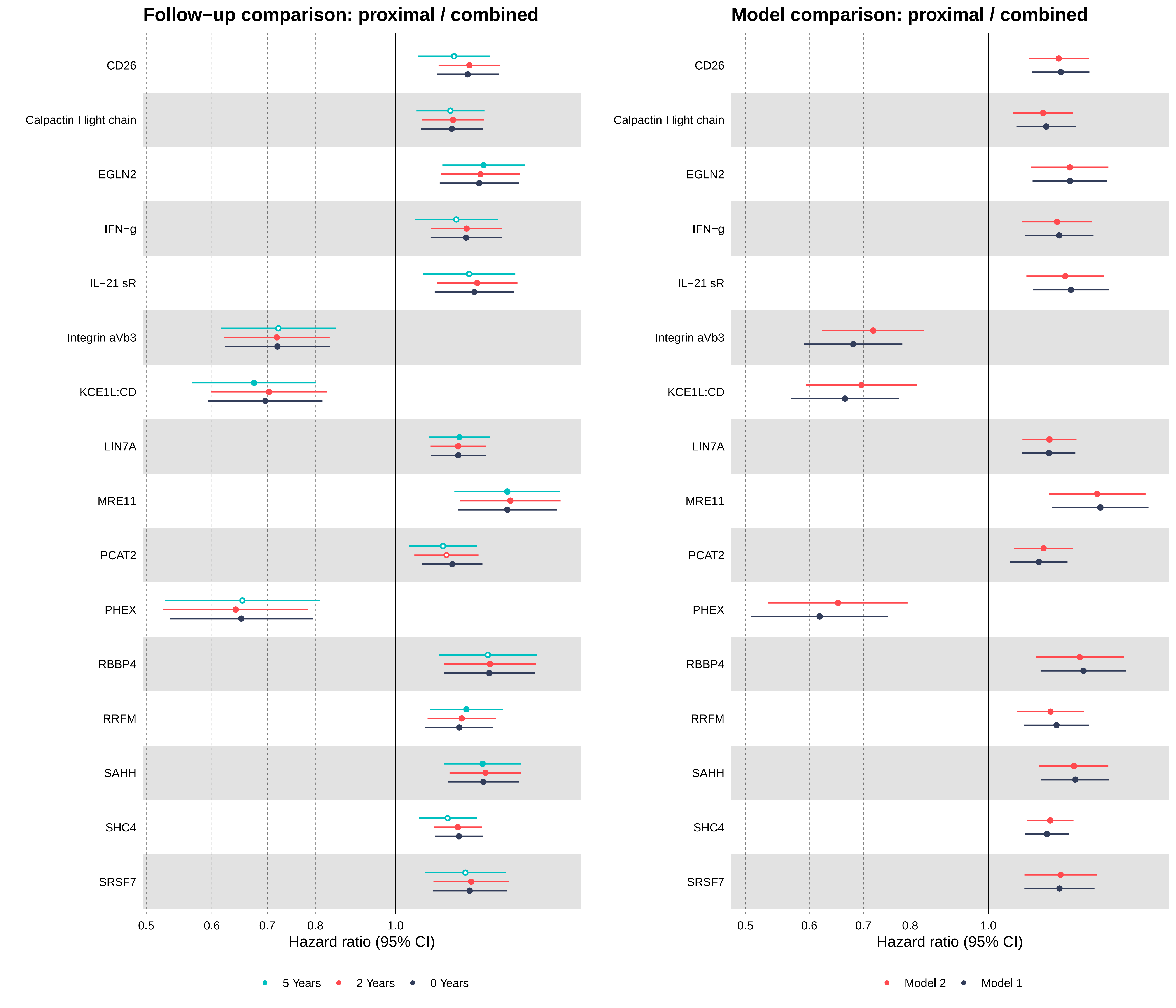
Follow−up comparison: proximal / combined
Model comparison: proximal / combined
CD26
Calpactin I light chain
EGLN2
IFN−g
IL−21 sR
Integrin aVb3
KCE1L:CD
LIN7A
MRE11
PCAT2
PHEX
RBBP4
RRFM
SAHH
SHC4
SRSF7
0.5
0.6
0.7
0.8
1.0
Hazard ratio (95% CI)
5 Years
2 Years
0 Years
Model 2
Model 1

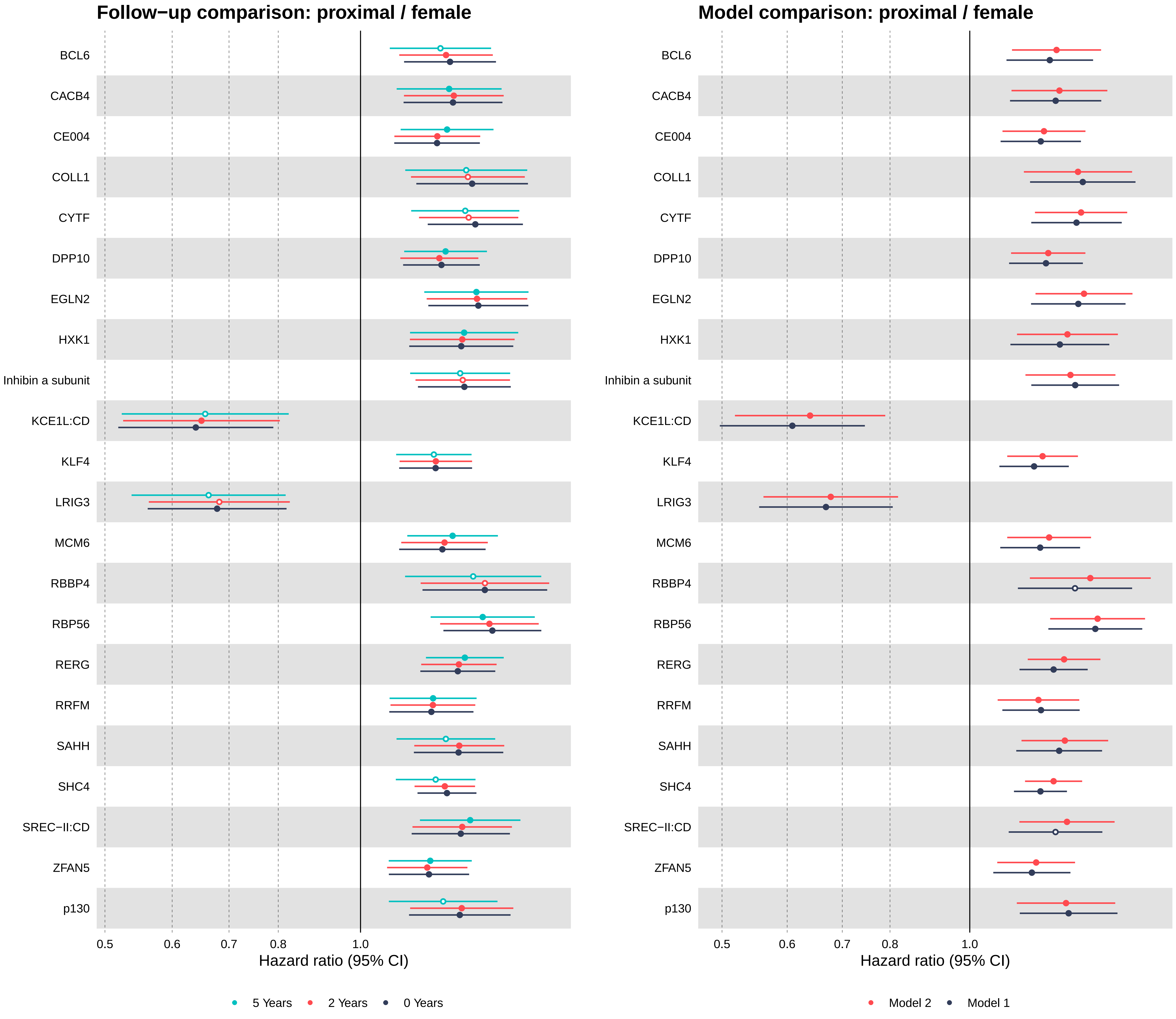

Follow−up comparison: proximal / female
Model comparison: proximal / female
BCL6
CACB4
CE004
COLL1
CYTF
DPP10
EGLN2
HXK1
Inhibin a subunit
KCE1L:CD
KLF4
LRIG3
MCM6
RBBP4
RBP56
RERG
RRFM
SAHH
SHC4
SREC−II:CD
ZFAN5
p130
0.5
0.6
0.7
0.8
1.0
Hazard ratio (95% CI)
5 Years
2 Years
0 Years
Model 2
Model 1

No FDR−significant proteins: proximal − male

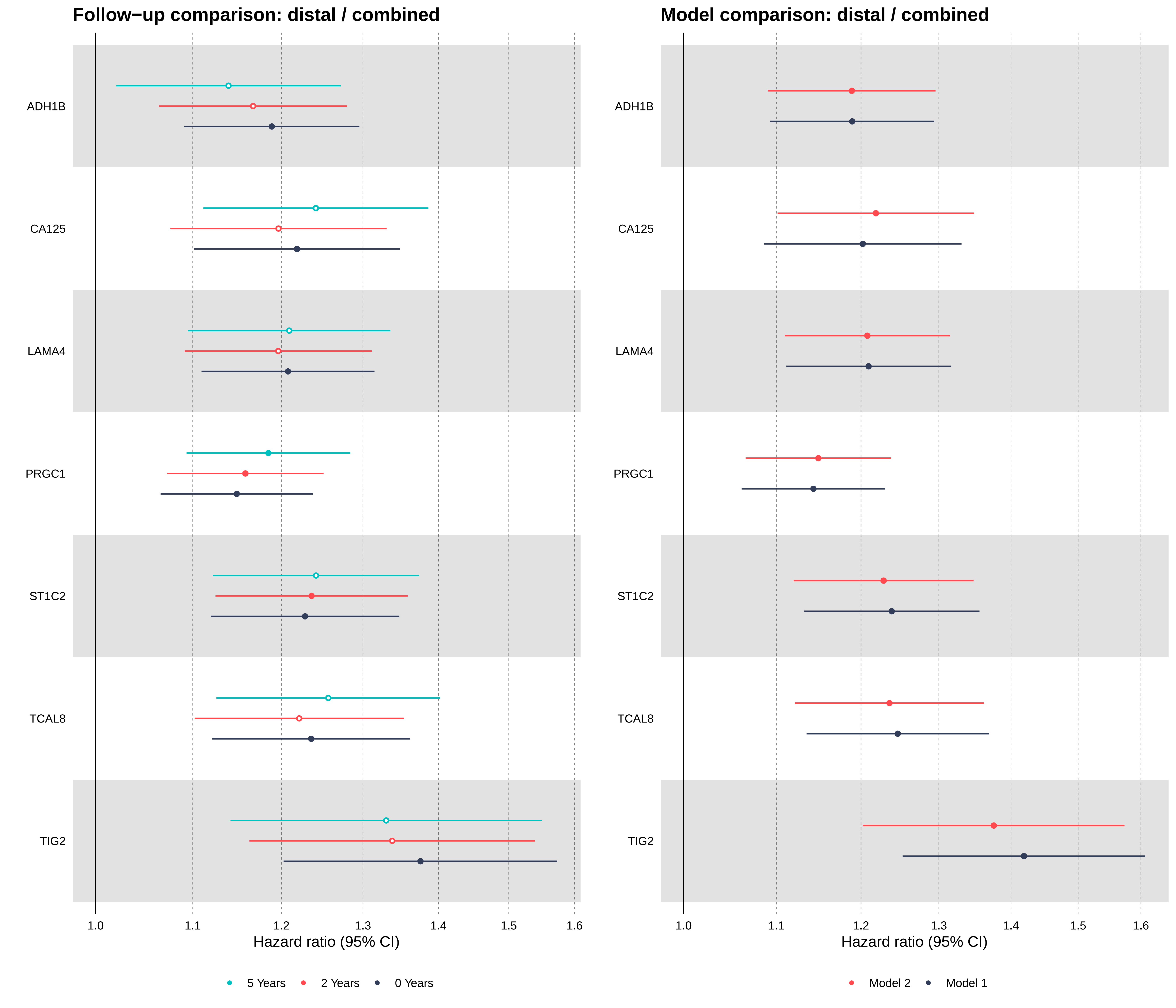
Follow−up comparison: distal / combined
Model comparison: distal / combined
ADH1B
CA125
LAMA4
PRGC1
ST1C2
TCAL8
TIG2
1.0
1.1
1.2
1.3
1.4
1.5
1.6
Hazard ratio (95% CI)
5 Years
2 Years
0 Years
Model 2
Model 1

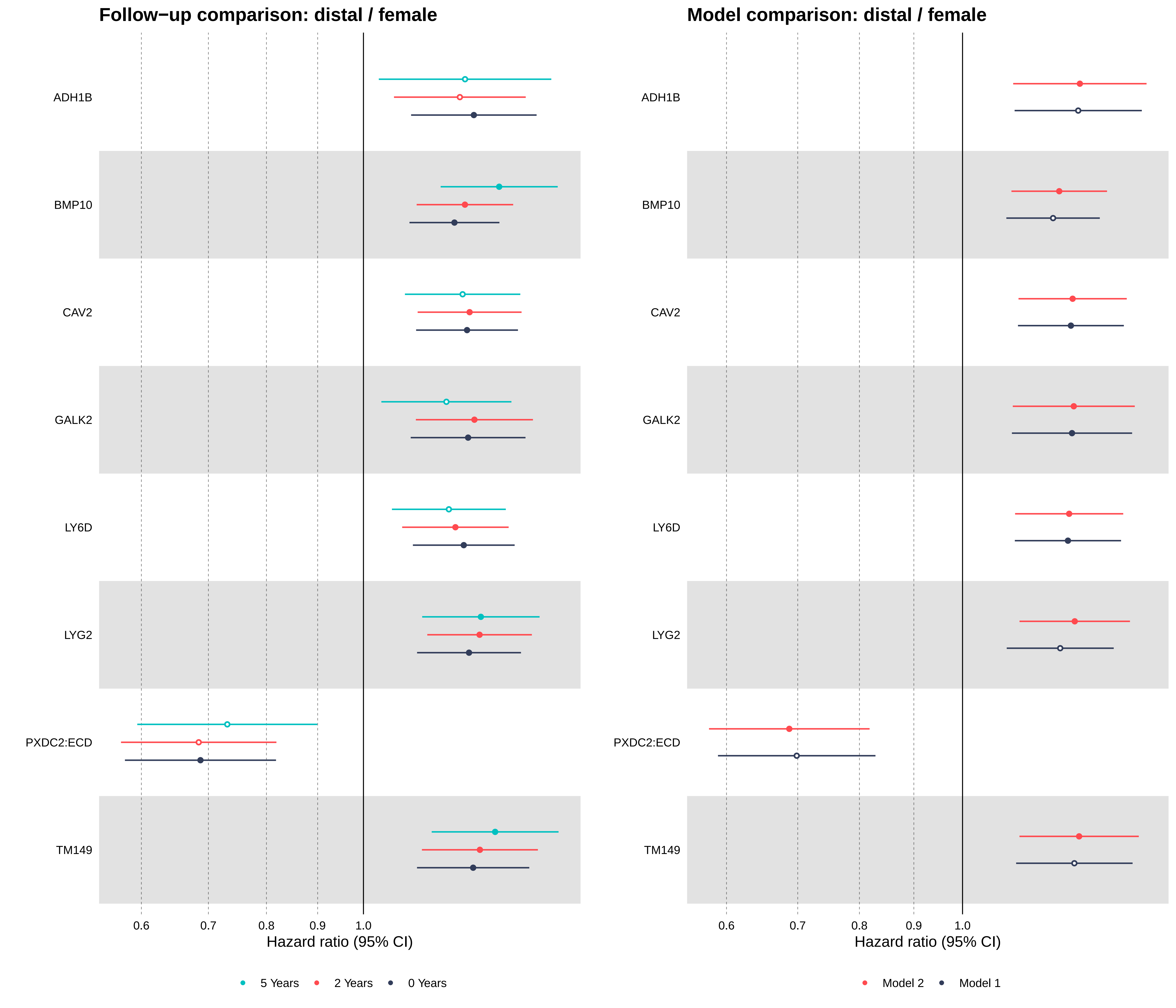

Follow−up comparison: distal / female
Model comparison: distal / female
ADH1B
BMP10
CAV2
GALK2
LY6D
LYG2
PXDC2:ECD
TM149
0.6
0.7
0.8
0.9
1.0
Hazard ratio (95% CI)
5 Years
2 Years
0 Years
Model 2
Model 1

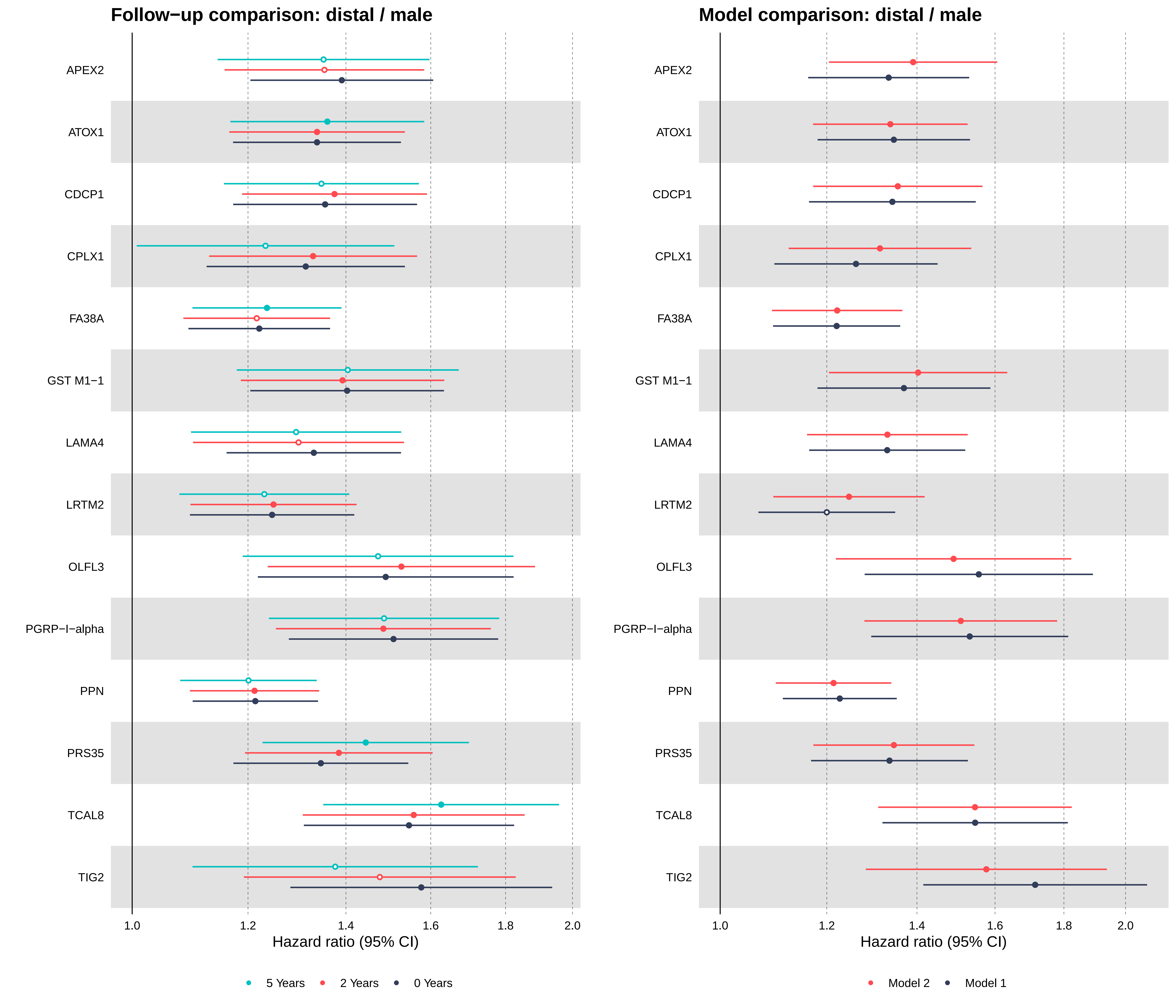

Follow−up comparison: distal / male
Model comparison: distal / male
APEX2
ATOX1
CDCP1
CPLX1
FA38A
GST M1−1
LAMA4
LRTM2
OLFL3
PGRP−I−alpha
PPN
PRS35
TCAL8
TIG2
1.0
1.2
1.4
1.6
1.8
2.0
Hazard ratio (95% CI)
5 Years
2 Years
0 Years
Model 2
Model 1
